# Expanded Chromatin Accessibility Mapping Explains Genetic Variation Associated with Complex Traits in Liver

**DOI:** 10.1101/2025.09.15.25335593

**Authors:** Brandon M. Wenz, Max F. Dudek, Shweta Ramdas, Kate Townsend Creasy, Dong Xin, Kim M. Olthoff, Abraham Shaked, Daniel J. Rader, Christopher D. Brown, Benjamin F. Voight

## Abstract

Genome-wide association studies (GWAS) have identified thousands of loci associated with a variety of common, complex human traits. Recent efforts have focused on characterizing chromatin accessibility to discover regulatory elements that modify the expression of nearby genes, suggesting that trait associations are mediated through changes in gene regulation. Genetic variants associated with differences in chromatin accessibility, known as chromatin accessibility quantitative trait loci (caQTLs), are established contributors to gene expression differences, providing mechanistic hypotheses for signals identified by GWAS. Using the assay for transposase-accessible chromatin with sequencing (ATAC-seq), we assessed chromatin accessibility in 189 diverse human liver samples, identifying over two million accessible chromatin regions enriched for gene regulatory features and, in 175 of these samples, over 14,000 caQTLs. Focusing subsequently on liver-relevant complex traits, we obtained publicly available blood lipids GWAS data and identified 157 loci where caQTLs, expression quantitative trait loci (eQTLs), and GWAS signals colocalized. This generated specific molecular hypotheses about regulatory elements, affected genes, and, in some cases, implicated transcription factors. Finally, we enumerated the set of blood lipid trait signals that lack an obvious proposed mechanism beyond catalogs of liver caQTLs and eQTLs. After integrating 10 multi-omic QTL regulatory mechanism datasets whilst considering limitations in statistical power, we found that approximately 20% of blood lipid GWAS signals lacked a statistical link to a proposed mechanism. Our results demonstrate the value of integrating multiple genomic datasets to improve understanding of GWAS signals, while emphasizing the need for additional experimental approaches to fully characterize complex trait associations.

## Introduction

Genome-wide association studies (GWAS) have made great strides forward, having identified thousands of genetic variants associated with complex human traits and disease^1–3^. Proving the correct, causal genetic mechanism underlying GWAS-implicated loci beyond reasonable doubt is a non-trivial endeavor, requiring detailed functional follow-up studies which have progressed at a much slower rate than locus discovery^4^. Many GWAS signals are found in blocks of linkage disequilibrium (LD) located in noncoding regions outside of core promoter elements and are hypothesized to regulate distal but nearby genes. As such, the central challenge is to pinpoint the specific causal variant(s) and mechanism, along with the target gene(s) that these association signals ultimately regulate^5,6^. A leading hypothesis is that noncoding variants identified by GWAS impact the function of gene regulatory elements and have downstream consequences on gene expression^7,8^. Leveraging the catalogs of variation associated with change in gene expression (i.e., expression quantitative trait loci, or eQTLs) generated by the Genotype-Tissue Expression (GTEx) Consortium^9^ along with many other sources^10–12^, trait-association signals appear to coincide with gene expression differences in ∼40-60% of loci examined via statistical colocalization^13,14^, generating a gap of mechanistic hypotheses from the loci that remain. It has been suggested that the incomplete overlap of eQTLs and GWAS could potentially result from systematic differences in the types of variants identified by these analyses^15^. Given the disparity between GWAS signals identified and those mechanistically characterized, advancing the understanding of causal variant mechanisms in post-GWAS experiments remains a key priority in the field.

Projects such as Encyclopedia of DNA Elements (ENCODE)^16^ and the NIH Roadmap Epigenomics^17^ consortium have made progress in constructing catalogs of functional elements and generation of epigenomic marks at scale that map the location of gene regulatory elements across the genome. Chromatin accessibility is another epigenomic characteristic associated with regulatory factor binding and gene regulation^18^, and can be measured to define gene regulatory elements within the noncoding genome. The degree of chromatin accessibility is affected by events such as transcription factor (TF) binding^19^, a key step in gene expression regulation^20^. Functional genetic variants may alter TF binding, resulting in chromatin accessibility changes^21^ and subsequent effects on gene expression, ultimately contributing to their causal role in complex trait heritability. A more thorough investigation of genetic variants that affect chromatin accessibility (i.e. caQTLs) will improve our understanding of the genetic mechanisms and architecture underlying complex trait-associated loci.

The liver has emerged as a central tissue where genetic variation identified by GWAS influences susceptibility to many medically-relevant diseases^22^. The liver performs hundreds of vital functions in humans, including removing waste products from the bloodstream, controlling blood sugar levels, and making essential nutrients^23^. The liver also plays a central role in the regulation and metabolism of blood lipids^24,25^. Blood lipids are heritable risk factors contributing to traits such as coronary artery disease (CAD), and GWAS have identified hundreds of genetic variants associated with blood lipid levels across diverse ancestries^26^. While chromatin accessibility in the liver has been used to characterize genetic variants associated with disease, many current caQTL studies have limited sample size^27^. Increasing the number of samples with chromatin accessibility measurements is essential for enhancing caQTL discovery power and advancing our understanding of GWAS mechanisms^28^, ultimately towards closing the missing mechanistic hypothesis gap.

A thorough understanding of GWAS loci and trait-associated variants is crucial for downstream applications and for understanding the regulatory architecture of complex human traits. To address the current limitations of post-GWAS studies, we conducted a joint analysis of the effects of genetic variants on liver gene expression and chromatin accessibility to quantify their combined contribution to complex human traits. We identified hundreds of loci where common genetic variants implicated gene regulatory elements and target genes that likely share a causal variant with GWAS signals for liver-relevant traits. Furthermore, we quantified the remaining GWAS signals whose mechanisms were not explained by effects on gene expression or chromatin accessibility in the liver, proposed potential mechanisms for these signals, and reported the proportion of liver-relevant GWAS signals that remain unresolved. These results highlight the need for future studies and the key considerations necessary for their design.

## Results

### Comprehensive chromatin accessibility profiling in human liver identifies millions of open chromatin regions

We collected 189 pre-transplantation donor liver samples from the Penn Transplant Institute and performed ATAC-seq to characterize variation in chromatin accessibility in this population. With these data, we identified 2,518,633 open chromatin peaks across all samples (**Methods**) and initially sought to characterize their properties. We found that the median peak length was 292 base pairs and peak regions cumulatively covered approximately 41% of the genome. The peak length distribution proved multimodal, with the highest proportion of peaks from presumed nucleosome-free regions, as well as peaks that are likely to be derived from open chromatin regions that span a nucleosome or multiple nucleosomes (**Supplementary Figure 1**). We found that the majority of peaks were located in intronic and intergenic regions (∼68% of annotations; **Supplementary Figure 2**). Compared to matched random controls, ATAC-seq peaks were significantly enriched in many annotation categories with the most prominent enrichment in FANTOM enhancer regions^29,30^ and gene 5’UTRs. In contrast, peak regions were significantly depleted in intergenic regions and gene 3’UTRs (**Supplementary Figure 3, Table S1**). Based on liver sample epigenomic annotations from the NIH Roadmap Epigenomics Consortium^17^, we found that ATAC-seq peaks were significantly enriched for all tested histone marks except H3K4me3, a marker of active promoters (**Supplementary Figure 4, Table S2**). Prior work in human liver tissue has identified thousands of putative gene regulatory elements based on the epigenetic marks H3K27ac and H3K4me3^31^. Open chromatin regions that we identified in the human liver using ATAC-seq were enriched 1.86-fold and 1.83-fold in H3K27ac and H3K4me3 regions, respectively, compared to size-matched, randomly selected regions (permuted P < 0.001 for both histone marks, **Table S3, Methods**). The distances from each peak to the nearest protein-coding gene transcription start site (TSS) followed a log-normal distribution (**Supplementary Figure 5**), with a median distance of 82,184 base pairs (bp). Additionally, 17,370 peaks directly overlapped their closest protein-coding gene TSS. These characteristics and enrichments support the model that peaks of accessibility we have identified are consistent with putative cis-regulatory elements broadly across the genome in the liver.

### Thousands of variants are associated with chromatin accessibility in the human liver

To identify genetic factors influencing chromatin accessibility differences at open chromatin peaks, we mapped chromatin accessibility quantitative trait loci (caQTLs) in 175 samples with available genotype data (**Methods**)^32^. Specifically, we identified caQTLs that exert a local influence on chromatin accessibility (cis-caQTLs) by testing for variants that are associated with chromatin accessibility within the ATAC-seq peak itself, or in a cis window extending 10 kilobases (kb) on either side of the peak boundaries. We utilized a method that considers both population and allele-specific signals and identified 14,076 caQTLs at a 5% false discovery rate (FDR) (**Figure 1A, Table S4, Methods**). The rate of replication of lead caQTLs and peaks identified in our study was strong (π_1_ replication = 0.73, **Methods**)^33,34^ based on a previously published liver caQTL study with fewer samples^27^ (**Supplementary Figure 6**). caQTL features were significantly enriched in several genomic annotation categories when compared to randomly selected regions, including gene promoters and FANTOM enhancer regions, suggesting enrichment for regions likely to regulate gene expression (**Supplementary Figure 7, Table S5**). caQTL features, however, were significantly depleted in several categories, including introns and 3’ UTRs. We also compared caQTL peaks to histone marks defined by the Epigenomic Roadmap in liver samples, observing enrichments for H3K9me3, H3K9ac, and H3K4me1 (**Supplementary Figure 8, Table S6**). H3K9ac and H3K4me1 are markers of promoters and enhancers, however, H3K9me3 is often associated with heterochromatin. Significant depletions were observed for H3K27me3, associated with heterochromatin, and H3K36me3, associated with gene bodies. Additionally, compared to all peaks, we found that caQTL peaks were found closer to protein-coding gene transcription start sites (median distance = 55,361 bp) (**Supplementary Figure 9**). Together, these results support the role of caQTL peaks in gene regulation.

**Figure 1.**
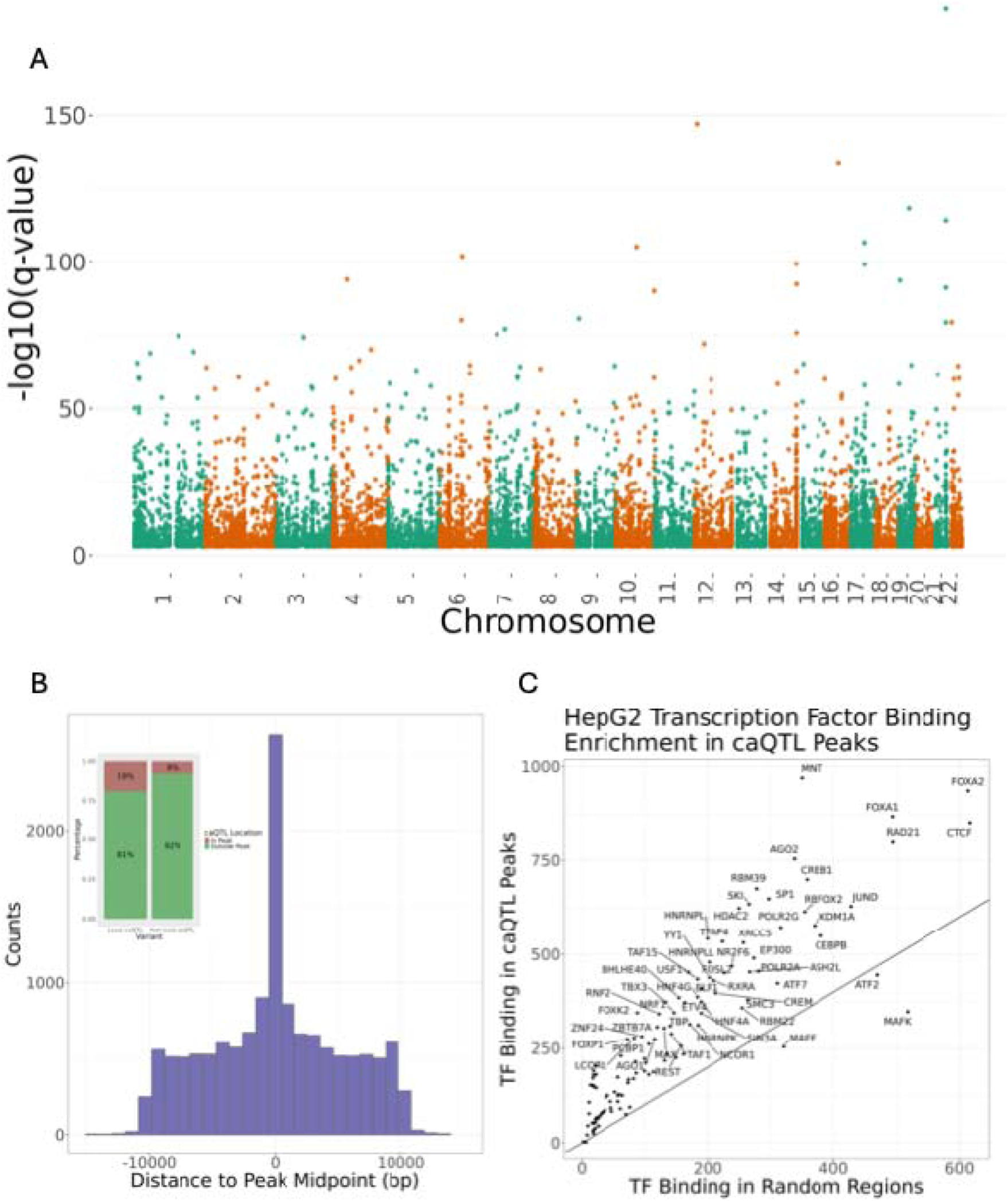
Thousands of caQTLs identified in potential gene regulatory regions. **(A)** 14,076 autosomal caQTL peaks were identified across 175 samples. **(B)** Lead caQTL variants were enriched within the peak that they are associated with. **(C)** caQTL peaks are enriched/depleted for transcription factor binding sites in HepG2 ChIP-seq.

One potential mechanistic model underlying caQTLs is that genetic variants might influence transcription factor (TF) occupancy, which can be measured through differences in chromatin accessibility at gene regulatory elements. To examine this, we quantified the distance between the lead caQTL and the center of the associated caQTL peak. Consistent with this model, we find that these 14,076 lead caQTLs are enriched within the chromatin accessibility peak they are associated with (Odds Ratio (OR) = 2.87, 95% Confidence Interval (95% C.I.) = 2.75 – 2.99; P < 2.2 x 10^-16^), suggesting a direct link between caQTLs and regulatory element activity (**Figure 1B**). To assess the extent to which disrupted TF binding might contribute to caQTL mechanisms, we leveraged chromatin immunoprecipitation with sequencing (ChIP-seq) TF binding evidence for 119 TFs in the liver model HepG2 cell line. We demonstrated that, compared to matched controls, caQTL peaks that we identified are enriched for transcription factor binding events, many of which are enriched across lipid traits^25^, as well as depleted for other TFs (**Figure 1C, Supplementary Figure 10, Table S7**). Through cis-caQTL mapping in human liver tissue, we showed that genetic variation significantly influences chromatin accessibility at putative regulatory elements enriched for promoter and enhancer marks, likely through effects on transcription factor binding.

The samples that we collected for ATAC-seq included individuals from diverse genetic backgrounds. We leveraged this diverse collection of samples by performing caQTL mapping in a subset of individuals with non-European ancestry. Specifically, we identified 26 individuals whose genetically inferred ancestry was similar to populations collected by the 1000 Genomes Project from continental Africa (**Methods**). We performed caQTL mapping in these 26 samples and identified 2,088 caQTLs at a 5% FDR (**Supplementary Figure 11**). Of the 2,088 caQTLs identified, 833 (∼40%) were also found in the analysis of all samples. As expected, the concordance rate between sets that either included or excluded those shared with the overall analysis was observed to be very high (π_1_=0.75 or π_1_=0.64, respectively, **Supplementary Figures 12-13**)^33,34^. Examples of caQTL peaks specific to the cohort with African ancestry illustrate the increased power to detect caQTLs in populations with distinct allele frequency distributions **(Supplementary Figure 14).** To evaluate the consistency of the direction of effect for the African ancestry specific caQTL peak-variant pairs in the full dataset, we performed a binomial sign test and found a significant concordance between the African ancestry and full sample datasets (P < 2.2 × 10L¹L), with approximately 85% of caQTL summary statistic pairs sharing the same direction of effect (**Methods**). While the generation of ancestry-specific results, particularly in understudied groups, are important for downstream analyses^35^, in what follows, we utilize caQTL results using all samples for downstream analyses to maximize statistical power.

### Gene expression differences are driven by chromatin accessibility in cis-regulatory elements in the liver

caQTLs identify potential gene regulatory elements but do not, on their own, identify target genes. To assess evidence of shared causality between variants that affect chromatin accessibility and variants that affect gene expression, and to nominate caQTL target genes, we performed colocalization^36^ analyses between caQTLs and eQTLs from GTEx V8 Liver data. We identified a total of 2,996 colocalizations between 1,832 unique chromatin accessibility peaks and 1,035 unique genes (**Table S8**). Of these colocalization events, 164 had the same eQTL and caQTL lead variant.

### Closing the gap of missing mechanisms using chromatin accessibility data

Armed with a catalog of variation associated with chromatin accessibility, we next turned to address the extent to which these data can provide hypothesis linking variation associated with complex traits relevant to the liver to an underlying mechanism. Toward this end, we first performed colocalization analyses between liver caQTLs and a range of complex human traits with known or suspected liver involvement (**Methods**). Across 18 tested traits (**Table S9**), we identified 980 colocalizations involving 526 unique caQTL peaks and 500 unique GWAS signals spanning 15 of these traits (**Table S10**).

In addition to effects on chromatin accessibility, GWAS variants may also be associated with gene expression. We quantified the extent of signal sharing between GWAS signals and eQTLs by performing colocalization between GTEx V8 liver eQTLs and GWAS signals. Across these 18 traits, we found between 1 and 175 colocalizations per trait, for a total of 1059 colocalizations involving 685 unique sentinel GWAS variants, 469 unique eGenes, and 449 unique lead eQTL variants (**Table S11**). Notably, eQTL colocalization identified putative regulatory mechanisms for 1.49-fold more GWAS signals compared to caQTL colocalizations (PLJ=LJ1.7LJ×LJ10LJ¹³; ORLJ=LJ1.60, 95% CI: 1.41–1.81). Overall, both liver eQTLs and caQTLs colocalized with a median of approximately 25% of GWAS signals across these 18 traits (**Figure 2**).

**Figure 2.**
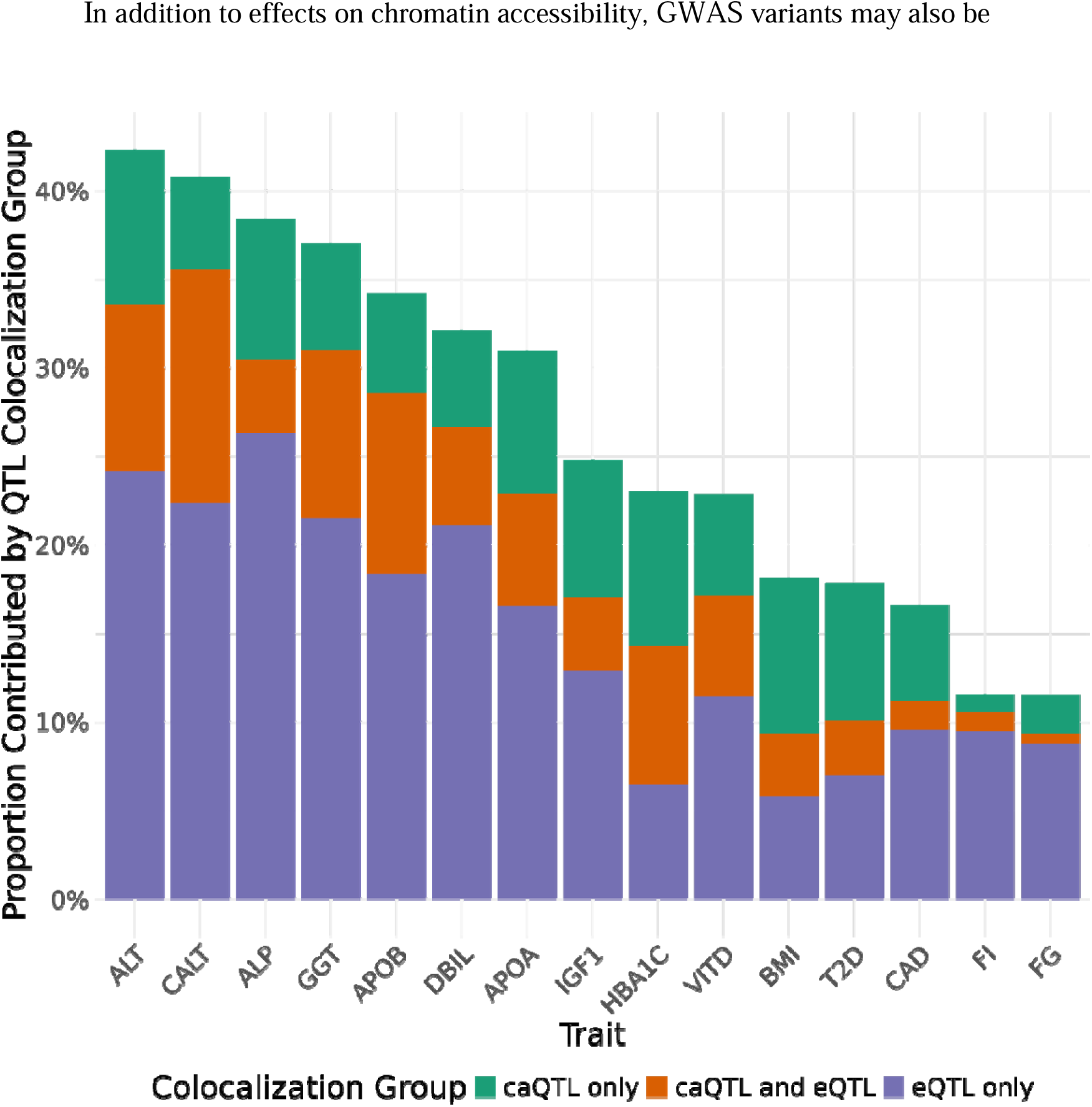
caQTLs and eQTLs colocalize with complex trait-associated loci. Across 18 traits that likely involve liver biology, we found a range of colocalizations **(Methods)** for both caQTLs and eQTLs. eQTLs explain 1.49-fold more GWAS signals compared to caQTLs. Only traits with colocalizations that pass filters shown. ALT, Alanine Aminotransferase; CALT, Chronic ALT; ALP, Alkaline Phosphatase; GGT, Gamma-Glutamyl Transferase; APOB, Apolipoprotein B; DBIL, Direct Bilirubin; APOA, Apolipoprotein A; IGF1, Insulin-like Growth Factor 1; HBA1C, Hemoglobin A1c; VITD, Vitamin D; BMI, Body Mass Index; T2D, Type 2 Diabetes; CAD, Coronary Artery Disease; FI, Fasting Insulin; FG, Fasting Glucose.

### A subset of blood lipids GWAS signals are not explained by eQTLs or caQTLs

Although the liver is involved in a variety of physiological processes, it is particularly important in lipid metabolism. To evaluate evidence of signal overlap between variants that affect chromatin accessibility and blood lipid traits, we next focused on colocalization analyses using GWAS summary statistics from the Global Lipids Genetics Consortium (GLGC)^26^ for five lipid traits: HDL cholesterol (HDL), LDL cholesterol (LDL), total cholesterol (TC), triglycerides (logTG), and nonHDL cholesterol (nonHDL). Given the large proportion of caQTL samples are similar to individuals from 1000 Genome collected from continental Europe, we first searched for shared causal variants between caQTLs and lipid trait GWAS signals identified in the GLGC European ancestry cohort. We identified a total of 287 colocalization events involving a caQTL peak and GLGC blood lipid GWAS signals (**Table S12**). To establish a baseline of colocalization events between GLGC lipid trait signals and regulatory variants, we compared the results of our caQTL/GWAS colocalizations with the results of GTEx V8 liver eQTL/GWAS colocalizations (**Figure 3**). We found a total of 547 colocalization events between eQTLs and GLGC blood lipid GWAS signals (**Table S13**). Overall, liver eQTLs explained 1.90-fold more European ancestry GLGC lipid GWAS signals through colocalization than liver caQTLs (PL=L1.3L×L10L¹²; ORL=L2.14, 95% CI: 1.72–2.67).

**Figure 3.**
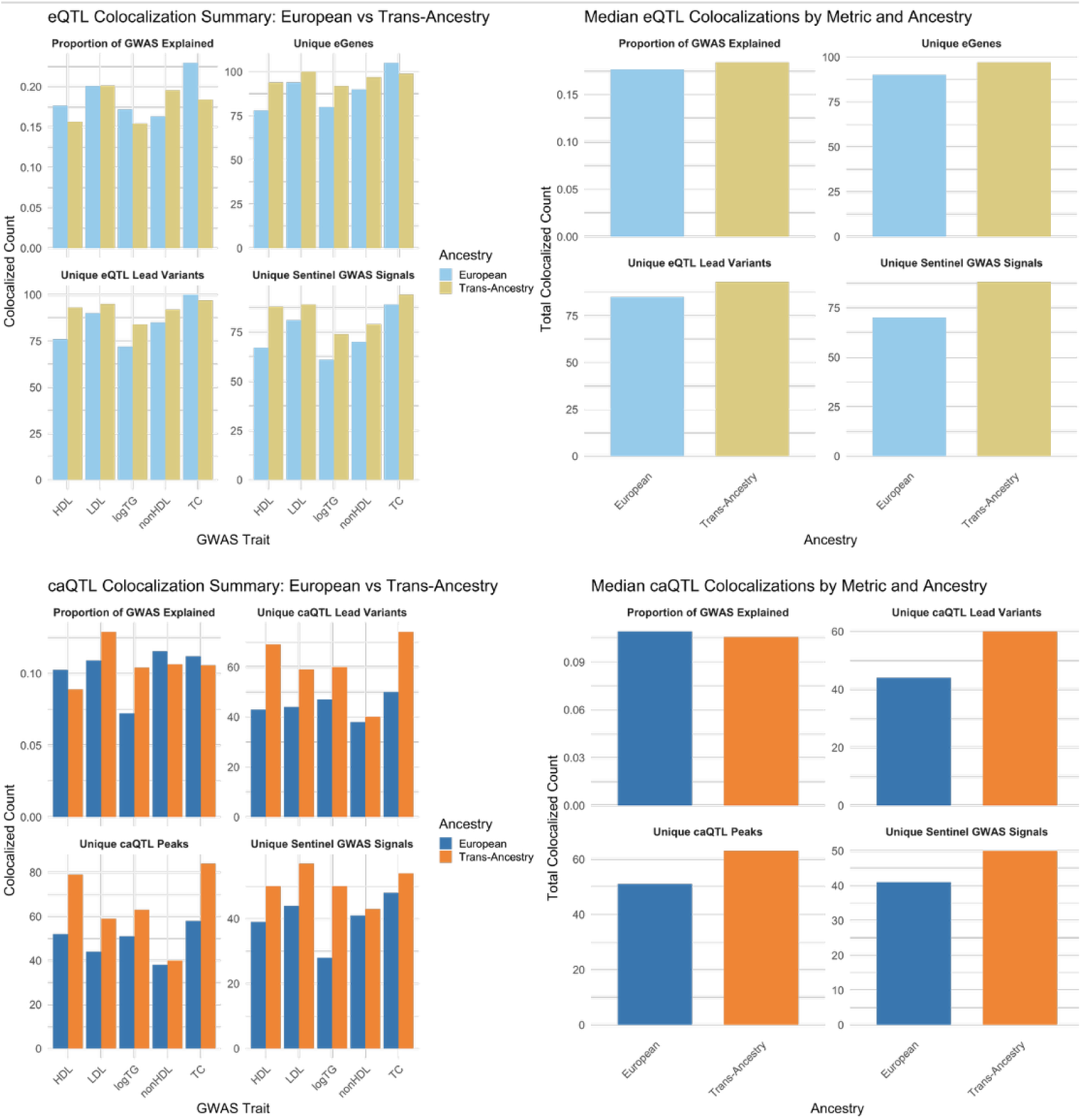
caQTLs and eQTLs colocalize with blood lipids associated loci, a key liver-related trait. Across five blood lipid traits that primarily involve the liver, we find a range of colocalizations for both caQTLs and eQTLs in both European and trans-ancestry sample cohorts (**Methods**). Through colocalization analyses, eQTLs explain 1.76- to 1.90-fold more GWAS signals compared to caQTLs.

The samples that we performed caQTL mapping in also included individuals with non-European ancestries. To leverage this genetic diversity and the increased power of trans-ancestry GWAS, we performed colocalization analyses between liver caQTLs and trans-ancestry GLGC lipid traits GWAS signals and identified 398 colocalization events involving a caQTL peak and GLGC lipid phenotype (**Table S14**). We also assessed the gain in discovery power when considering shared causal signals between trans-ancestry GLGC lipid GWAS signals and GTEx V8 liver eQTLs and found a total of 606 colocalizations (**Table S15).** These results suggest that, compared to caQTLs, eQTLs explained 1.76-fold more trans-ancestry GWAS signals through colocalization analyses (PLJ=2.8 x 10^-12^; ORLJ=LJ1.95, 95% CI: 1.61–2.37; **Figure 3, Supplementary Figure 15**). Overall, these results highlight the added value of performing GWAS in populations with non-European ancestry.

### Increased discovery power in eQTL mapping explains additional GWAS signals

The power to detect QTLs is influenced by several factors, including sample size^37^. A recent study meta-analyzed four eQTL mapping datasets in human liver tissue, resulting in a combined sample size more than five times larger than the GTEx liver eQTL dataset^38^. To determine the extent to which caQTLs and these eQTLs share a causal signal, we performed colocalization analyses and found a total of 1,679 colocalizations between 1,287 unique chromatin accessibility peaks and 948 unique genes (**Table S16**). Of these colocalization events, 56 involved a shared lead variant between the eQTL and caQTL. We assessed the sharing of liver caQTL colocalization results across both liver eQTL datasets and found significant overlap of shared caQTL peaks and genes (**Supplementary Figures 16-17**). We additionally considered the correlation of effect size and direction between colocalizing liver eQTLs and caQTLs with shared lead variants, finding a significant positive relationship (PL=L2.46L×L10LL; rL=L0.28, 95% CI: 0.15–0.40; **Supplementary Figure 18**).

To determine how increased eQTL mapping power impacts the number of GWAS signals explained through colocalization, we performed colocalization analyses between the meta-analyzed liver eQTL dataset and trans-ancestry GLGC lipid GWAS traits. Across all five lipids traits, we found a total of 998 colocalization events, representing a 1.65-fold increase compared to GTEx eQTLs (PL<L2.2L×L10L¹L; ORL=L1.95, 95% CI: 1.67–2.29; **Supplementary Figure 19, Table S17**). We again compared results obtained from the two eQTL datasets and found significant overlap at both the gene and sentinel GWAS variant levels (**Supplementary Figures 20-21**). Through these colocalization analyses, we demonstrated that increased eQTL mapping power led to greater discovery of shared causal variants between regulatory elements and GWAS loci, highlighting the importance of sample size and statistical power in revealing molecular mechanisms underlying complex traits.

### Joint consideration of multiple molecular QTLs underlying GWAS signals suggests causal pathways at lipids GWAS loci

Colocalizations that occur jointly across GWAS, eQTLs, and caQTLs are of particular interest, as these allow for the development of specific hypotheses at a given locus regarding causal variants, regulatory mechanisms, and target genes contributing to downstream phenotypes. To maximize three-way colocalization discovery, we integrated results from both eQTL datasets across the five GLGC lipids traits and identified a total of 298 unique three-way colocalizations, including 49 shared trait–gene–peak–sentinel GWAS signal combinations across both eQTL datasets (**Supplementary Figure 22, Tables S18-S20**). We leveraged these colocalizations involving multiple traits to answer several fundamental questions about gene regulation in the human liver. Loci implicated in GWAS can be complex, and a single open chromatin region can often colocalize with multiple genes simultaneously. To address how common this phenomenon is, we assessed how many of the identified three-way colocalizations involve a single gene. Of the 298 three-way colocalization events, 75 implicated a single gene. These instances may enable more accurate prediction of causal mechanisms and target genes at GWAS loci (**Supplementary Figure 23**).

Gene expression regulation is often a complex process, as regulatory elements do not always interact with the nearest gene and can regulate multiple genes.^39^ Three-way colocalizations involving a single gene allow for the identification of the likely causal gene at a locus. We used this information to identify whether the putative regulatory element colocalizes with the nearest gene, based on proximity to gene transcription start sites (TSSs). Interestingly, of the three-way colocalizations involving a single protein-coding gene, we found that the gene regulatory element colocalized with the nearest gene only 21% of the time. We found that in cases where colocalization does not involve the nearest gene, the median number of intervening genes was five (**Supplementary Figure 23**). This result suggests that causal genes underlying GWAS signals sometimes, but not always, implicates the nearest one to association signals.

### Integration of multiple lines of evidence recapitulate known mechanisms and suggest novel regulatory mechanisms at GWAS loci

To assess the validity of our experiments and workflow, we examined a well-known locus where a genetic variant disrupts TF binding, affecting hepatic gene expression of the sortilin 1 (*SORT1*) gene, blood lipid levels and myocardial infarction (MI) risk.^41^ A single common noncoding variant, rs12740374, has been fine-mapped as the causal variant through creation of a C/EBP TF binding site by the minor allele. Consistent with our hypothesis, an ATAC-seq peak overlaps this causal variant and exhibits genotype-specific chromatin accessibility, with the same direction of effect as TF binding experiments and *SORT1* gene expression (**Supplementary Figures 24-25**).

Our pipeline also resulted in many novel loci where colocalizations occurred between eQTLs, caQTLs, and GWAS signals. One such example is an LDL-associated signal on chromosome 1 that colocalized with an eQTL for the gene *PRMT6* and a 2,107 base pair (bp) caQTL peak that spans the entire coding sequence of exon 1 of this single-exon gene. (**Figure 4A**). We generated billions of sequencing reads in our ATAC-seq experiments across the 189 human liver samples. Using these data, we performed footprint analysis^40^ to identify regions where active TF binding caused localized decreases in chromatin accessibility within peaks, revealing the binding locations of hundreds of TFs in our human liver samples (**Supplementary Figure 26**). We leveraged these footprints to infer the potential causal transcription factor at GWAS loci, enhancing the resolution of our experiments by identifying the specific TFs that may drive chromatin accessibility and gene expression differences, enabling a more detailed mechanistic understanding of GWAS associations. For example, through TF footprinting analysis, the lead caQTL variant, rs2232015, is predicted to overlap a *THAP11* binding site. The A allele of rs2232015 is associated with increased chromatin accessibility and increased *PRMT6* gene expression, and is in strong linkage with the LDL-decreasing G allele of the sentinel GWAS variant rs1977658 (r^2^=0.96) (**Supplementary Figure 27**). *PRMT6* has been shown to play a key role in hepatic lipid homeostasis, with its inhibition resulting in reduced lipid accumulation in primary hepatocytes^42^. Another example is a triglycerides-associated locus on chromosome 9, with lead variant rs150611042, that colocalized with an eQTL for the *ORM2* gene in the GTEx liver eQTL and with caQTL peak 2498643 (**Figure 4B**). However, at this locus, the eQTL and caQTL had opposite directions of effect, suggesting a more complex regulatory mechanism (**Supplementary Figure 28**). This caQTL peak spans a 3,565 bp region that encompasses the entirety of the *ORM1* gene. Additionally, a liver eQTL from the meta-analyzed liver dataset colocalized with this triglycerides signal and caQTL peak (**Supplementary Figure 29**). The lead variant of this caQTL peak, rs113354603, falls within intron 2 of the *ORM1* gene. *ORM2* has recently been implicated in hepatic lipid homeostasis due to its role in lipogenesis^43^. In this case, the caQTL might suggest that *ORM1* is the causal gene, given the location of the lead variant and caQTL peak. Loci such as this, where multiple lines of evidence support a regulatory mechanism, are strong candidates for downstream functional follow-up experiments.

**Figure 4.**
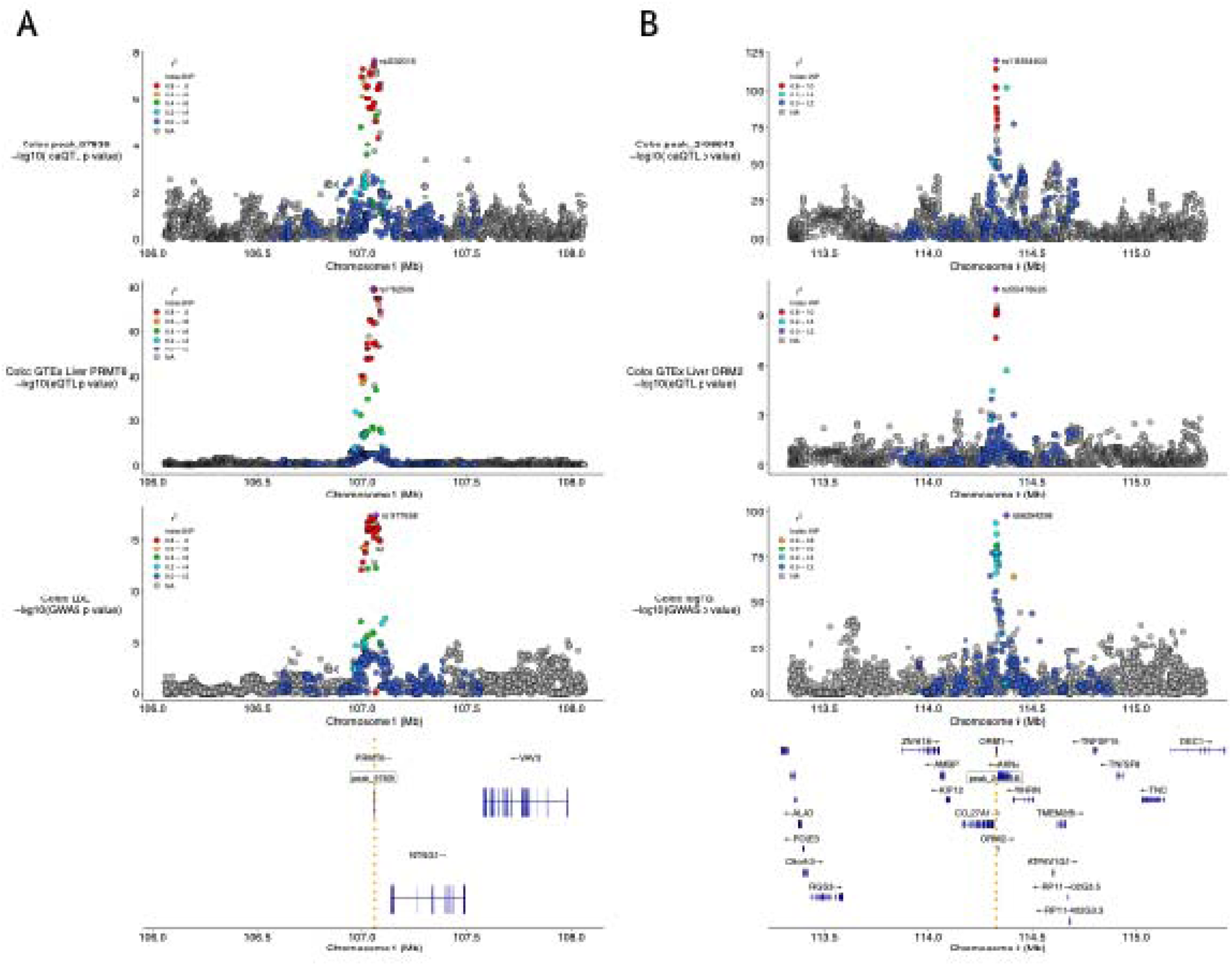
Novel mechanisms are inferred at blood lipids GWAS loci. **(A)** Colocalization plot for an LDL signal on chromosome 1. Gene *PRMT6* and caQTL peak 97839 both colocalize with the GWAS signal at this locus. Through TF footprint analysis, lead caQTL rs2232015 is predicted to overlap THAP11 TF binding site. **(B)** Colocalization plot for a triglycerides signal on chromosome 9. Gene *ORM2* and caQTL peak 2498643 both colocalize with the GWAS signal at this locus.

### Unresolved mechanisms remain for a minority of blood lipid GWAS signals

GWAS signals identified by GLGC might be explained by mechanisms beyond chromatin accessibility or gene expression. Given the catalog of different classes of molecular phenotype data in liver, including what we collected here as well as the vast repository of liver-relevant data in the public domain, we next sought to quantify the contribution of various functional genetic variants to these traits. Although most GWAS signals are noncoding variants, a subset of GWAS sentinel variants may have coding consequences. With respect to noncoding variation, current liver eQTL and caQTL datasets that we leveraged are likely underpowered, and large non-liver eQTL datasets may contribute to blood lipid traits, particularly in tissues that have been connected to blood lipid trait biology^12,44^. Beyond coding mechanisms and caQTLs/eQTLs, which together suggested a mechanism for approximately 45% of blood lipids GWAS signals (**Supplementary Figure 30**), additional noncoding regulatory processes may explain a subset of the remaining signals. To comprehensively characterize the GLGC blood lipid GWAS signals, we applied a stepwise framework to assign each sentinel variant to a mutually exclusive explanatory category. We sequentially evaluated the contribution of exonic variants^45^, eQTLs and caQTLs from liver and other tissues^12,46^, QTLs for protein levels (pQTLs)^47^ and splicing isoform usage (sQTLs)^48^, as well as the potential contribution of QTLs from non-steady-state adult tissues and underpowered liver caQTLs and eQTLs (**Supplementary Figure 30**).

Across all GLGC traits, a total of 138 signals (76 unique), were annotated as exonic, comprised of 132 nonsynonymous SNVs and 6 stopgain mutations. Although only about 6% of sentinel variants are coding, the functional effects of these variants can be much easier to predict (**Figure 5**). Next, we identified 119 sentinel variants that colocalized with liver caQTLs, 701 with eQTLs, and 122 with both liver eQTLs and caQTLs. Collectively, these noncoding regulatory mechanisms propose a hypothesis for approximately 39% of the total identified GWAS sentinel variants (**Figure 5**), while more than half of the sentinel GWAS signals we analyzed remained without a proposed mechanism. Extending our analysis to consider blood eQTLs from a large meta-analysis of 31,684 individuals^12^ provided a hypothesis for an additional 343 unique GWAS sentinel variants (**Figure 5**). Of the remaining GWAS signals not previously linked to a regulatory mechanism, 14 colocalized with liver sQTLs and a single GWAS signal was explained using liver pQTLs from a subset of quantifiable protein levels (**Figure 5**). Subsequently, an additional 111 unique GWAS signals colocalized with caQTLs identified in a large analysis performed across diverse cell and tissue types (**Figure 5**). Phenotypic measurements in primarily adult steady-state tissues and cell lines will not be able to capture all the specific contexts of regulatory variants. We considered two additional datasets – QTLs contributing to rhythmic gene expression (rhyQTLs) in liver^49^ and eQTLs identified in fetal-like iPSC-derived cardiovascular progenitor cells (CVPCs)^50^. Through colocalization analyses, these studies provided an explanation for 109 and 14 GWAS signals, respectively.

**Figure 5.**
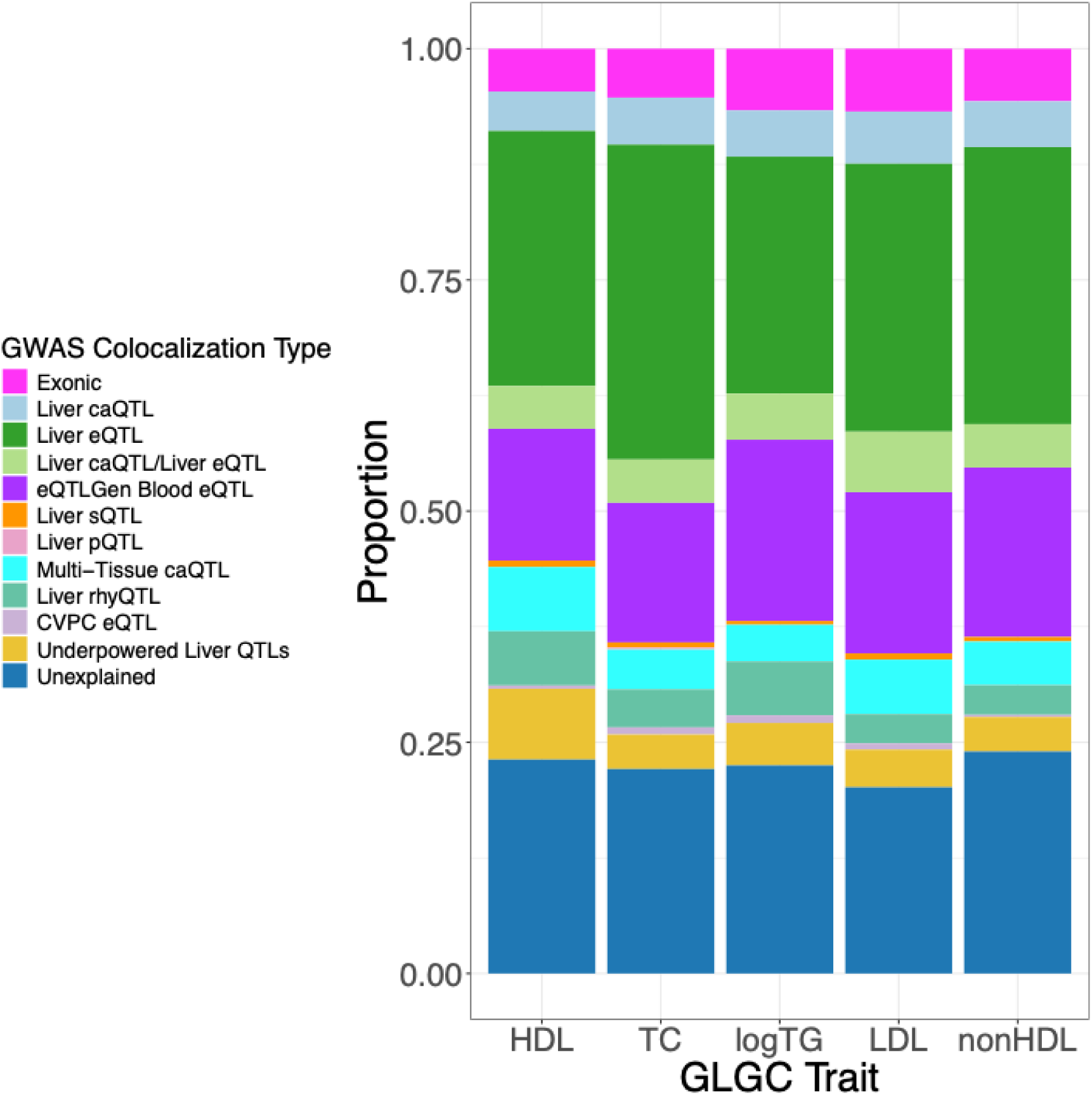
Nearly a quarter of GWAS signals remain unresolved, despite a plethora of data. Considering all investigated mechanisms, ∼22% of GWAS signals remaining without a proposed hypothesis. Sentinel variants were classified in a mutually exclusive manner, following the order shown in the legend.

Finally, we predicted the contribution of underpowered liver eQTLs and caQTLs by identifying the strongest, but not statistically significant, eQTL/caQTL signals that overlapped a remaining GLGC GWAS variant without a mechanistic explanation. While these QTLs did not reach statistical significance, their p-value distribution was skewed towards small p-values, suggesting that better powered QTL studies might elevate some of these QTLs to significance (**Supplementary Figures 31-32**). Considering both sets of underpowered liver QTLs, we identified 156 unique GWAS signals for which a mechanism might be hypothesized with larger sample sizes (**Figure 5**). These results underscore the need to generate more data from larger sample sizes to fully advance post-GWAS studies. Overall, across five GLGC lipids traits, and considering various regulatory and coding mechanisms, we proposed a hypothesis for approximately 78% of GWAS signals identified (**Figure 5, Supplementary Figures 33-37, Table S21**).

### Analysis of unresolved GWAS associations suggests contributing factors

We sought to identify potential reasons why many GWAS signals remain without a mechanistic hypothesis after considering a multitude of datasets. We first examined whether colocalization results are associated with the strength of the GWAS signal. We hypothesized that GWAS signals without a proposed mechanism through our analyses might have effects that are smaller in magnitude than exonic signals or those that colocalized with various molecular phenotypes. For each GWAS variant across the five GLGC phenotypes, we calculated the non-centrality parameter (NCP) of the association test statistic to enable power estimation. We categorized sentinel GWAS variants into three groups based on their calculated discovery power statistic and annotated each variant with its proposed mechanism through colocalization, if available. Overall, we found that most GWAS signals had high discovery power, with similar colocalization representation across power groups. Notably, even among GWAS signals with moderate to high discovery power, approximately 25% remained unlinked to a regulatory mechanism. We did note that GWAS signals without a proposed mechanism had a larger proportion in the lowest power bin relative to other groups, with pairwise comparisons showing significant differences between low vs. mid (PL=L0.0438) and low vs. high (PL=L0.0035) power bins (**Supplementary Figure 38, Methods**). The lowest power bin additionally contained lower porportions for GTEx liver eQTLs but higher proportions for eQTLs identified by eQTLgen. While hard to assess unambiguously, these trends are consistent with a model whereby GWAS signals with lower power in discovery presumably due to weaker effect sizes are linked with eQTLs, but with lower eQTL effect sizes such that much larger sample sizes are required for discovery.

It has been suggested that gene regulatory variants, such as eQTLs, do not provide a hypothesis for many GWAS signals due to systematic biases in discovery, including the observation that GWAS hits are further from the nearest TSS compared to eQTLs^15^. We investigated whether the distance to the nearest protein-coding gene TSS differed among GWAS signals that colocalized with different categories of regulatory variants in our study and those that did not colocalize at all (**Figure 6, Supplementary Figures 39-40**). We found that the GLGC blood lipids GWAS variants that did not colocalize in our study were the most distal from the nearest TSS (median = 46,290 bp), whereas those colocalized with liver eQTLs were more than twice as close (median = 16,823 bp). GWAS signals that colocalized with both liver eQTLs and liver caQTLs (median = 15,190 bp) were closer than GWAS signals that colocalized with liver caQTLs alone (median = 21,471 bp). These findings suggest that caQTLs may be more effective in identifying distal regulatory elements contributing to the genomic architecture of complex traits. Functional regulatory variant discovery power likely also plays a role, as variants from the larger eQTL and caQTL datasets colocalized with GWAS signals located farther from TSSs compared to those identified in the smaller liver-specific datasets.

**Figure 6.**
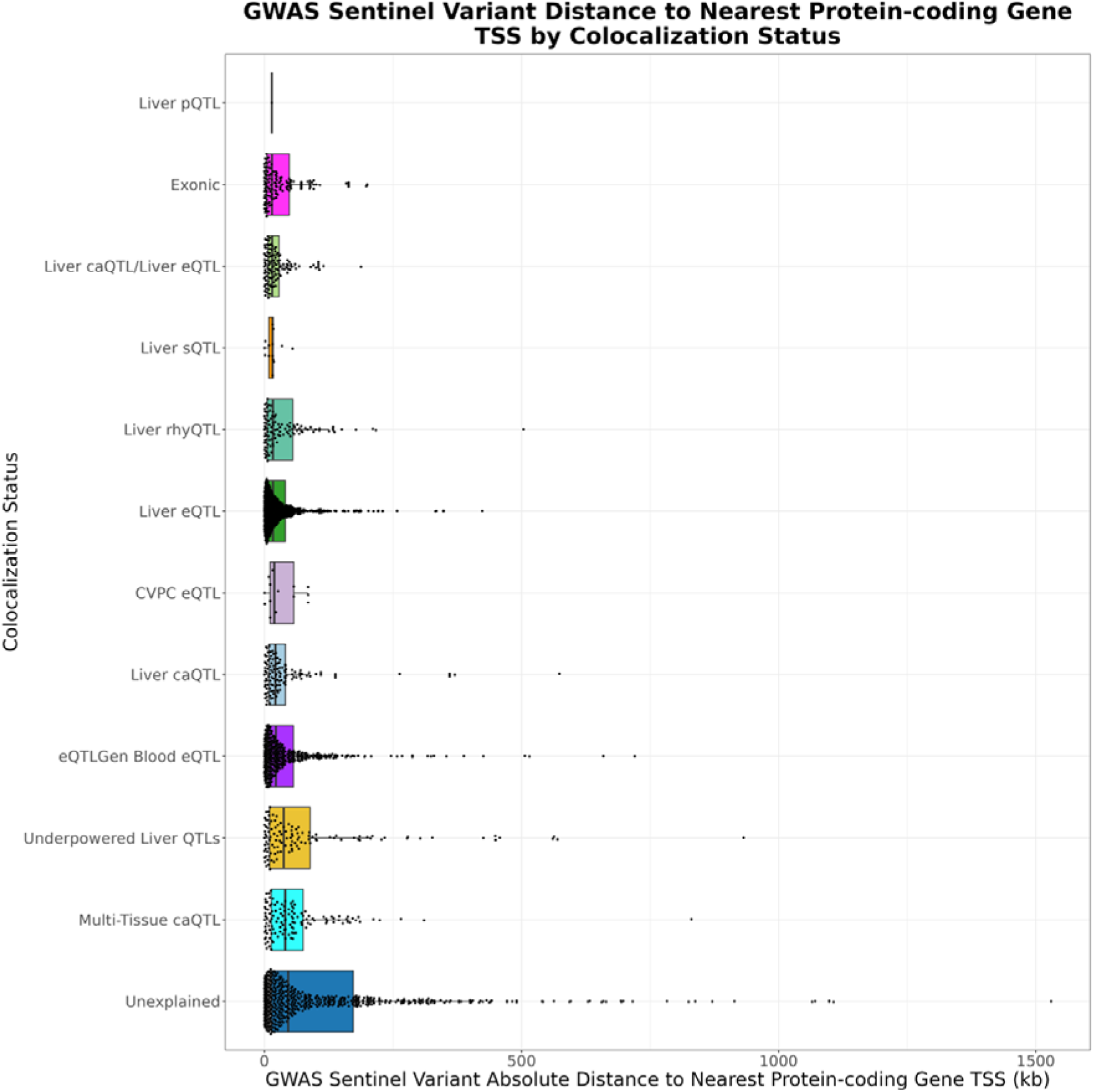
GWAS signals that do not colocalize with other traits tend to be located farther from protein-coding genes. For each GWAS signal, we calculated the distance to the nearest protein-coding gene TSS and grouped signals by colocalization status. As expected, exonic variants were closest, whereas non-colocalizing variants tended to be farthest away.

Although we performed exhaustive analyses to propose mechanisms for GLGC GWAS signals, 537 variants (∼22%) remained without a hypothesized mechanism. Allelic heterogeneity is a common phenomenon across molecular QTLs and GWAS loci. However, many pairwise colocalization techniques using summary statistics assume a single causal variant for each trait, which may lead to a lack of colocalization at loci where multiple causal signals are present. Colocalization results between GWAS signals that remained without a proposed mechanism and liver QTLs were often dominated by PPH3 in the colocalization results, which suggests distinct causal variants underlie the GWAS and QTL signals (**Supplementary Figures 41-42**). Loci with high PPH3 could also be the result of complex LD patterns or allelic heterogeneity, which could cause false negative colocalization results, warranting further investigation (**Supplementary Figure 43)**. Differences in GWAS population ancestry can also lead to false negatives in our colocalization analyses due to ancestry-specific risk variants or population-specific LD structure. To determine if differences in ancestry contributed to lack of colocalizations, we examined sentinel variants lacking a mechanistic hypothesis that were shared between the European-only and trans-ancestry GWAS analyses. We identified five such variants that colocalized only in the European cohort. The presence of multiple causal variants presents a more challenging problem to solve with many colocalization tools. To explore the contribution of this to our results, we manually reviewed colocalization Manhattan plots to identify loci with apparent allelic heterogeneity. We then assessed LD between the lead QTL and significant GWAS variants (P < 5 × 10LL) at the locus. We identified 33 GWAS signals with r² > 0.5 with a lead QTL, where visual inspection suggested the presence of multiple causal variants. After accounting for potential colocalization failures due to allelic heterogeneity, 467 GWAS signals (∼19%), representing 364 unique variants, still lacked a proposed mechanistic hypothesis (**Table S22**).

The remaining sentinel signals without a proposed mechanism present an opportunity to examine their properties and better understand why they may be difficult to classify. Notably, some GWAS signals that remained without a proposed mechanism were very significant and had high discovery power (**Supplementary Figure 44**). We first considered whether these variants were found in liver open chromatin regions identified in our ATAC-seq experiment and found that 282 (∼60%) overlapped a peak. Mechanistically uncharacterized GWAS signals that overlapped ATAC-seq peaks were signficantly closer to protein-coding gene TSSs (median = 98,112 bp) than those that did not (median = 179,111). As 40% of blood lipids GWAS signals without a proposed mechanism were not found in liver open chromatin regions identified in our ATAC-seq experiment, this suggests that they may not be active in adult human livers and/or may have more distal gene regulatory effects.

## Discussion

We performed ATAC-seq on 189 healthy human liver samples, identified regions of accessible chromatin across these samples, and generated a novel, large caQTL dataset from a subset of individuals with genotype data. We integrated this liver-specific caQTL dataset with existing datasets from liver and other tissues to quantify the proportion of current GWAS sentinel variants with a hypothesized mechanism. We highlighted several examples where a confluence of data suggests a comprehensive mechanism at GWAS loci, prioritizing genetic regulatory variants for functional follow-up experiments. Overall, we utilized colocalization analyses to generate mechanistic hypotheses for nearly 75% of blood lipid GWAS variants.

Although we integrated various lines of evidence, we still cannot propose a mechanistic hypothesis for all GWAS variants, even for traits (e.g., cholesterol) where profiling in the relevant tissue (e.g., liver) has been extensive across molecular phenotypes. Several factors may contribute to this residual gap. A strength of the study lies in the number of individuals profiled; however, achieving this scale required collection in bulk tissues from adults (and further, postmortem). Although most cells in the liver are hepatocytes^51^, functional regulatory variants might be specific to other, rarer cell types. Single-cell approaches could potentially tease out the role of variants on chromatin accessibility, gene expression, and other traits in specific cellular contexts which may be masked in our bulk analyses. A second potential cause is that regulatory variants might only be active in the presence of specific environmental factors or developmental cues^52–54^. For example, a recent study found that ∼5% of tested GWAS signals were found to colocalize only in specific developmental cell types^50^. This result suggests that defining a mechanism for every GWAS variant might require broader approaches to data collection beyond adult tissues or in the context of perturbed states^55,56^. Finally, one statistical limitation of many colocalization methods is the assumption of a single causal variant at each locus. It has been shown that multiple functional regulatory variants in strong LD can underlie GWAS signals^57^ and molecular QTLs^58^, leading to potential false negatives in our analyses.

In summary, we combined novel caQTL mapping results with publicly available data to learn about the regulatory architecture of complex human traits. This combination of data enabled an effective and efficient workflow that identified genetic variants that are likely involved in gene regulatory pathways for prioritization in functional follow-up assays. We also summarized contribution of existing datasets and highlighted the need to generate new, larger datasets across diverse cell types and biological contexts. Our expanded catalog of liver caQTLs provides a public resource for integration in future experiments and workflows. Importantly, these hypotheses will need to be verified with functional follow-up experiments in various model systems to establish causality.

## Methods

### Sample Collection

The study utilized data collected from specimens obtained from liver transplant recipients from their respective donor cohorts at the University of Pennsylvania, collected either 2012-2017 and 2018-2020 enrolled under the BioTIP study (Biorepository of the Transplant Institute at the University of Pennsylvania). Participants were enrolled in the prospective biorepository and clinical databases, collecting biological samples and clinical data at the time of transplantation, and at predetermined intervals after transplantation. The study was approved by the University of Pennsylvania’s Institutional Review Board (2012-2017: FWA 00004028, protocol 820091; 2018-2020: FWA 00004028, protocol 814870). All research was conducted in accordance with both the Declarations of Helsinki and Istanbul. The legal representatives of the organ donors signed informed consent prior to explantation and inclusion in this study. Specimens collected from this protocol used in this study were deidentified and subsequently anonymized.

### ATAC-Seq Library Generation

Human liver wedge biopsies were supplied by the Penn Transplant Institute. Samples were derived from human livers deemed fit for transplantation and were collected at the time of the surgery. Samples were flash frozen and stored at -80 C. Chromatin accessibility profiles were generated using a modified Assay for Transposase Accessible Chromatin with high-throughput sequencing (ATAC-seq) called Omni-ATAC.^59,60^ Briefly, approximately 20 mg of tissue was dounce homogenized in a homogenization buffer. Tissue homogenate was layered over Iodixanol density gradient and spun. Nuclei were extracted post-centrifugation and quantified using a hemocytometer. Approximately 50,000 nuclei were rinsed and added to the Omni-ATAC reaction mix. Transposition reactions were incubated at 37 C for thirty minutes. Reactions were cleaned with spin columns and eluted. Polymerase chain reaction (PCR) was initially performed for five cycles. At this point, a qPCR reaction was performed to determine the additional number of PCR cycles to use. The additional number of PCR cycles is determined by calculating the qPCR cycle at which the fluorescence intensity is equal to one-third the maximum fluorescent intensity of the reaction. Libraries were purified and profiles were measured using Bioanalyzer High-Sensitivity DNA Analysis Kit (Agilent). Libraries that pass visual quality control and concentration checks were frozen at -20 C.

### ATAC-Seq Library Sequencing

Libraries were pooled in two separate groups, 93 samples and 96 samples, and sequenced at Vanderbilt University Medical Center (VUMC VANTAGE (Vanderbilt Technologies for Advanced Genomics)). Libraries were sequenced on NovaSeq 6000 at PE 150 bp. Libraries were pooled and sequenced such that each sample was covered by approximately fifty million sequencing reads.

### ATAC-Seq Data Processing

FASTQ files were processed with fastp (v0.12.5) with parameters “-y -c -g -j fileName.json - h fileName.html -i Read1.fastq.gz -I Read2.fastq.gz -o Read1.fastpProcessed.fastq.gz -O Read2.fastpProcessed.fastq.gz”. FastP processed FASTQ files were aligned to GRCh38 using bwa mem (v0.7.17-r1188) with parameters “-a Read1.fastpProcessed.fastq.gz Read2.fastpProcessed.fastq.gz” and piped into samtools (v.1.9) view with parameters “-S -b -f 2 - > outFile.bam” to generate bam files. Duplicate reads were marked and removed using Picard Tools (v1.141) MarkDuplicates with parameters “INPUT=file.bam OUTPUT=file.PCRDupesRemoved.bam ASSUME_SORTED=true METRICS_FILE=file.dups.txt REMOVE_DUPLICATES=true”. Autosomal reads only were retained using samtools view with parameters “ input.bam -b {1..22} > ${i}.auto.bam”

### Peak Calling and CPM Generation

ATAC-seq peaks were called using Genrich (v0.6.1)^61^, with parameters “./Genrich -t “path/to/bam/file” -j -o outfile -v -E hg38.blacklist.bed -m 10 -g 50 -e “MT, non-canonical chr,etc,”. Reads in peaks were quantified with FeatureCounts (v.1.4.6-p5) with options: ‘featureCounts -T 25 -F SAF -s 0 -a genrichAllPeaks_m10_g50_9.15.21.noBLnarrowpeak.saf -o genrichAllPeaks_m10_g50_9.15.21.noBLnarrowpeak.featureCounts.9.20.21 /home/bwenz/Genrich/bamFiles_5.25.21/*.bam’.

### ATAC-Seq/ChIP-Seq Enrichment

We downloaded the H3K27ac and H3K4me3 ChIP-seq datasets from GEO accession GSE128072. We lifted genomic coordinates of ChIP-seq data from hg19 to hg38 using standard liftover tools. Enrichment of epigenomic datasets was performed by comparing overlap of each ChIP-seq dataset to chromatin accessible regions (ATAC-seq) generated in human liver. Background was calculated by performing 1000 iterations of randomly selecting genomic regions matched to the size and chromosome of true ATAC-seq regions. Overlaps between the ChIP-seq data and the true and shuffled ATAC-seq data were identified using bedtools intersect function. Counts of overlapping ATAC-seq/ChIP-seq regions and sum of overlapping base pairs of these overlaps were calculated for each iteration and the median values were compared to the observed values to perform enrichment calculations. Bedtools (v. 2.30.0) closest function was used to identify the nearest protein-coding gene to ATAC-seq peaks.

### Genotyping and Imputation from Low Coverage Whole Genome Sequencing

Sample genotype was obtained using low-pass whole genome sequencing from Gencove configuration “Human low-pass GRCh37 v2.5 [0.1x-6x])”. Genotypes were filtered to retain only polymorphic sites within our sample population. Polymorphic genotypes were filtered on MAF > 0.05 and variants that deviated from Hardy-Weinberg equilibrium were noted. Remaining genotypes were filtered on genotype posterior probability (GP) > 0.8. Genotypes were phased using Eagle^62^ (v2.4.1).

### Estimating Population Structure

Genotype principal component analysis (PCA) was performed using PLINK (v1.90b6.24 (16 April 2021)).^63^

### Mapping *cis*-Chromatin Accessibility Quantitative Trait Loci (*cis*-caQTLs)

Cis-caQTLs were identified using RASQUAL (v1.1). Covariates included in the model were the first 16 PEER factors (hidden latent factors)^64^, first 3 genotype principal components (PCs), sample sex, and sample ATAC-seq sequencing batch. All variants within 10 kilobases from the start and end of the ATAC-seq peak were tested for association with chromatin accessibility. Samples used in the African ancestry caQTL mapping analysis were chosen based on a combination of genotype principal component analysis (PCA), ancestries reported from Gencove, and results from the software ADMIXTURE (v1.3.0)^65^. Genotype PCA was generated using PLINK (v1.90b6.24 (16 April 2021)) and annotated with Gencove reported ancestries, suggesting the presence of a group of samples with a greater proportion of African ancestry (**Supplementary Figure 45**). Based on the results of cross-validation across K values to minimize error, ADMIXTURE was run on plink genotype files with K=3 ancestral populations. From these results, we chose the 26 samples with evidence suggesting the proportion of the African ancestry population was greater than 0.5 (**Supplementary Figure 46**). We further validated these ancestry groupings by combining liver sample genotypes with 1000g data and plotting genotype PCA (**Supplementary Figure 47**).

### Integrative Analyses of Liver *cis*-caQTLs with Other Functional Datasets

We downloaded the “encRegTfbsClusteredWithCells.hg38.bed.gz” file from https://hgdownload.soe.ucsc.edu/goldenPath/hg38/encRegTfbsClustered/. Using this file, we extracted the binding sites of 119 transcription factors that were obtained in HepG2 cell line. Peaks with *cis*-caQTLs were overlapped with 119 different transcription factors’ binding sites in HepG2 cells using bedtools2 (v2.30.0) intersect function. Enrichment of overlap was calculated relative to 1,000 sets of randomly chosen peaks, matched on size, GC, and distance to TSS. One-sided Fisher’s exact test was used to assess the significance of the enrichment.

### Colocalization Analyses

Colocalization was performed using coloc (v5.2.3). Colocalizations were performed between GWAS/caQTLs for caQTL peaks within 1 Mb of the GWAS sentinel lead variant, GWAS/eQTL colocalizations were performed for eGenes with TSS within 1 Mb of the GWAS sentinel lead variant, and caQTL/eQTL colocalizations were performed for caQTLs within 1 Mb of the eGene transcription start site (TSS). Summary statistics for all variants within a 1 Mb cis-window around significant caQTL peaks from the discovery analysis were generated, and colocalization was performed using matched variants from the corresponding studies. Colocalization results were filtered to retain only colocalization loci that were sufficiently powered, requiring PP3+PP4 > 0.8. For those events that surpass this threshold, we assessed whether the colocalization is considered high confidence, PP4 >= 0.8, or medium confidence, 0.5 < PP4 < 0.8. GTEx v8 data were downloaded from https://www.gtexportal.org/home/downloads/adult-gtex/bulk_tissue_expression.GTExv8. GTEx genes for colocalization were defined based on GTF file ftp://ftp.ebi.ac.uk/pub/databases/gencode/Gencode_human/release_26/gencode.v26.annotation.gtf.gz. Source of GWAS data sets is in **Table S9**.

### Unresolved GWAS Signal Analyses

We annotated unresolved GWAS signals using ANNOVAR (version released 7 June 2020)^45^ to identify exonic variants.

### Power Calculation

R package winnerscurse (v0.1.1) and method FDR_IQT was used to adjust effect sizes for winner’s curse. Non-centrality parameter was calculated by dividing the effect size by the standard error. Power was calulcated with R function Power.Chisq() from package OptSig (v. 2.2). The power groups were 0 to 0.5 for low, 0.5 to 0.95 for medium, and 0.95 to 1 for high. Enrichment of power categories within each colocalization group was assessed using a chi-square test with Pearson-adjusted residuals.

### TF Footprinting Analysis

Transcription factor footprinting analysis was performed using all reads from all samples combined. Footprinting analysis was performed using TOBIAS (v.0.15.1)^66^ using the BINDetect function with default parameters. TF motifs for BINDetect came from the JASPAR CORE 2022 vertebrate non-redundant dataset (https://jaspar2022.genereg.net/downloads/).

## Data Availability

FASTQ sequencing files for the liver ATAC-seq samples used in this study have been deposited in the Gene Expression Omnibus (GEO; accession GSE277774).

## Authors’ contributions

C.D.B., B.F.V., and B.M.W. designed the study. B.M.W. generated the data, performed computational analyses, and prepared the figures and tables. B.F.V., M.F.D. and S.R. assisted with data analysis. K.T.C., D.X., K.M.O., A.S., and D.J.R. contributed to sample acquisition. B.F.V. and B.M.W. drafted and revised the manuscript. All authors read and approved the final manuscript.

## Data and Code Availability

FASTQ sequencing files for the liver ATAC-seq samples used in this study have been deposited in the Gene Expression Omnibus (GEO; accession GSE277774). Code will be made available upon acceptance of the manuscript.

## Acknowledgements

B.F.V. is grateful for support for the work from the NIH/NIDDK (DK126194).

