## Supplementary Figures for "Expanded Chromatin Accessibility Mapping Explains Genetic Variation Associated with Complex Traits in Liver"

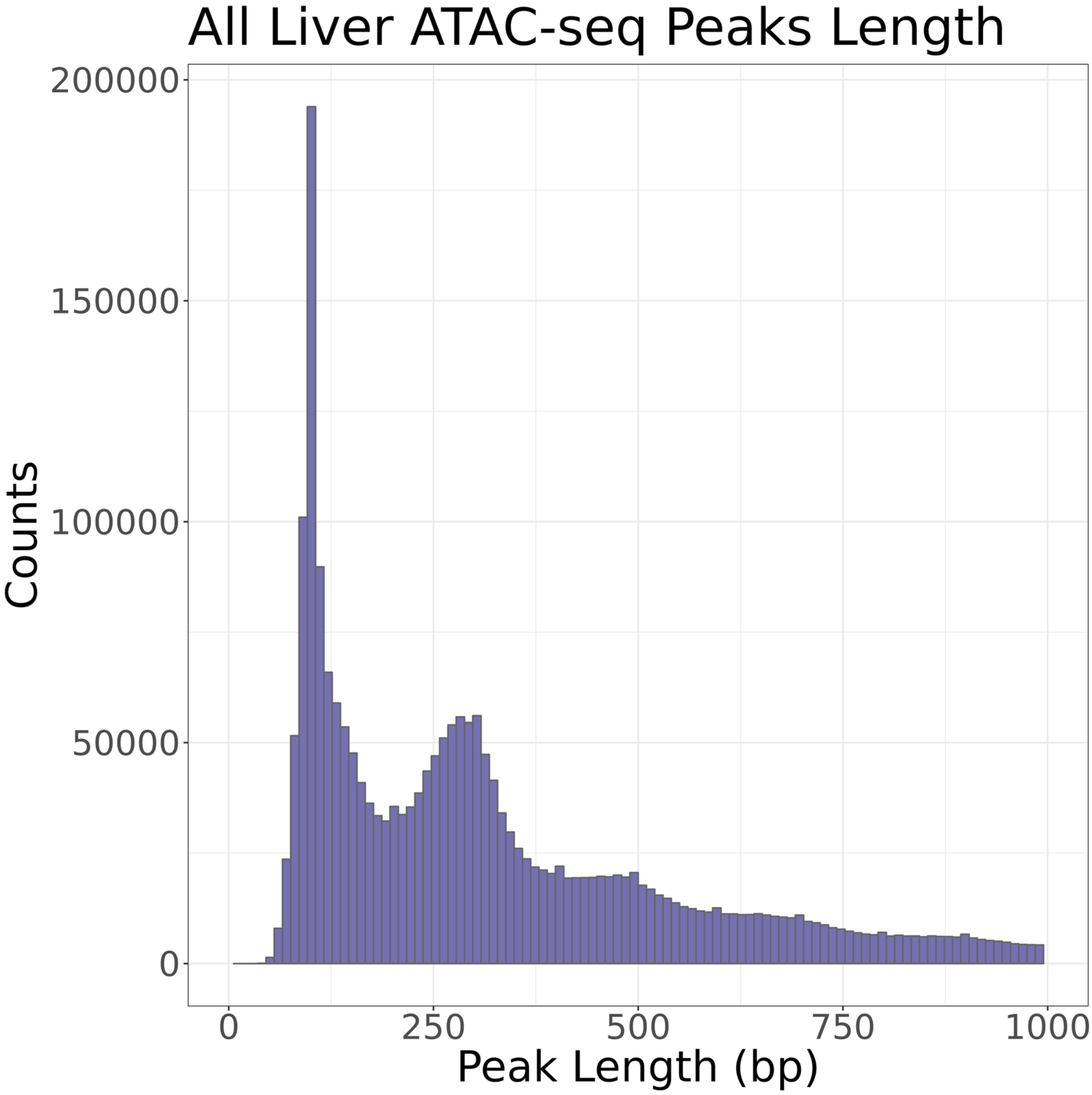


Supplementary Figure 1: 2,518,633 ATAC-seq peaks were identified. Median peak length was 292 bp.

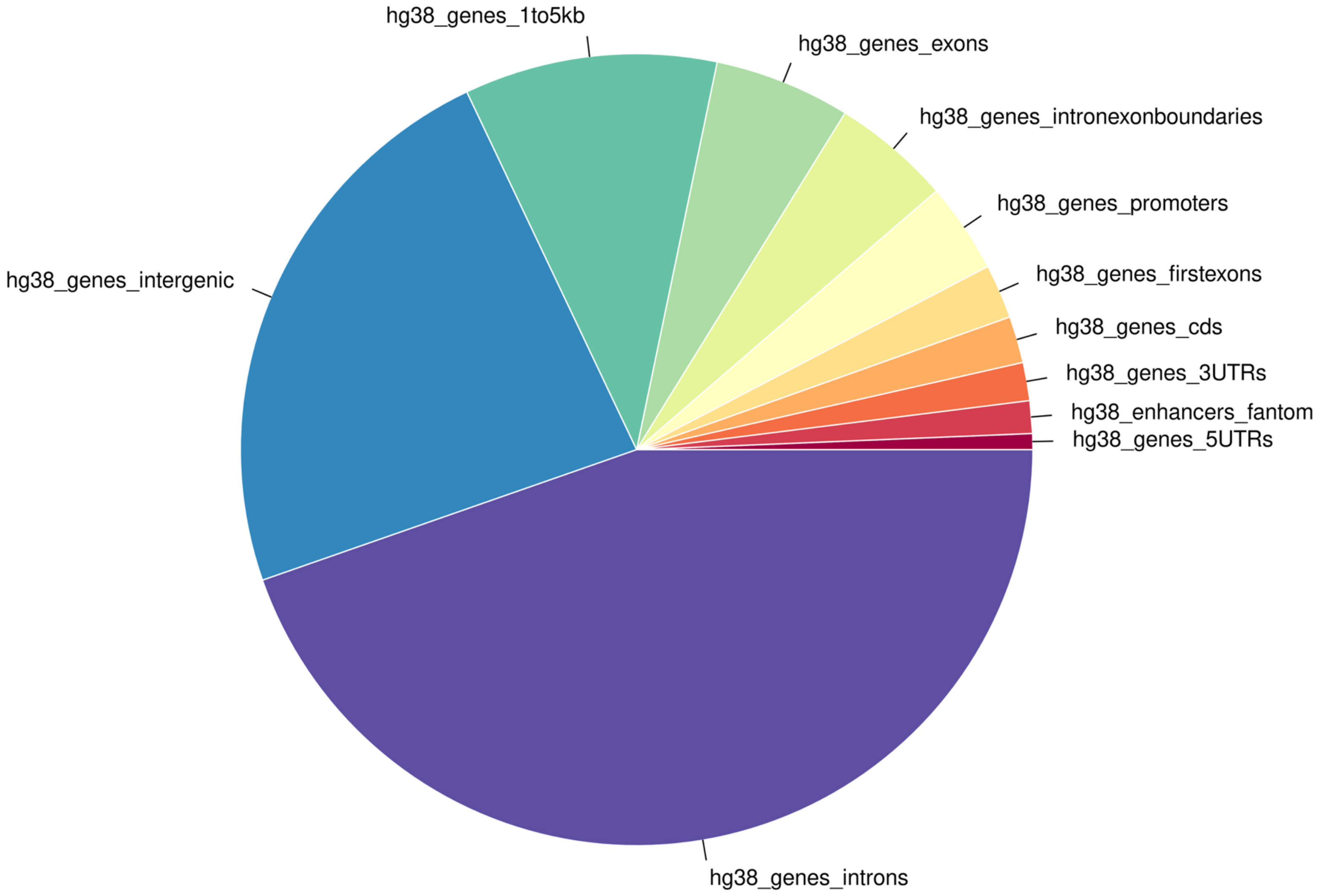


Supplementary Figure 2: Liver ATAC-seq peaks were annotated to genomic regions using annotatr.

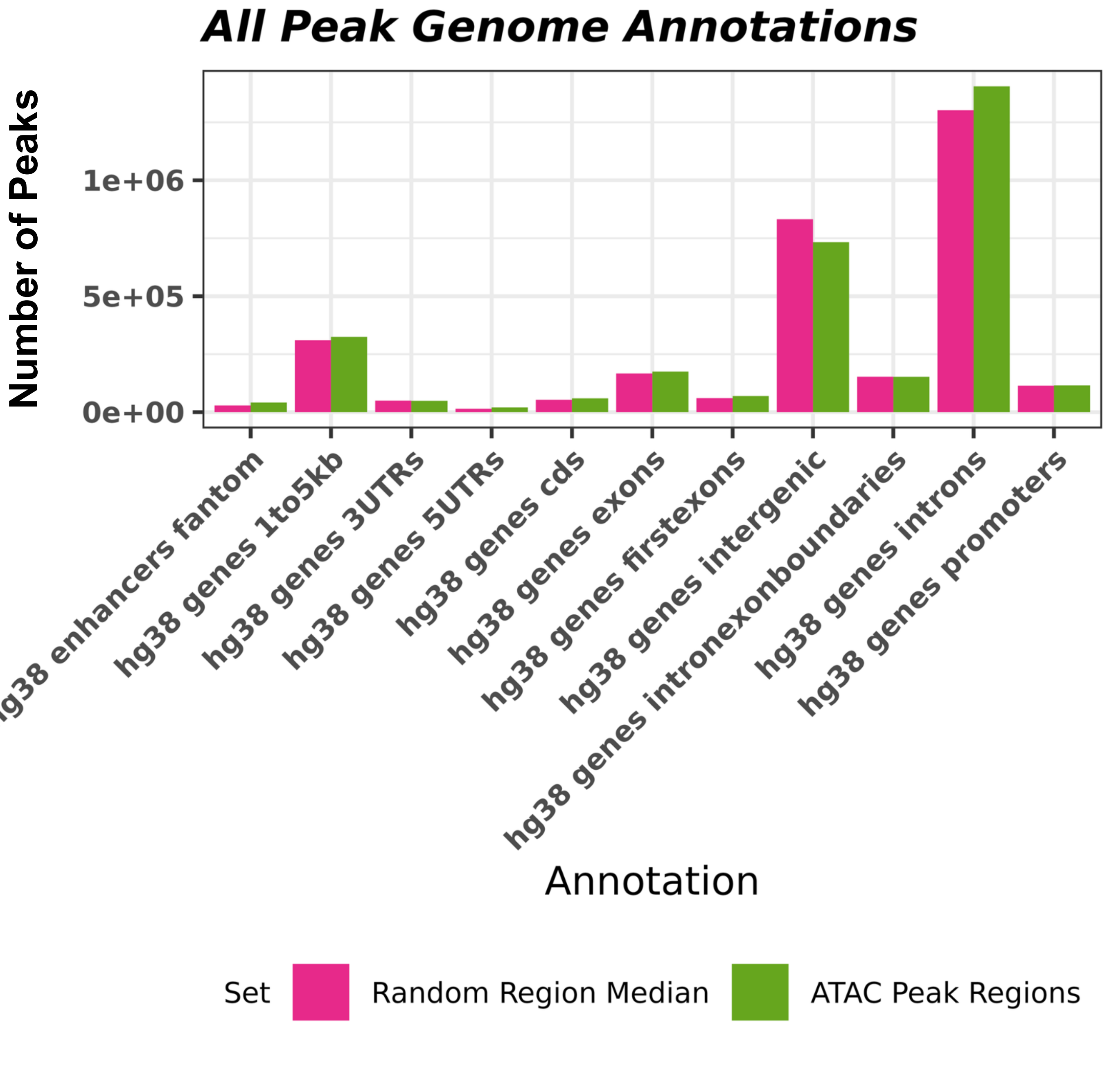


Supplementary Figure 3: Peaks were enriched for a variety of genomic annotation categories, most significantly in FANTOM enhancer regions and gene 5' UTRs. Peaks were significantly depleted in intergenic regions and gene 3' UTRs.

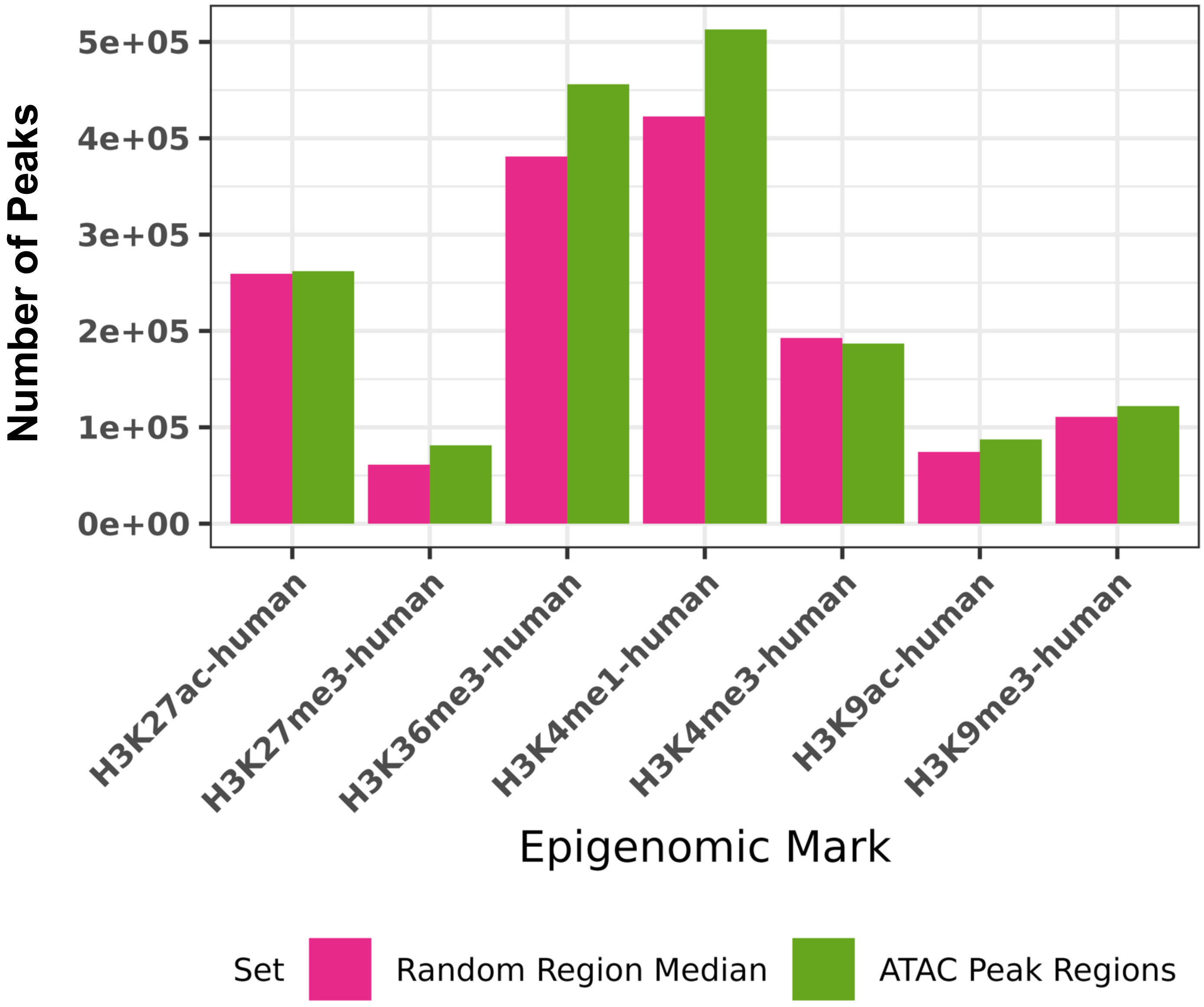


Supplementary Figure 4: Peaks were significantly enriched for all histone marks from the NIH Roadmap Epigenomics Consortium ,other than H3K4me3, which was significantly depleted.

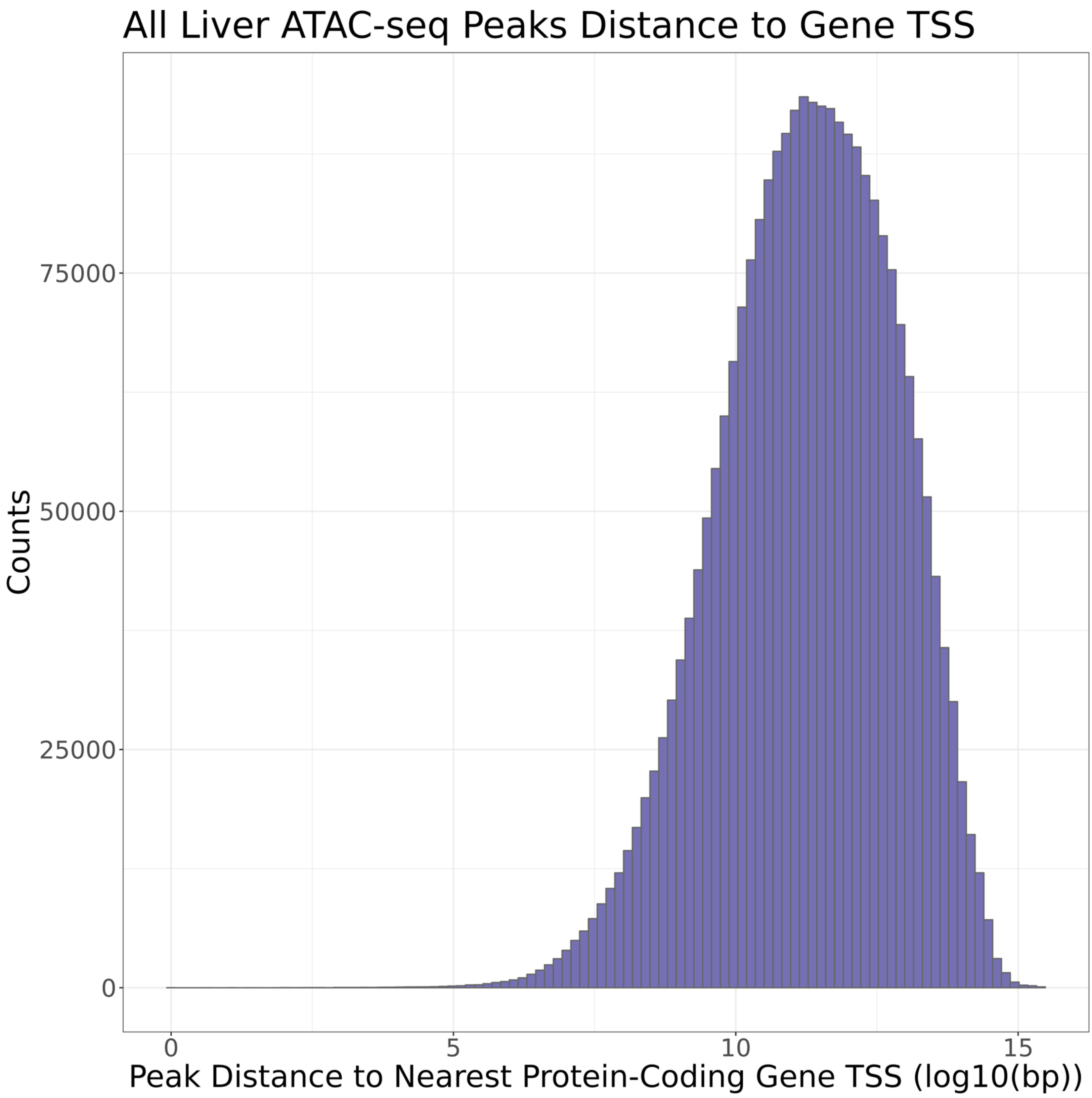


Supplementary Figure 5: The distance between liver ATAC-seq peaks and the nearest protein-coding gene TSS was calculated. The median distance was 82,184 base pairs and 17,370 peaks directly overlapped the nearest TSS.

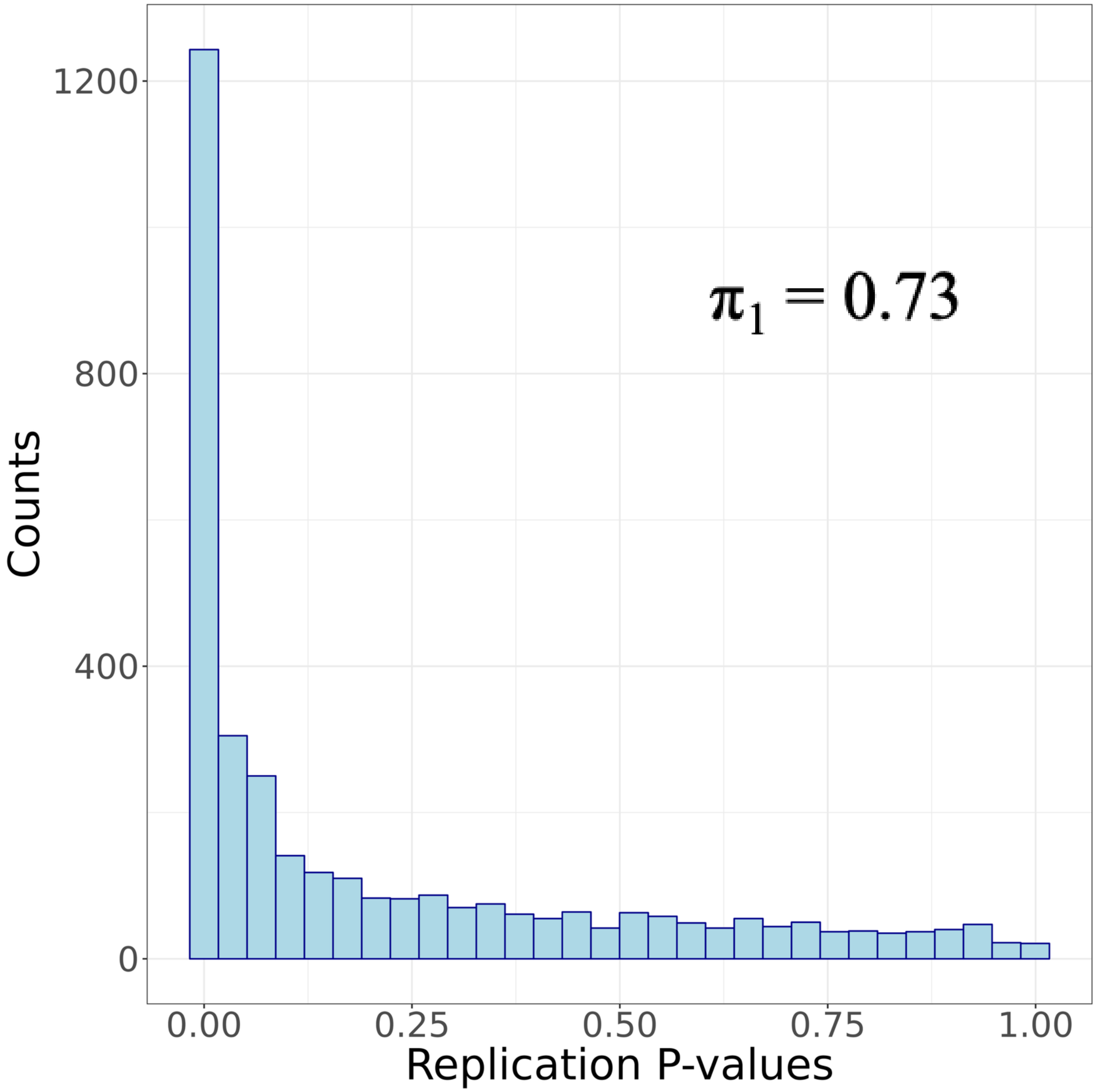


Supplementary Figure 6: Replication of caQTLs identified in our study compared to a previously published liver caQTL study. For caQTL peaks in our study that overlapped peaks in the external study, we matched lead variants in our study to the external study, keeping only the most significant association. Plotted are the p-values from the replication data set.

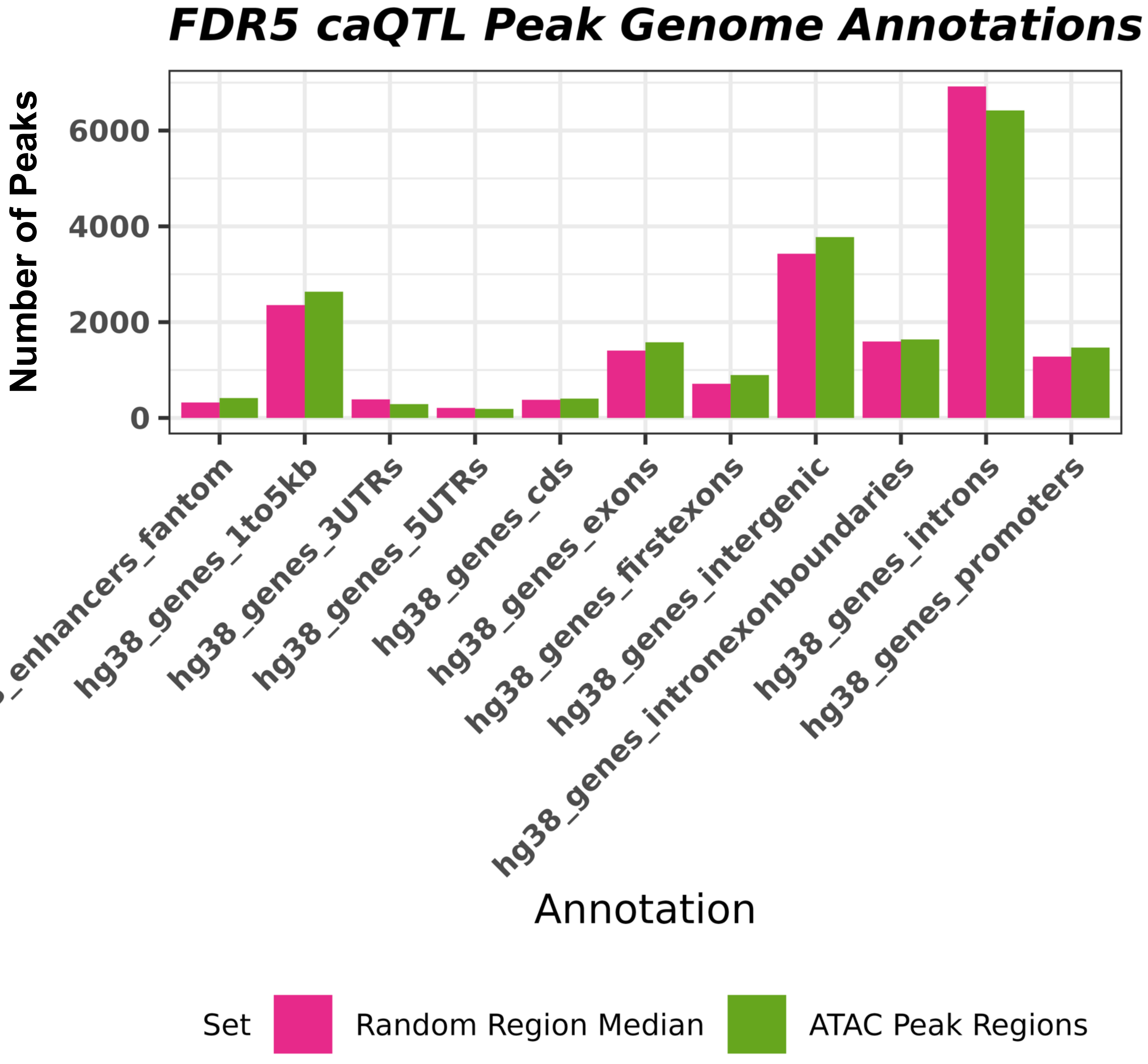


Supplementary Figure 7: caQTL peaks were enriched for a variety of genomic annotation categories, most significantly in FANTOM enhancer regions and gene 5' UTRs. Peaks were significantly depleted in intergenic regions and gene 3' UTRs.

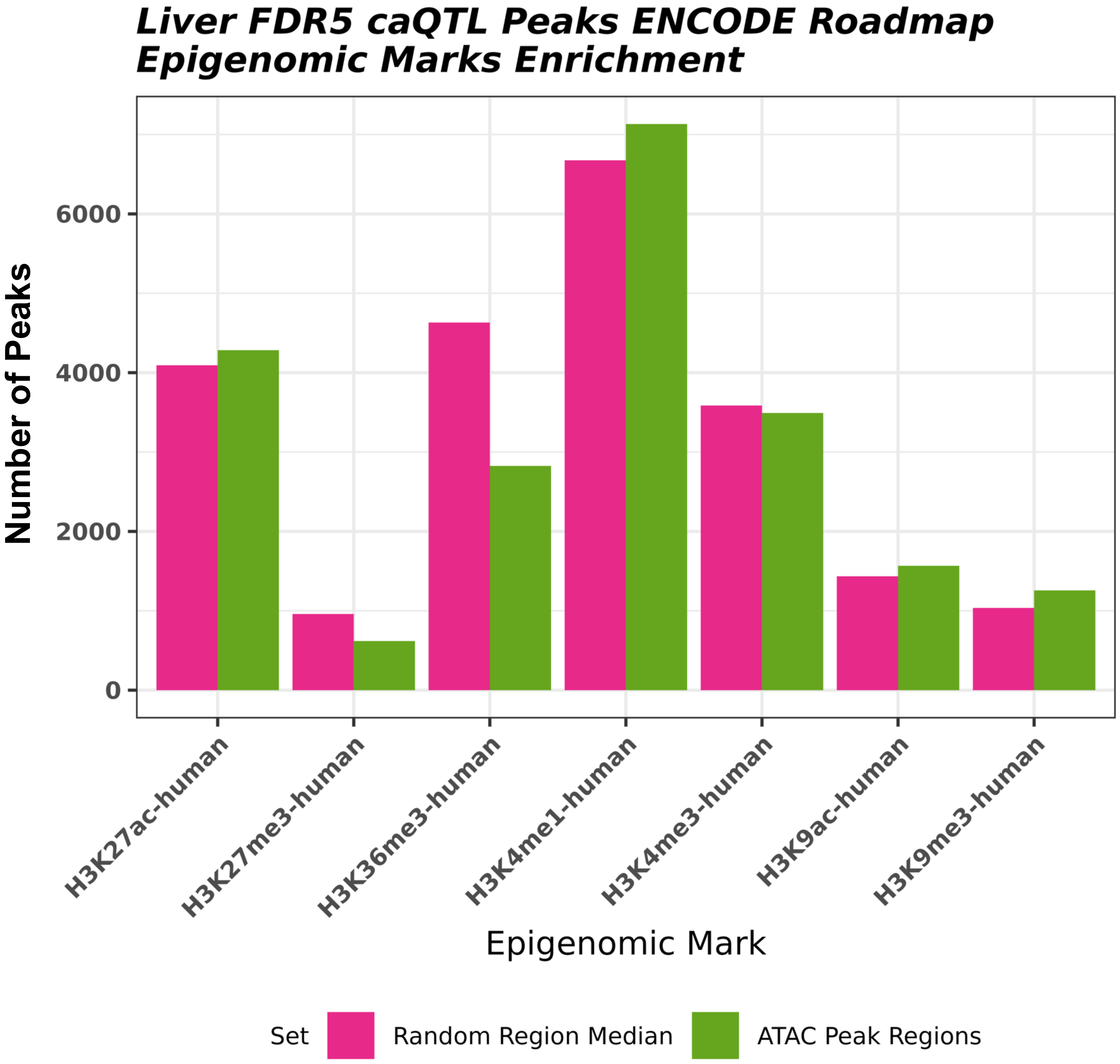


Supplementary Figure 8: caQTL peaks were significantly enriched for H3K9me3, H3K9ac, and H3K4me1 and significantly depleted for H3K27me3 and H3K36me3.

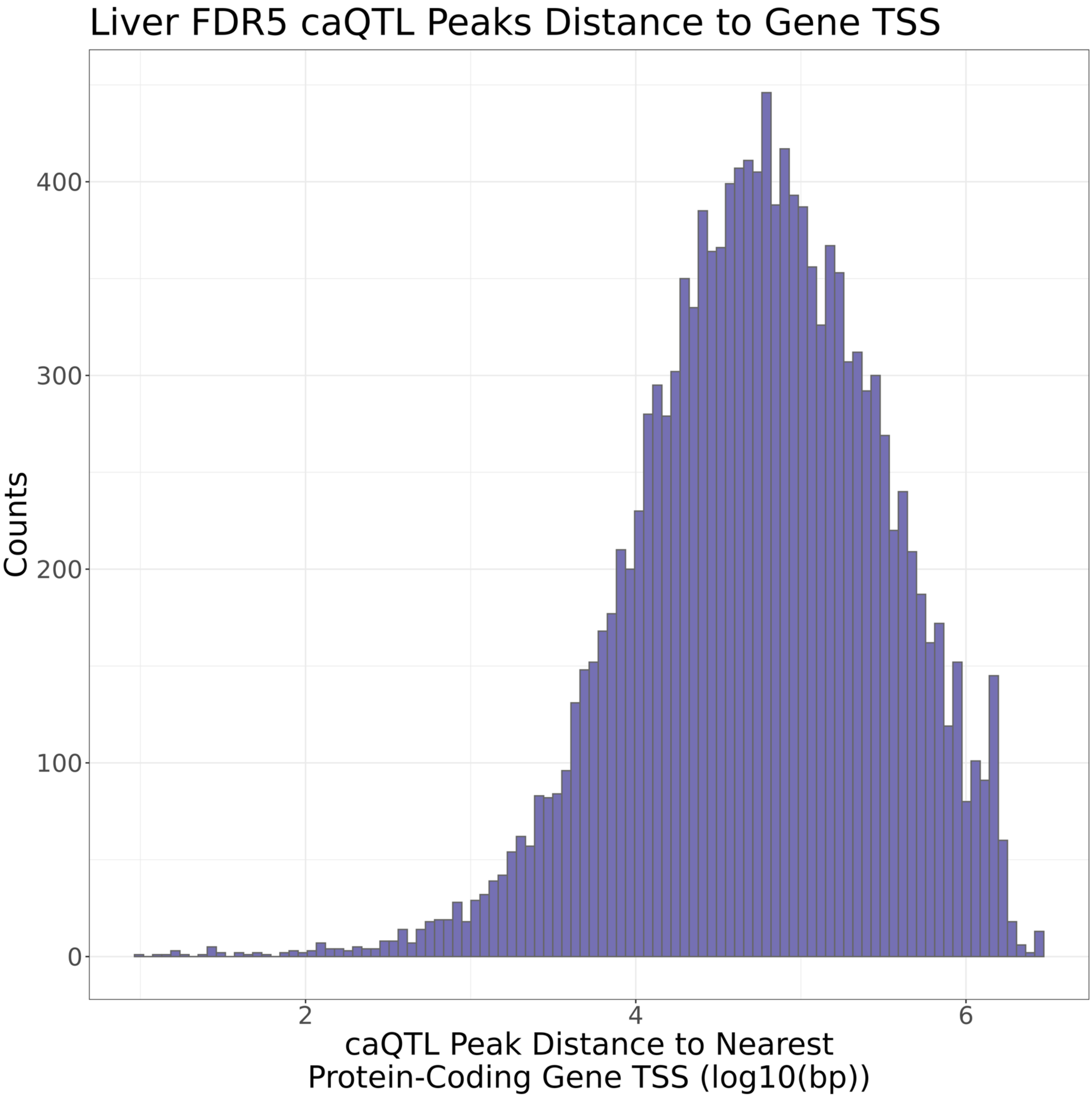


Supplementary Figure 9: The distance between liver FDR5 caQTL peaks and the nearest protein-coding gene TSS was calculated. The median distance was 55,361 base pairs and 387 peaks directly overlapped the nearest TSS.

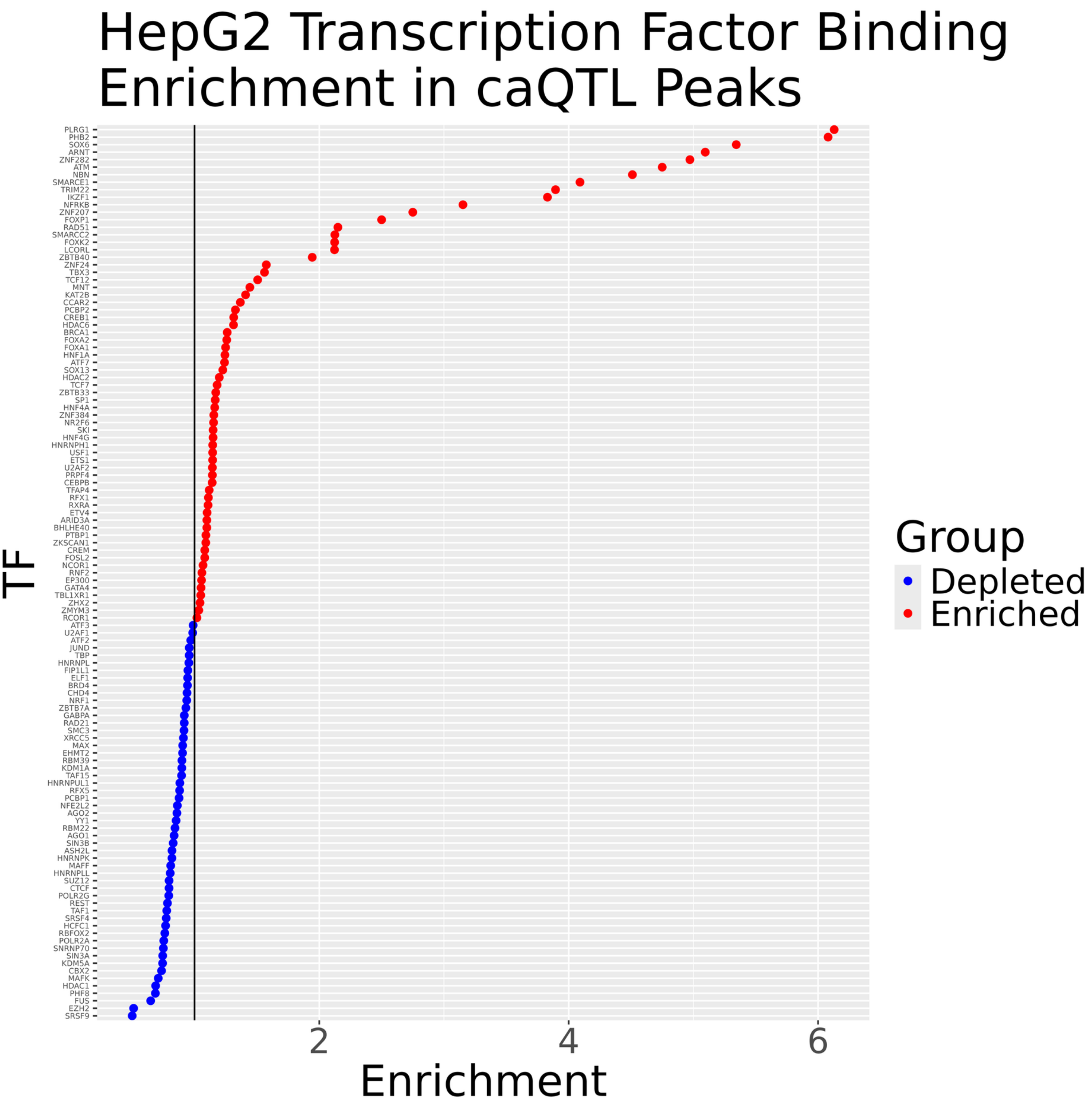


Supplementary Figure 10: Liver caQTL peaks were intersected with ENCODE ChIP-seq data from HepG2 cell lines. Shown are transcription factors that were enriched and depleted compared to matched random controls.

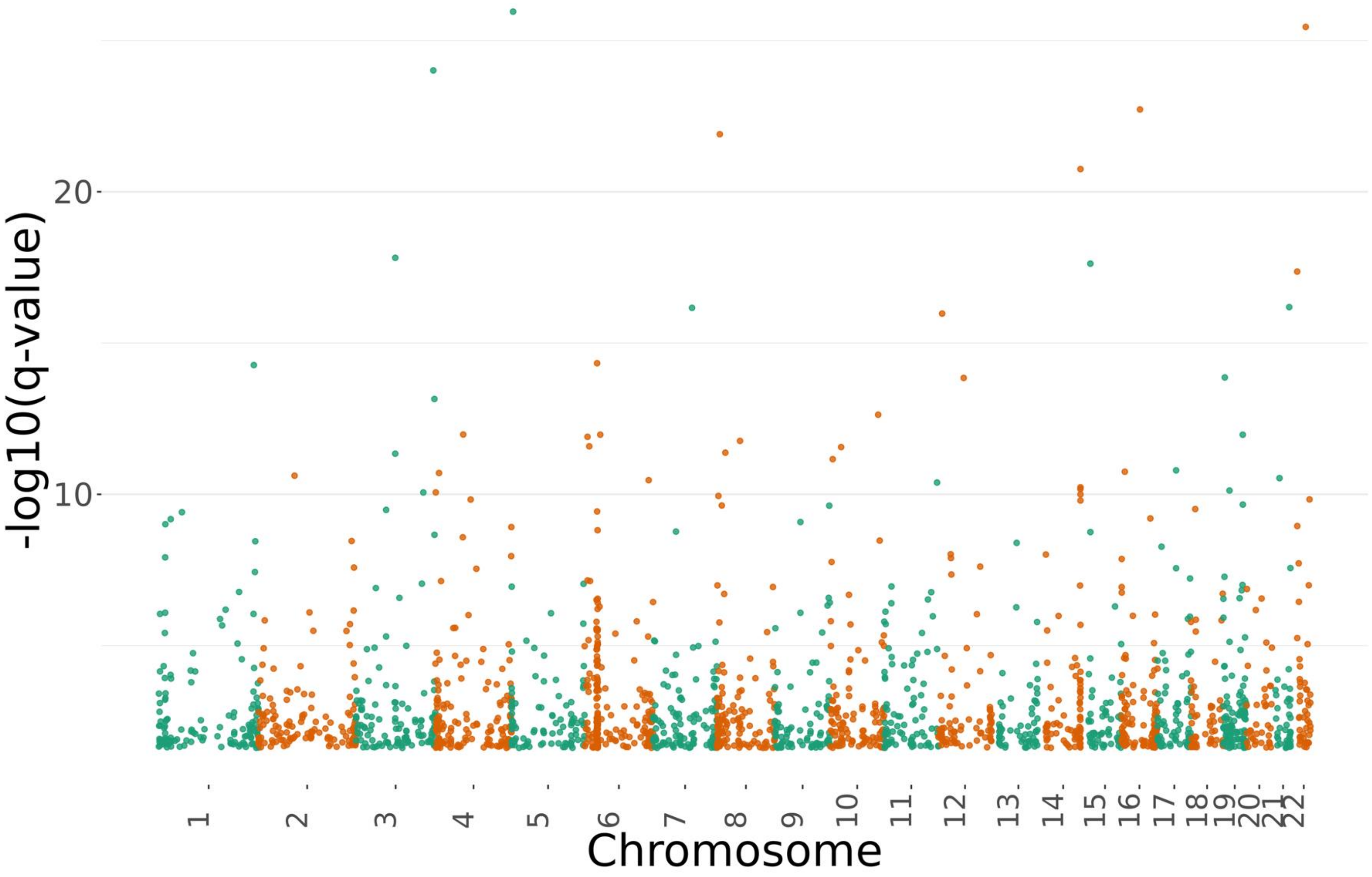


Supplementary Figure 11: We identified 26 samples that had predominant evidence of African ancestry and mapped 2,088 FDR5 caQTLs in this subset.

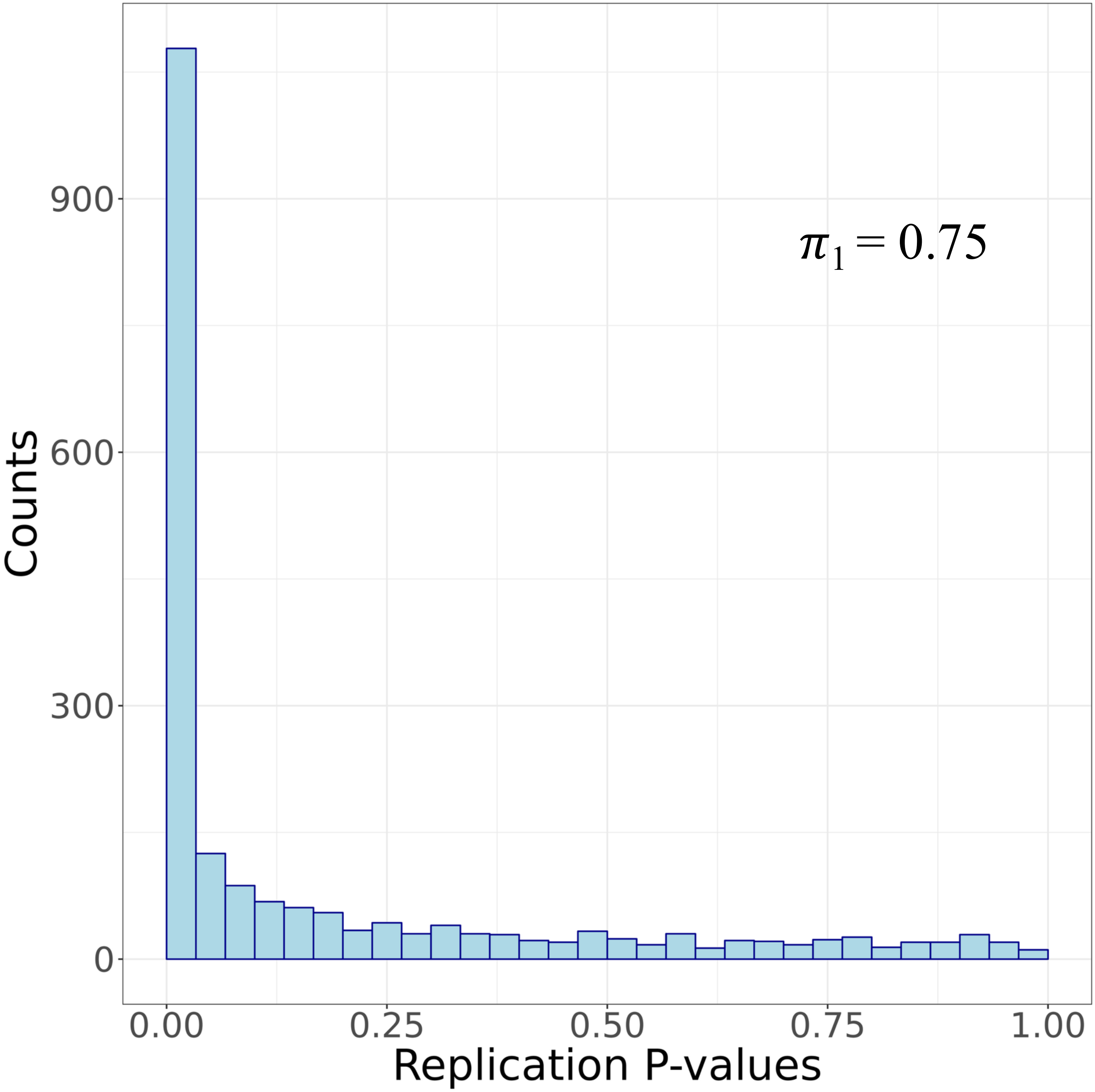


Supplementary Figure 12: FDR5 caQTL peak-lead variant pairs identified in the African ancestry analysis were extracted from the full sample set results and replication was estimated. Full sample set p-values are plotted.

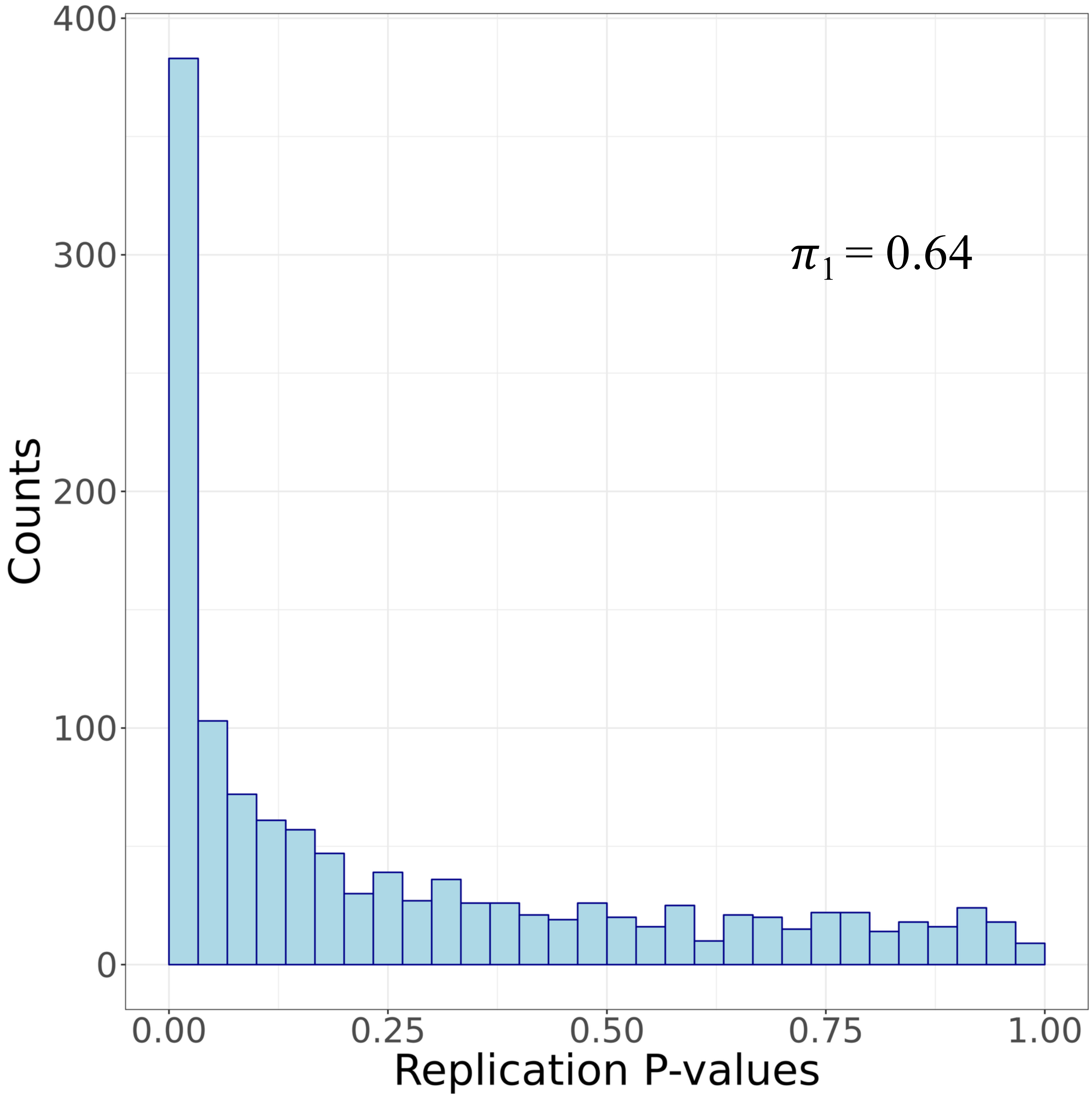


Supplementary Figure 13: FDR5 caQTL peak-lead variant pairs specific to the African ancestry analysis were extracted from the full sample set results and replication was estimated. Full sample set p-values are plotted.

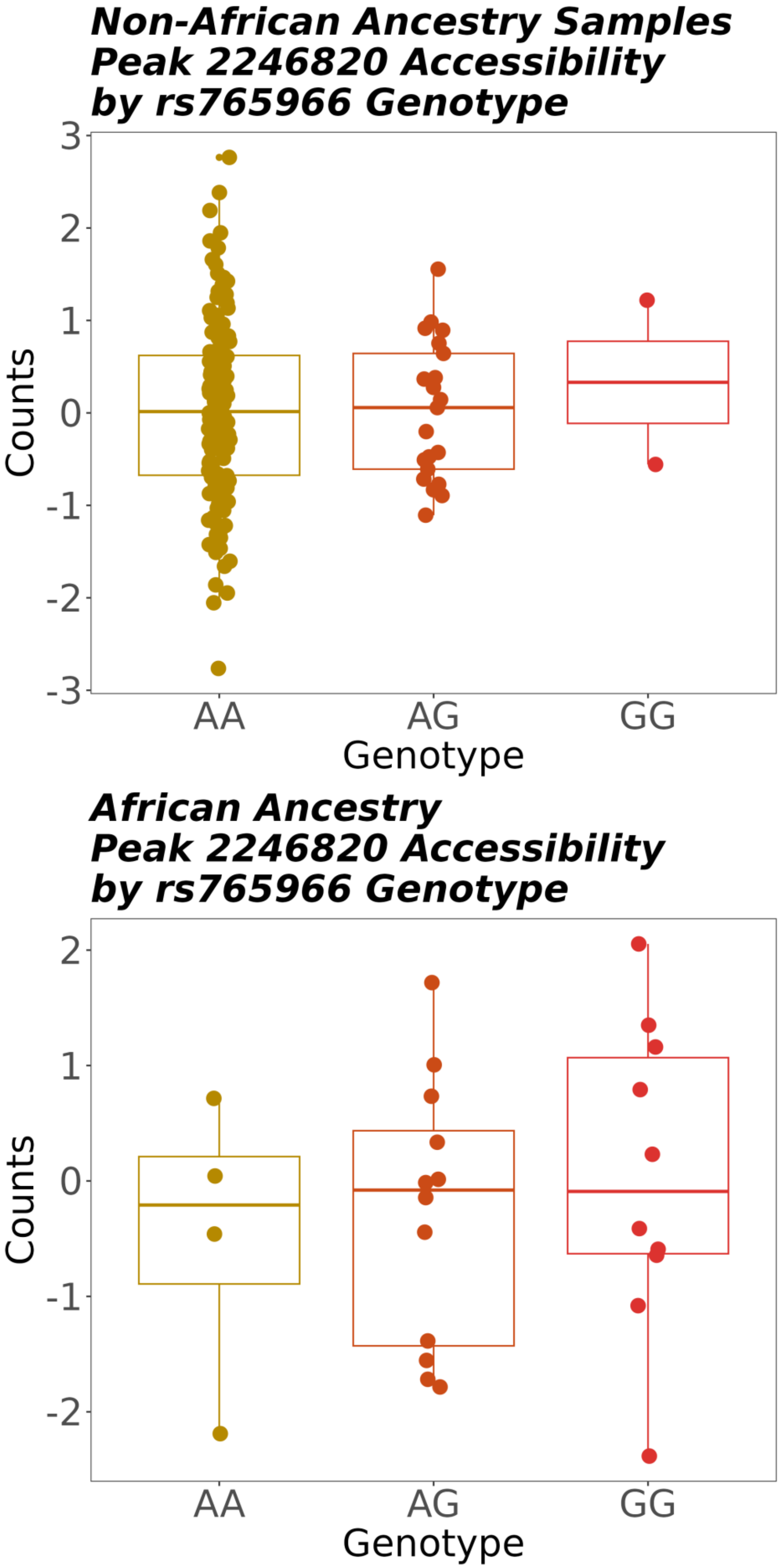


Supplementary Figure 14: Sample read counts for a caQTL peak specific to the African ancestry caQTL mapping analysis were plotted in the individuals with African ancestry and those not included in the African ancestry analysis to highlight differences in allele frequencies between these two groups.

### EUR Only GLGC Lipids Colocalizations

| GWAS Trait | Unique caQTL Peaks Colocalized | Unique caQTL Lead Variants Colocalized | Unique Sentinel GWAS Signals Colocalized |
|---|---|---|---|
| HDL | 52 | 43 | 39 |
| LDL | 44 | 44 | 44 |
| logTG | 51 | 47 | 28 |
| nonHDL | 38 | 38 | 41 |
| TC | 58 | 50 | 48 |

| GWAS Trait | Unique eGene Colocalizations | Unique eQTL Lead Variants | Unique Sentinel GWAS Signals |
|---|---|---|---|
| HDL | 78 | 76 | 67 |
| LDL | 94 | 90 | 81 |
| logTG | 80 | 72 | 61 |
| nonHDL | 90 | 85 | 70 |
| TC | 105 | 100 | 89 |

### Trans-ancestry GLGC Lipids Colocalizations

| GWAS Trait | Unique caQTL Peaks Colocalized | Unique caQTL Lead Variants Colocalized | Unique Sentinel GWAS Signals Colocalized |
|---|---|---|---|
| HDL | 79 | 69 | 50 |
| LDL | 59 | 59 | 57 |
| logTG | 63 | 60 | 50 |
| nonHDL | 40 | 40 | 43 |
| TC | 84 | 74 | 54 |

| GWAS Trait | Unique eGene Colocalizations | Unique eQTL Lead Variants | Unique Sentinel GWAS Signals |
|---|---|---|---|
| HDL | 94 | 93 | 88 |
| LDL | 100 | 95 | 89 |
| logTG | 92 | 84 | 74 |
| nonHDL | 97 | 92 | 79 |
| TC | 99 | 97 | 94 |

Supplementary Figure 15: Across five blood lipid traits that primarily involve the liver, we find a range of colocalizations for both caQTLs and eQTLs in both European and trans-ancestry sample cohorts. Through colocalization analyses, eQTLs explain 1.66- to 1.8-fold more GWAS signals compared to caQTLs.

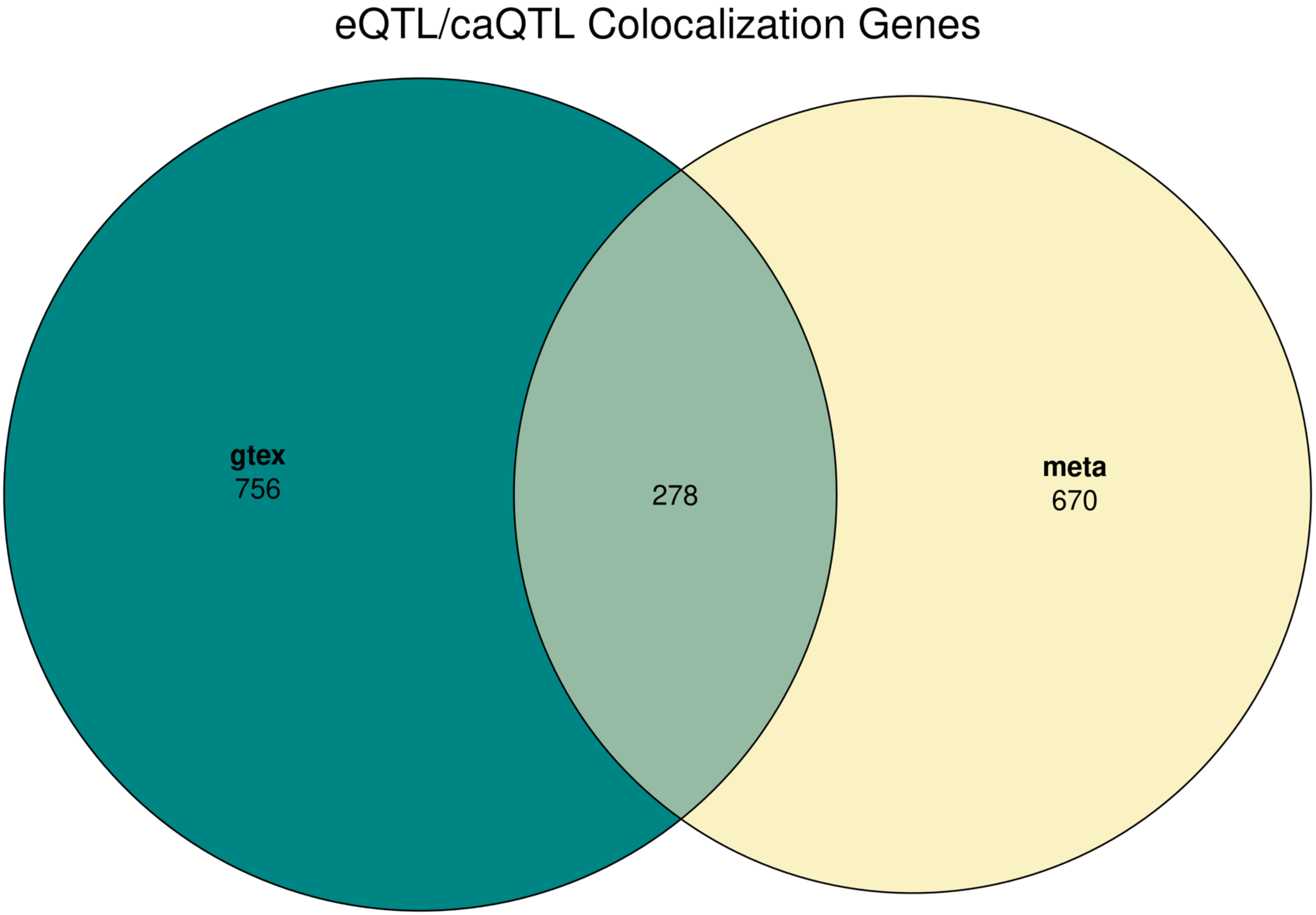


Supplementary Figure 16: Colocalizations were performed between liver caQTLs and liver eQTLs from GTEx and a meta-analysis. Sharing of genes that colocalized is shown (Odds Ratio = 10.4, p < 2.2e-16 , 95% CI: [8.85, 12.2]).

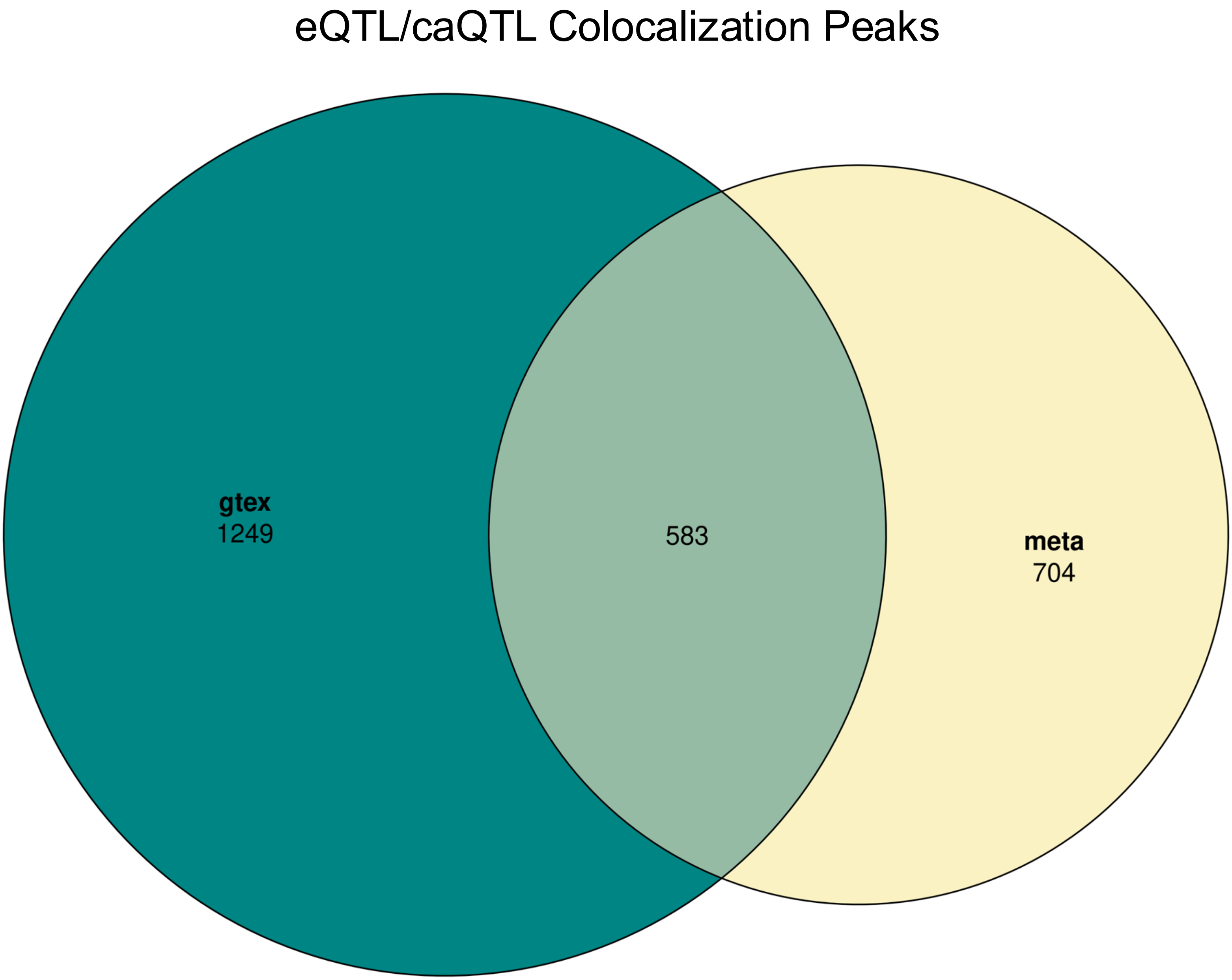


Supplementary Figure 17: Colocalizations were performed between liver caQTLs and liver eQTLs from GTEx and a meta-analysis. Sharing of caQTL peaks that colocalized is shown (Odds Ratio = 7.7, p < 2.2e-16 , 95% CI: [6.74, 8.68]).

Effect Size Correlation Between Colocalizing GTEx/Meta-analysis eQTLs and Liver caQTLs Sharing the Same Lead Variant

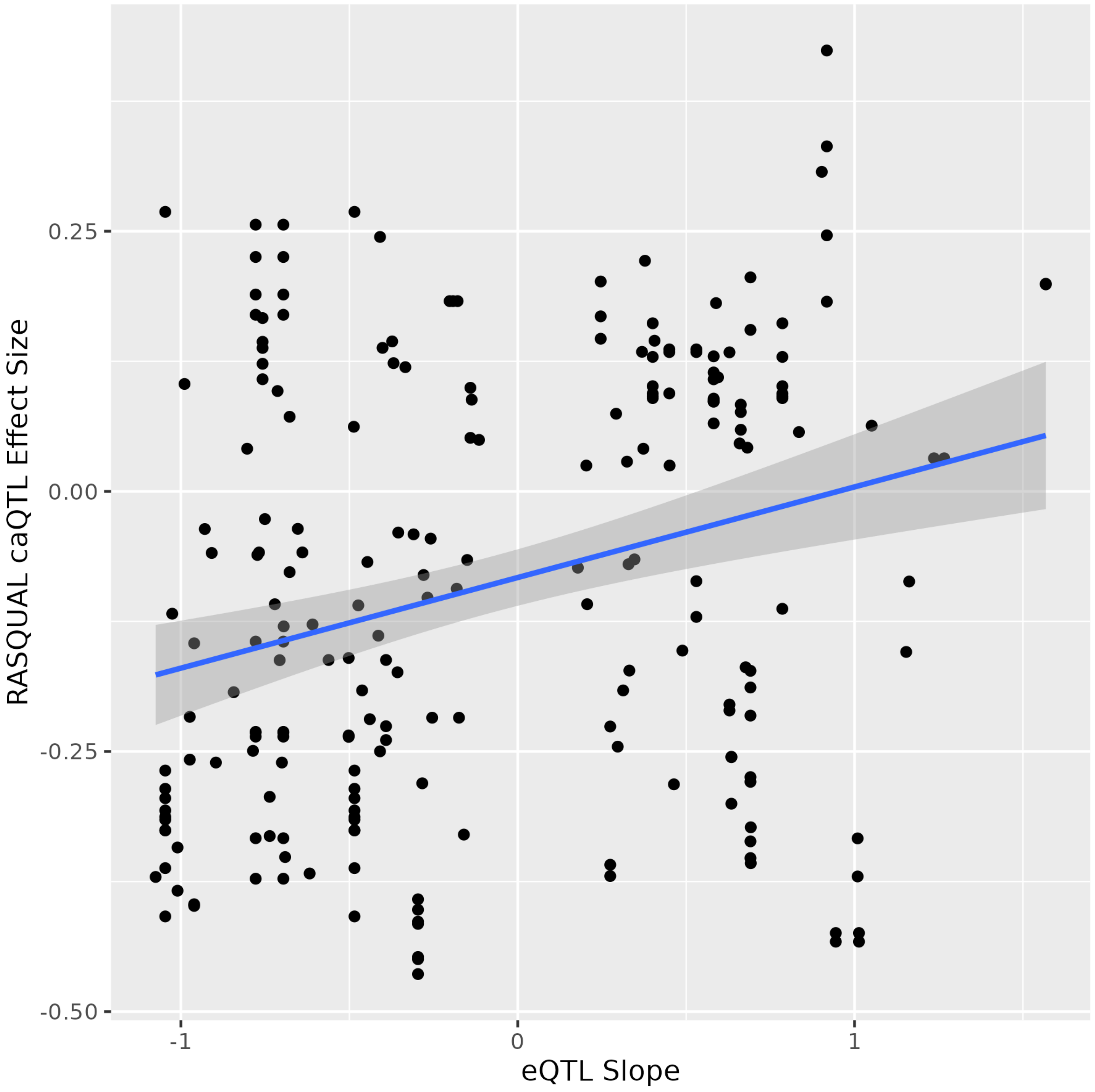


Supplementary Figure 18: Colocalizations were performed between liver caQTLs and liver eQTLs from GTEx and a meta-analysis. For pairs that colocalized and shared lead variants, effect sizes are plotted (r = 0.28, p = 2.459e-05, 95% CI: [0.15, 0.40]).

### Trans-Ancestry GLGC Lipids – Meta-analysis Liver eQTL Colocalizations

| GWAS Trait | Unique eGene Colocalizations | Unique eQTL Lead Variants | Unique Sentinel GWAS Signals |
|---|---|---|---|
| HDL | 145 | 143 | 151 |
| LDL | 153 | 144 | 139 |
| logTG | 119 | 115 | 129 |
| nonHDL | 146 | 137 | 120 |
| TC | 197 | 185 | 174 |

Supplementary Figure 19: Across 5 blood lipids traits that primarily involve the liver, we find a range of colocalizations between GWAS signals identified in the trans-ancestry sample cohort and eQTLs from a liver expression meta-analysis.

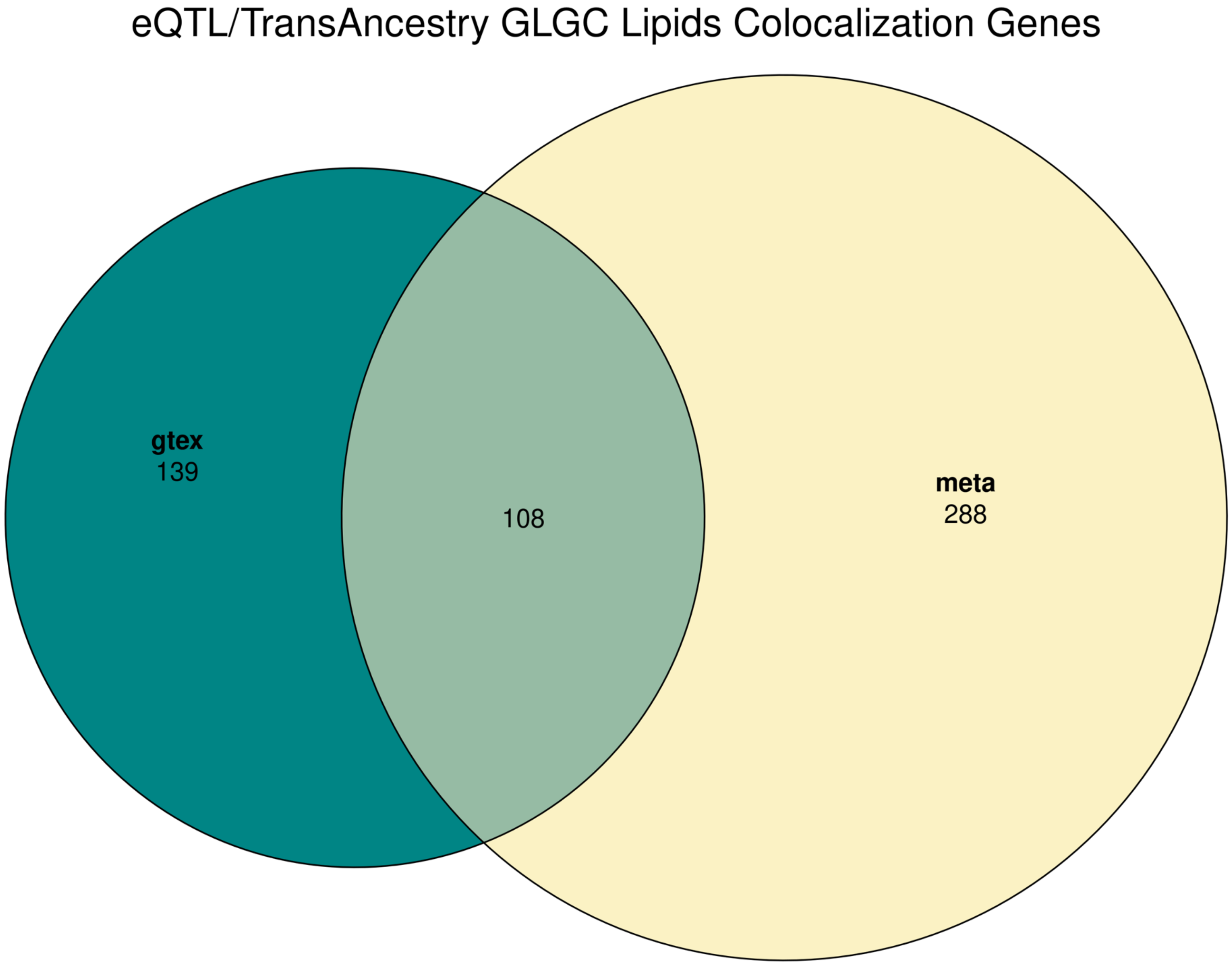


Supplementary Figure 20: Colocalizations were performed between liver caQTLs and liver eQTLs from GTEx and a meta-analysis. Sharing of genes that colocalized is shown (Odds Ratio = 54.2, $p < 2.2e-16$ , 95% CI: [40.71, 72.36]).

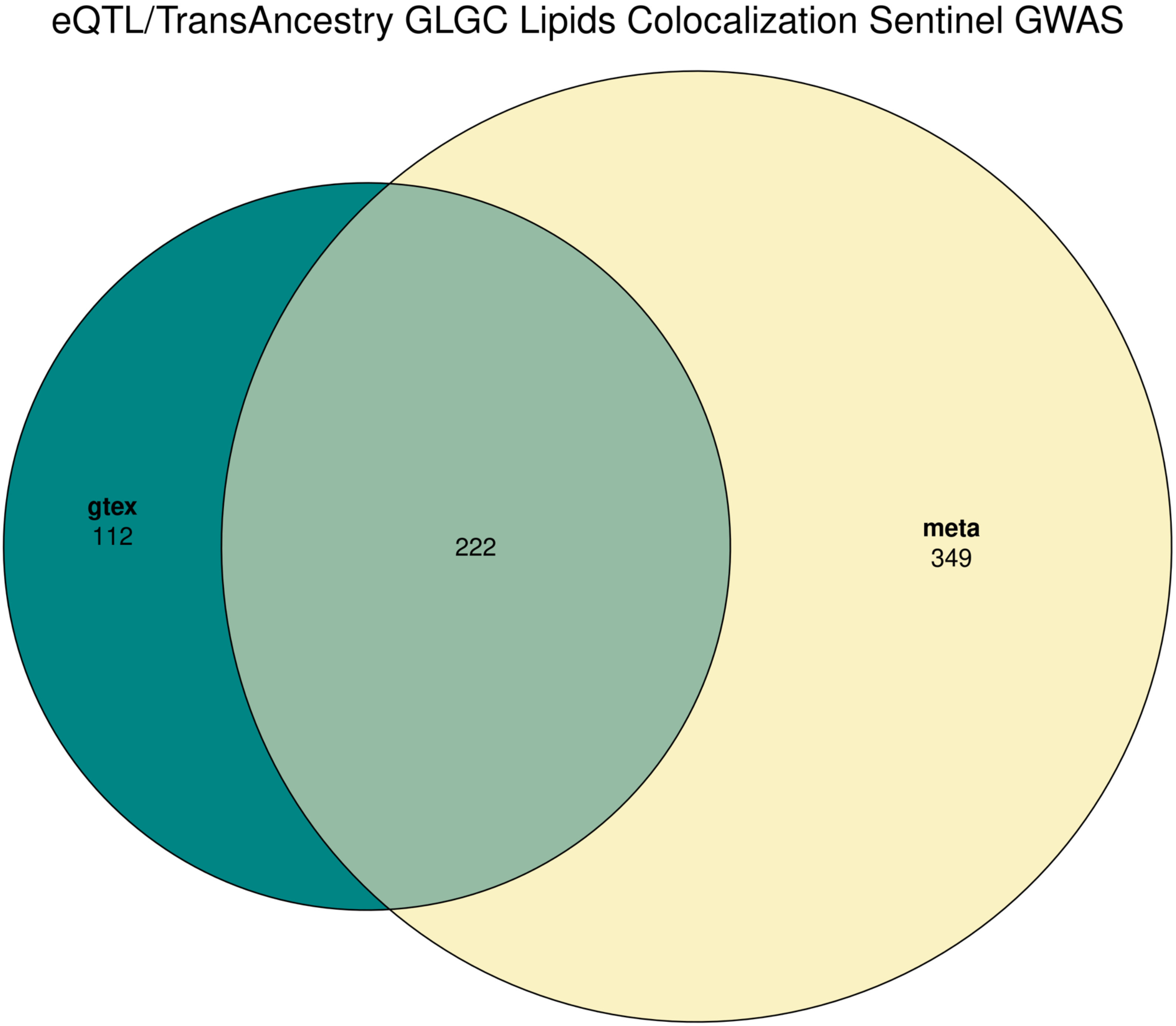


Supplementary Figure 21: Colocalizations were performed between liver caQTLs and liver eQTLs from GTEx and a meta-analysis. Sharing of GWAS sentinels that colocalized is shown (Odds Ratio = 9.73, p < 2.2e-16 , 95% CI: [7.5, 12.7]).

| GWAS Trait | Number of Three-Way Colocalizations | Unique Peaks | Unique caQTL Lead Variants | Unique GWAS Signals | Unique eGenes |
|---|---|---|---|---|---|
| HDL | 64 | 29 | 27 | 19 | 24 |
| LDL | 51 | 23 | 23 | 22 | 27 |
| logTG | 103 | 29 | 26 | 16 | 29 |
| nonHDL | 29 | 12 | 12 | 13 | 19 |
| TC | 51 | 30 | 28 | 18 | 24 |

Supplementary Figure 22: Across 5 blood lipids traits that primarily involve the liver, we find a range of colocalizations between caQTLs, GTEx and/or meta-analyzed eQTLs, and trans-ancestry blood lipids GWAS signals.

A

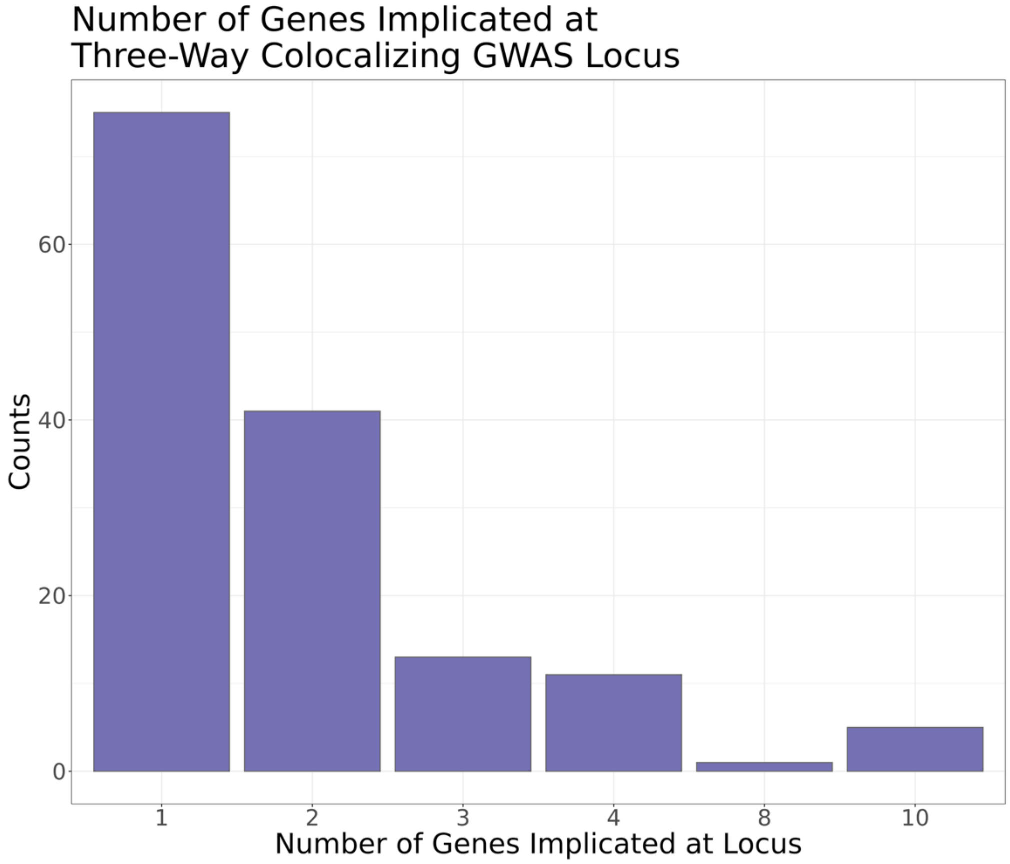


B

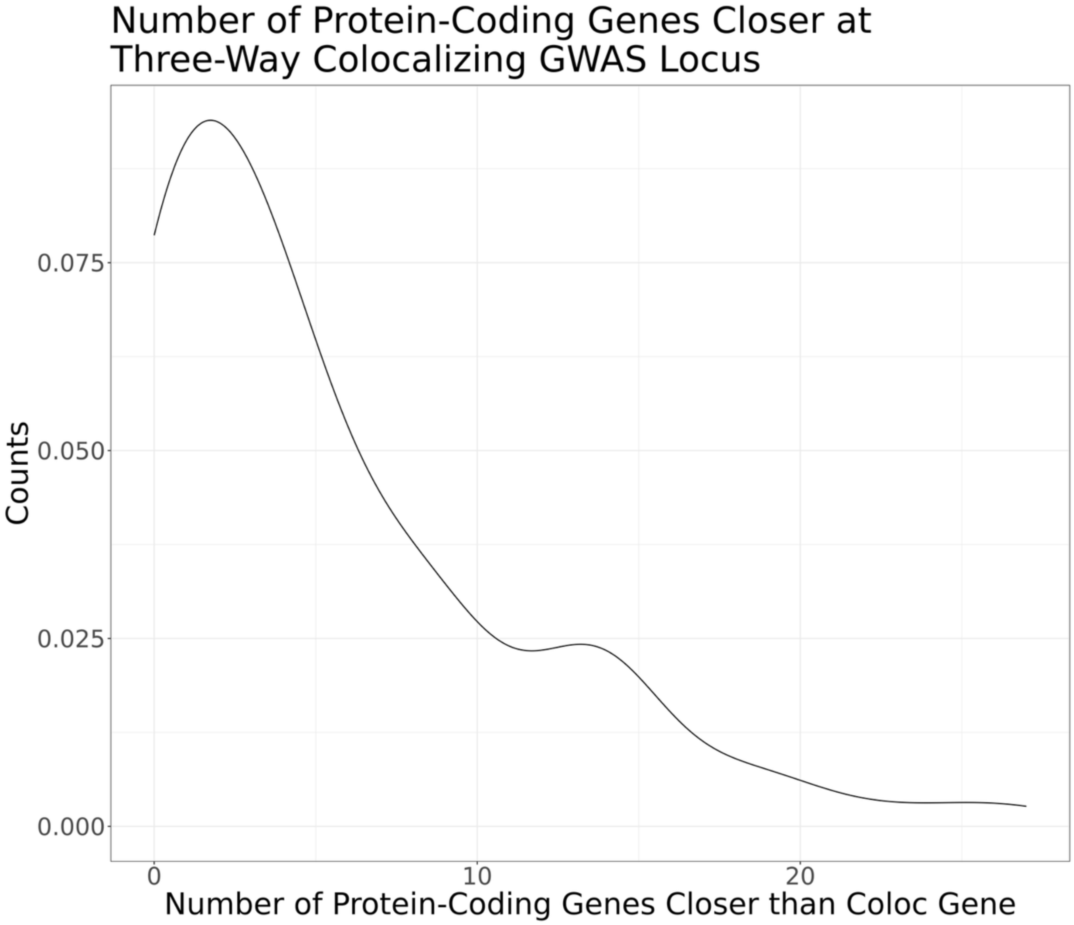


Supplementary Figure 23: A. Of the 298 three-way colocalization events involving caQTLs, GTEx and/or meta-analyzed eQTLs, and trans-ancestry blood lipids GWAS signals, 75 implicated a single gene. B. For three-way colocalizations involving a single protein-coding gene, there were a median of 5 TSSs closer than the colocalizing gene.

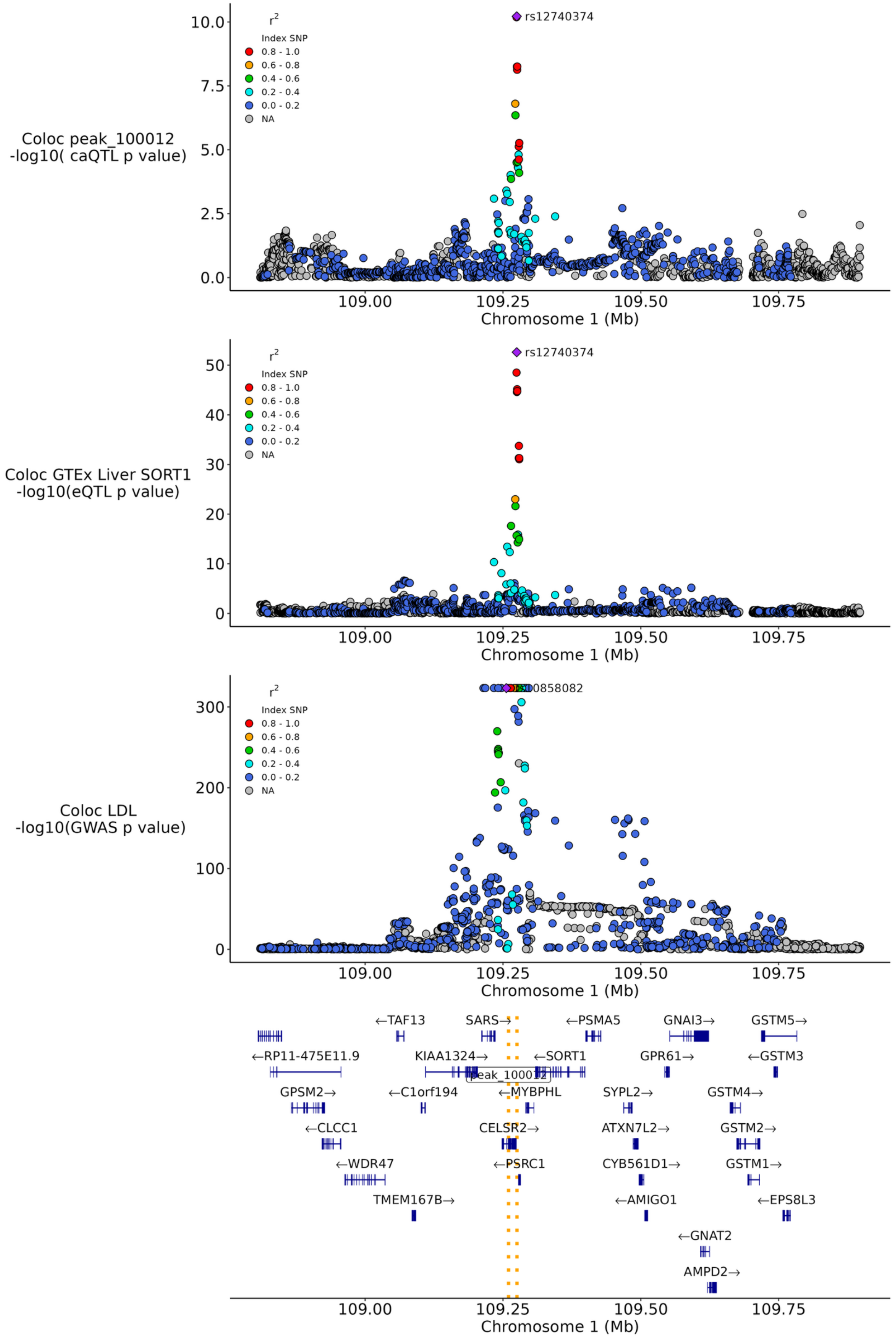


Supplementary Figure 24: Colocalization positive control locus where known LDL GWAS signal shares a causal variant with *SORT1* liver eQTL and peak 100012 in our dataset.

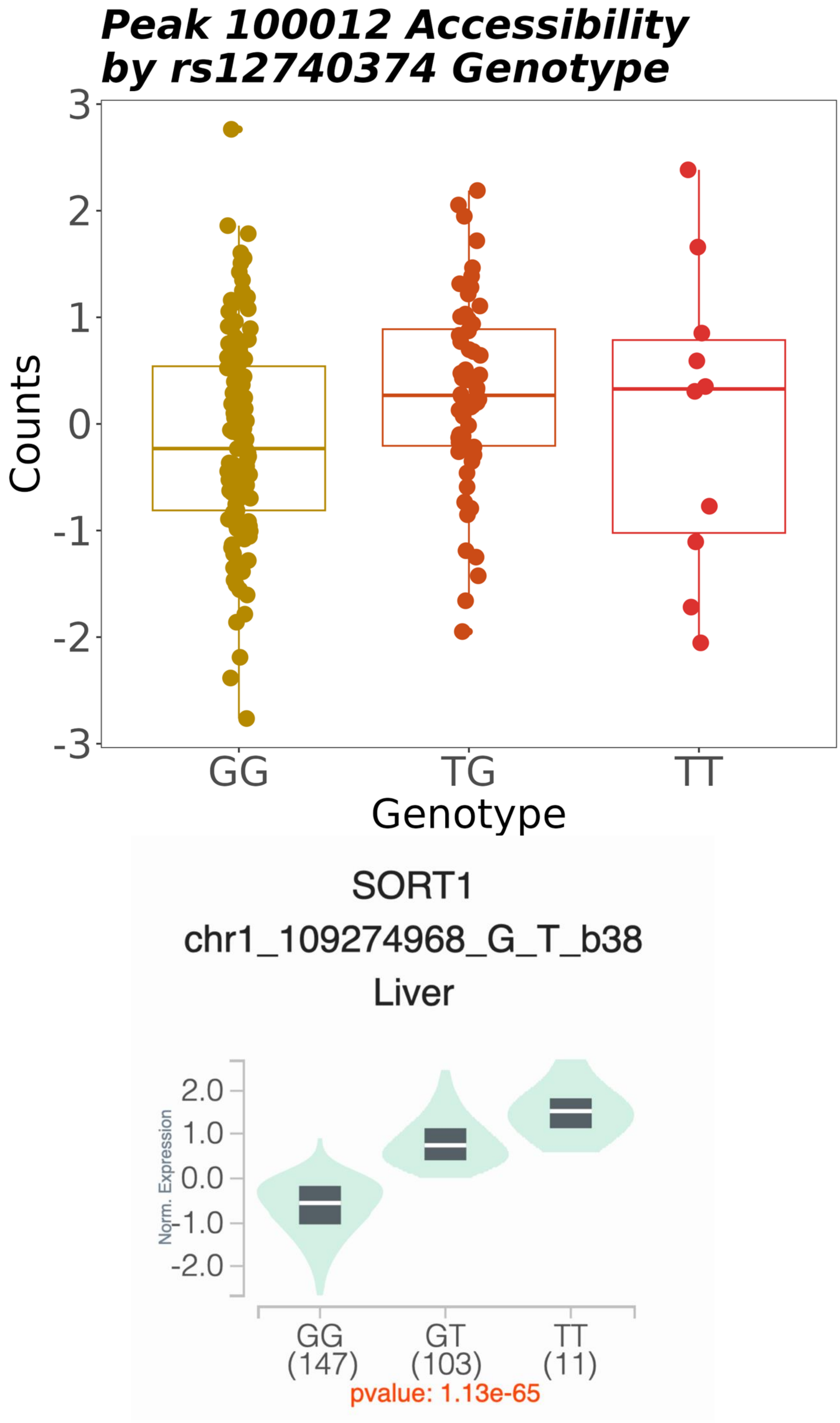


Supplementary Figure 25: Colocalization positive control locus where known LDL GWAS signal shares a causal variant with *SORT1* liver eQTL and peak 100012 in our dataset. caQTL and eQTL share effect direction.

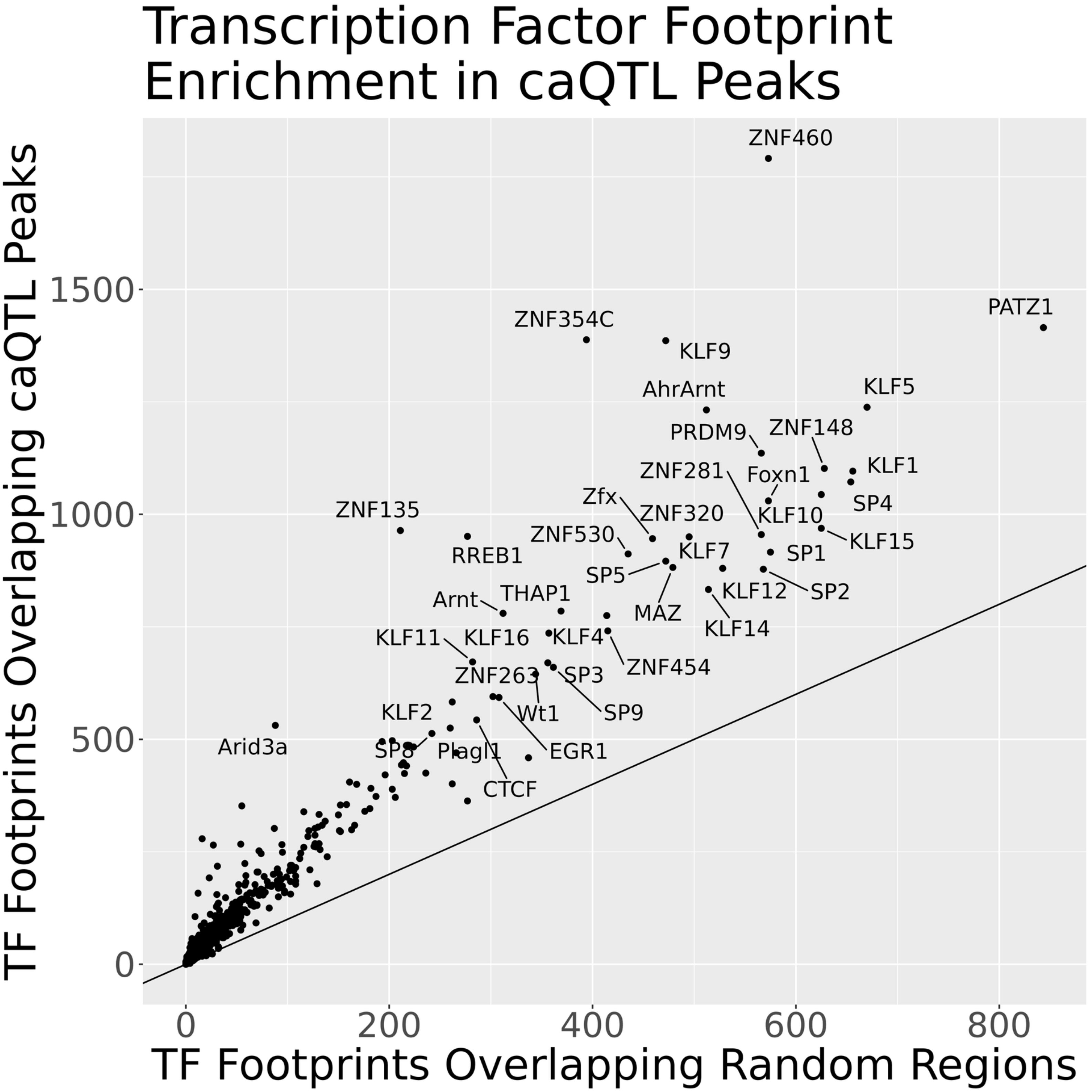


Supplementary Figure 26: TF footprints were inferred from ATAC-seq data. Enrichment of predicted TF binding was calculated in caQTL peaks compared to matched background peaks.

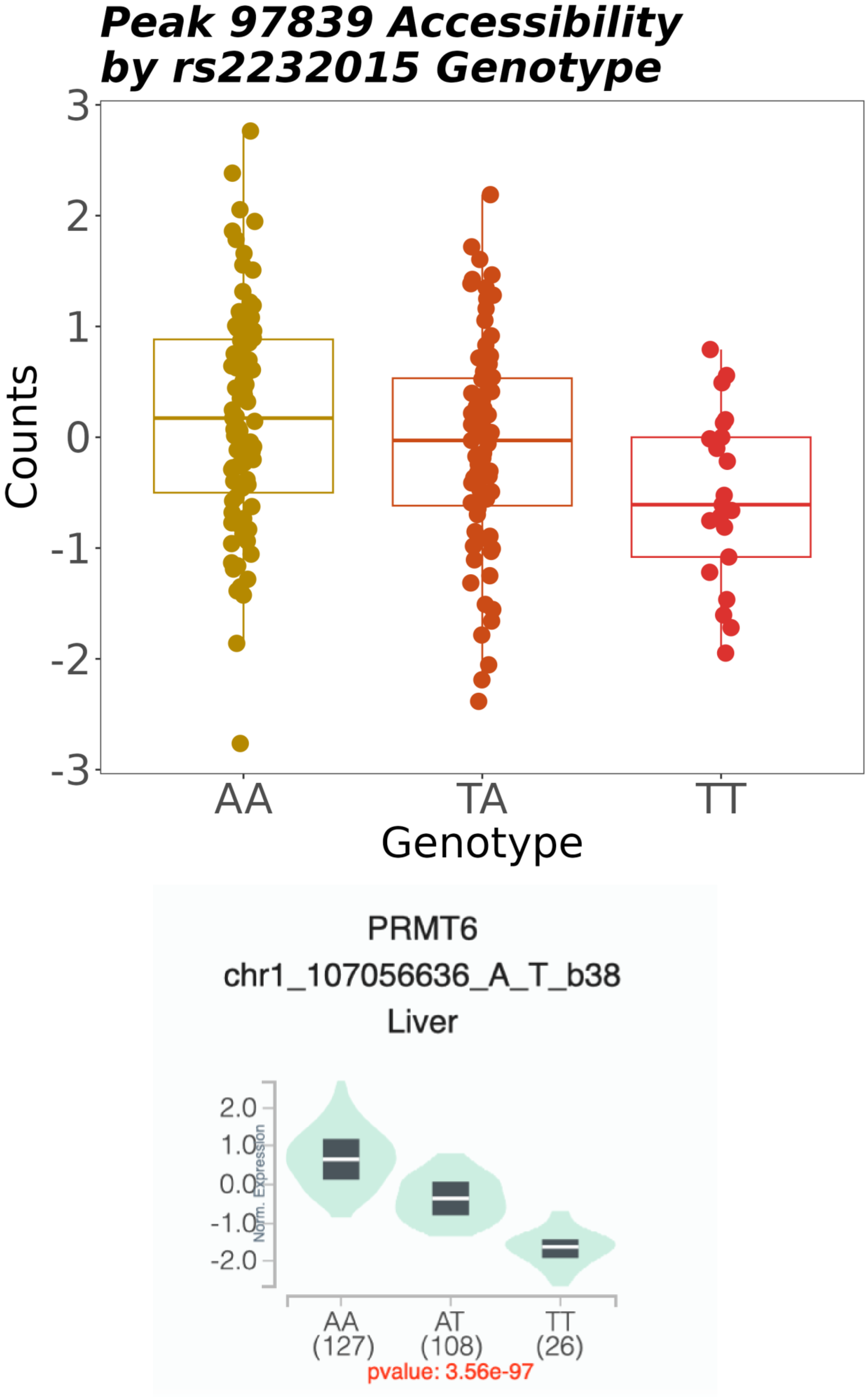


Supplementary Figure 27: Three-way colocalization that occurred between caQTL peak 97839, *PRMT6* gene, and LDL with consistent effect direction between caQTL and eQTL.

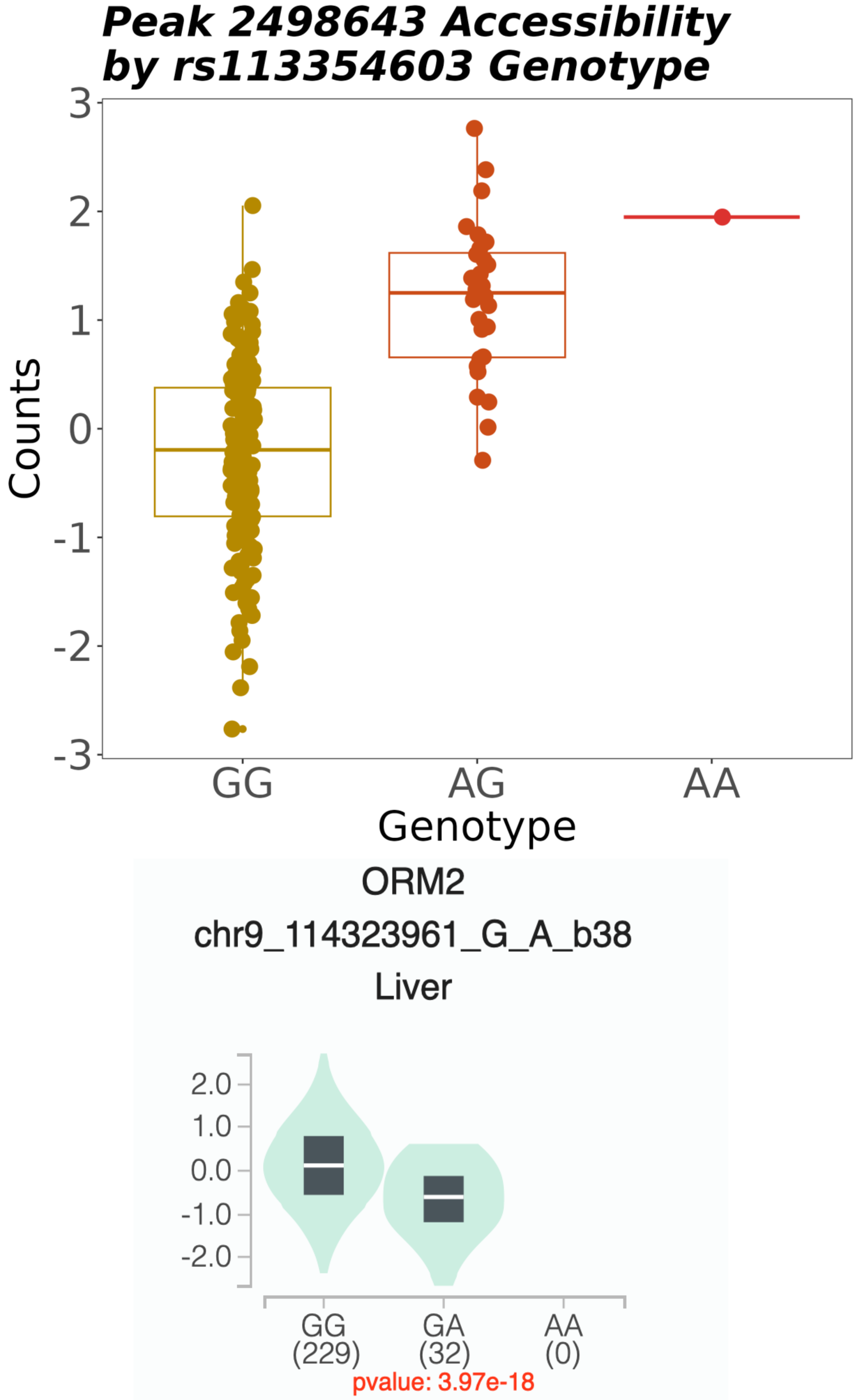


Supplementary Figure 28: Three-way colocalization that occurred between caQTL peak 2498643, *ORM2* gene, and trigylcerides with opposite effect direction between caQTL and eQTL.

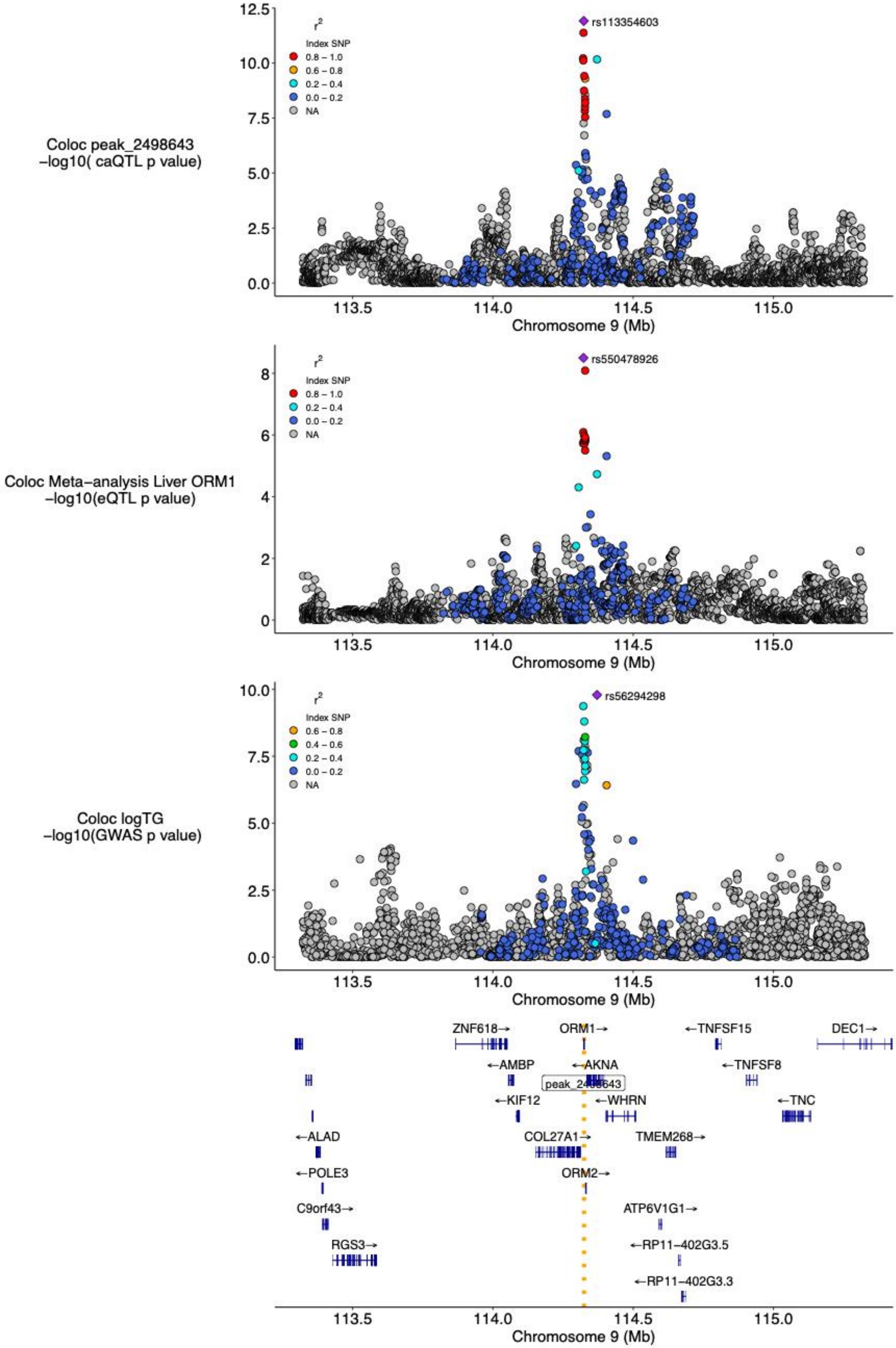


Supplementary Figure 29: Three-way colocalization locus where logTG GWAS colocalized with *ORM1* liver eQTL and peak 2498643 in our dataset.

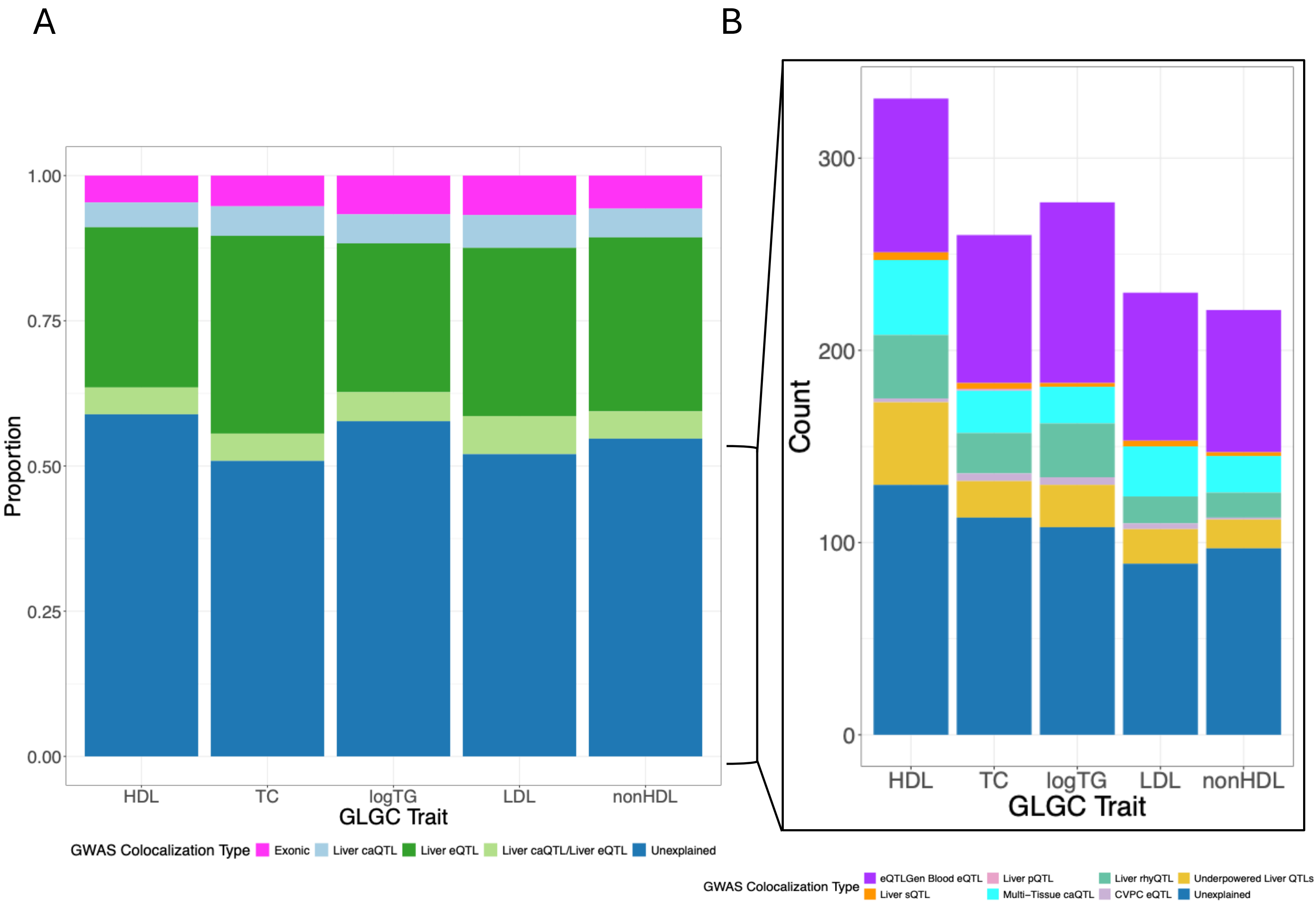


Supplementary Figure 30: (A) Coding variants, liver caQTLs/eQTLs, or both, explain approximately 45% of GWAS signals for GLGC blood lipid traits. (B) Expanding beyond coding variants and liver accessibility and gene expression mechanisms enabled for the prediction of ~33% additional GWAS signals.

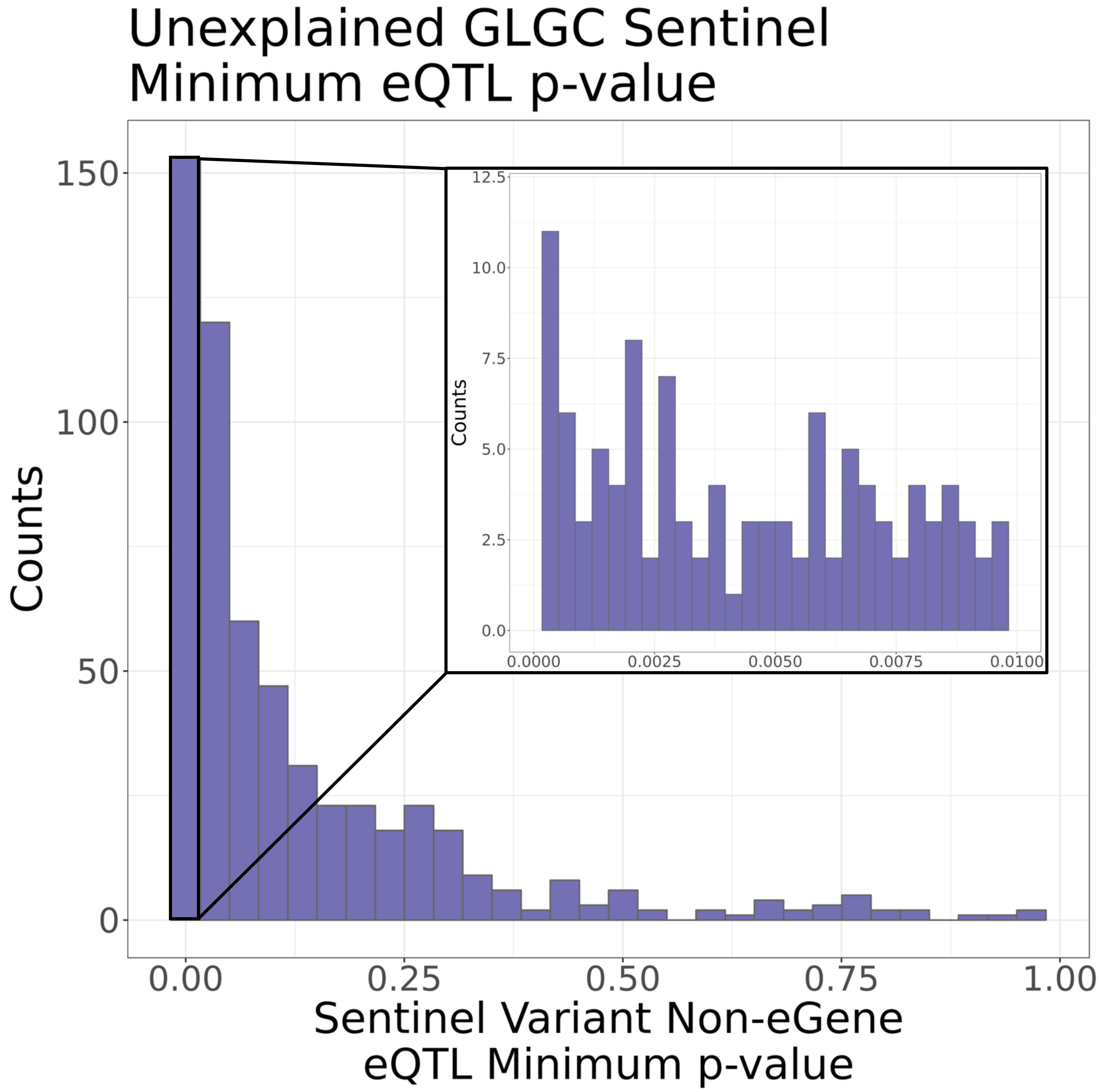


Supplementary Figure 31: The liver meta-analysis eQTL dataset was queried for GWAS sentinel variants that remained without a mechanistic explanation. The strongest signal that was not a significant eQTL was chosen and p-values are plotted. The inset shows p-values < 0.01.

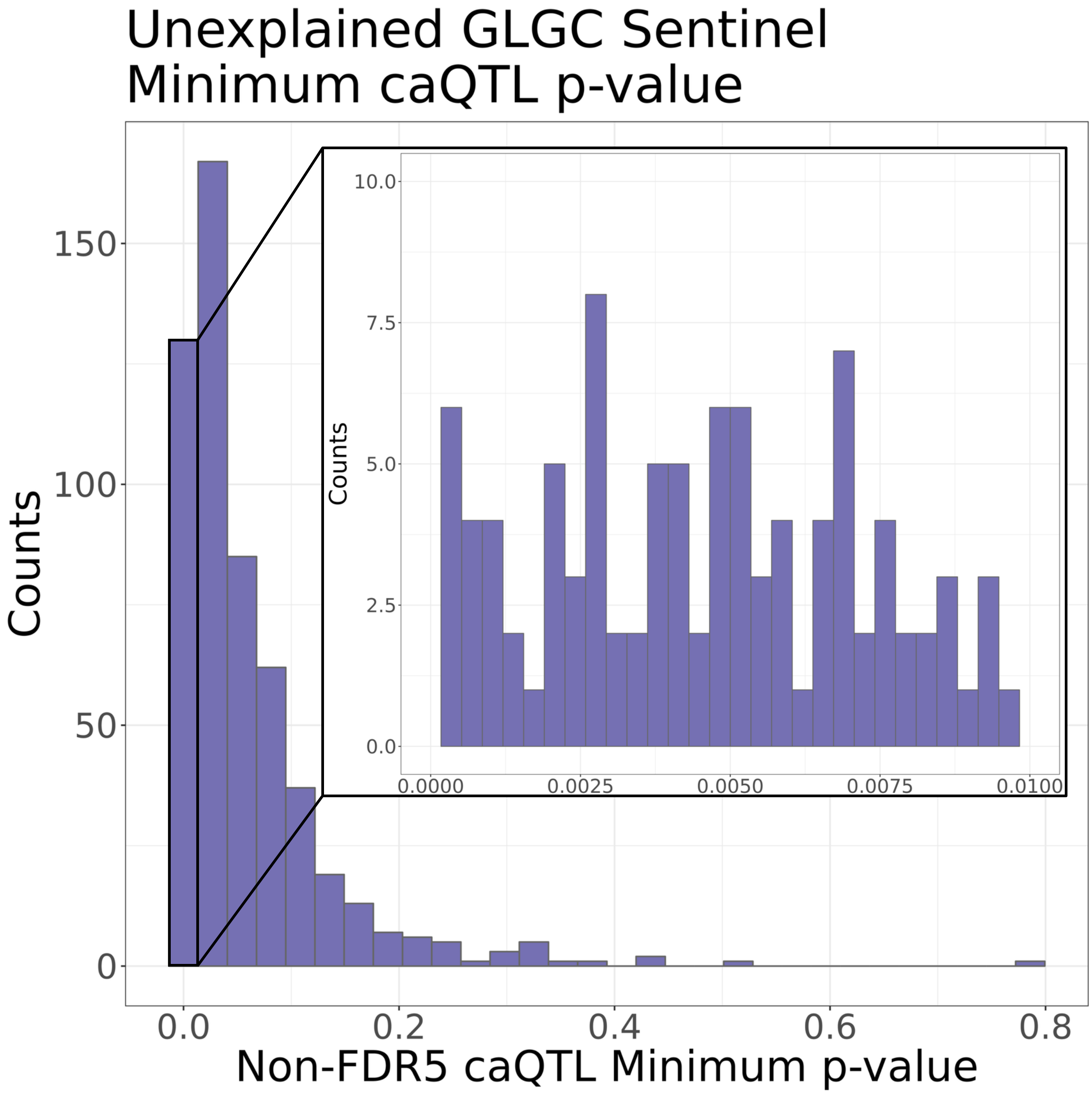


Supplementary Figure 32: The liver caQTL dataset was queried for GWAS sentinel variants that remained without a mechanistic explanation. The strongest signal that was not a significant caQTL was chosen and p-values are plotted. The inset shows p-values < 0.01.

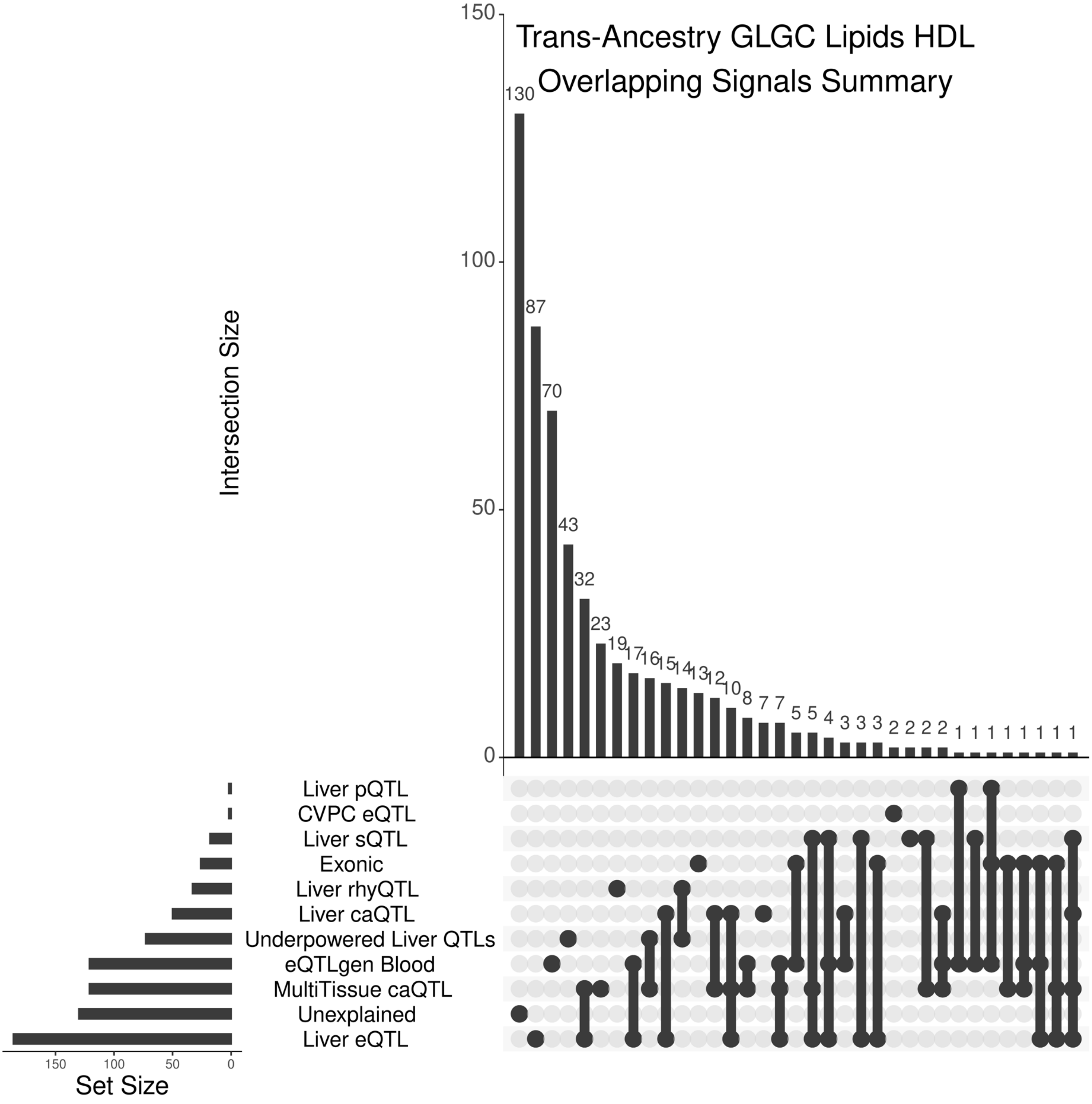


Supplementary Figure 33: Upset plot showing the overlap and sharing of various mechanistic hypotheses with sentinel GWAS variants for HDL.

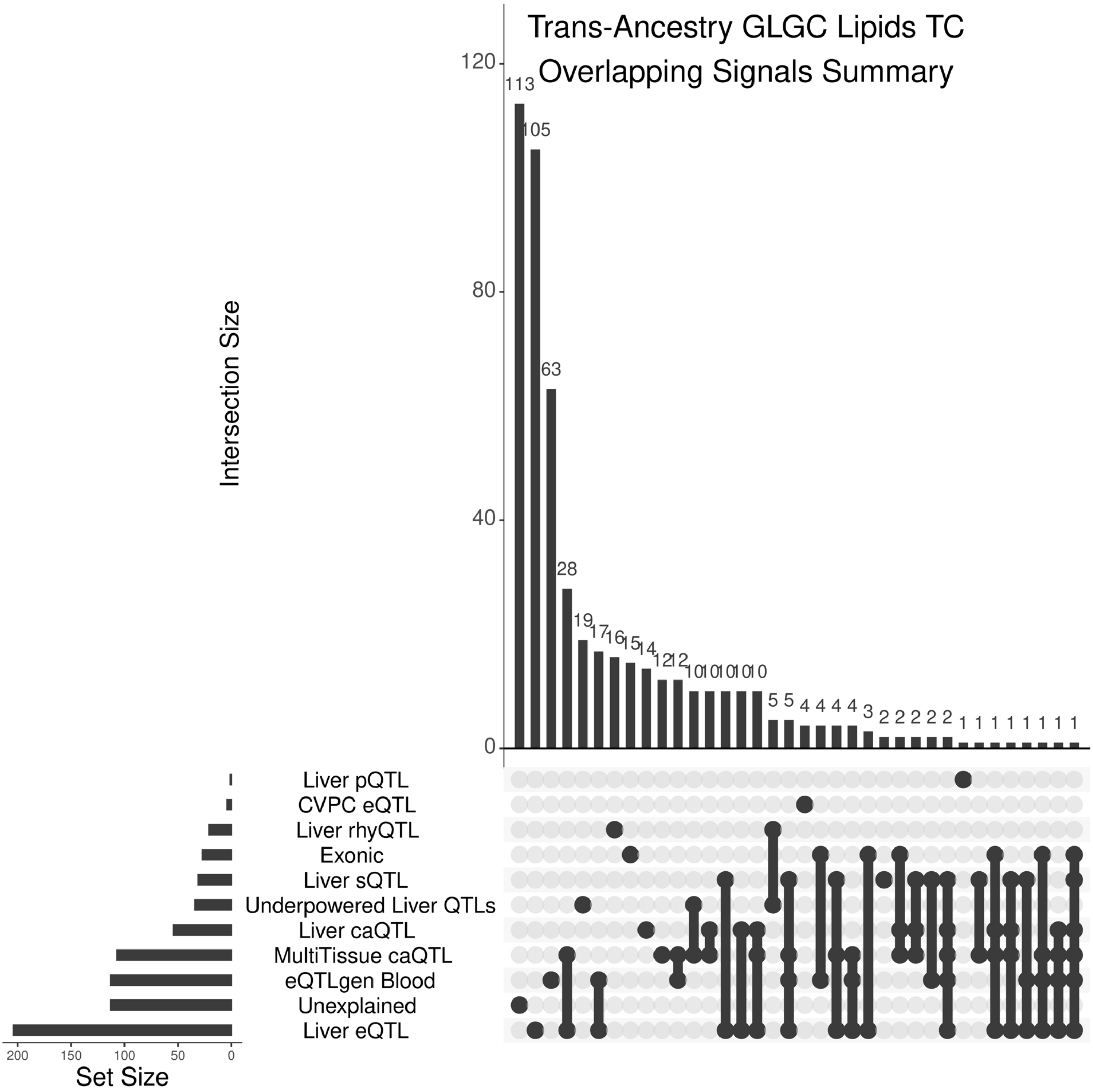


Supplementary Figure 34: Upset plot showing the overlap and sharing of various mechanistic hypotheses with sentinel GWAS variants for TC.

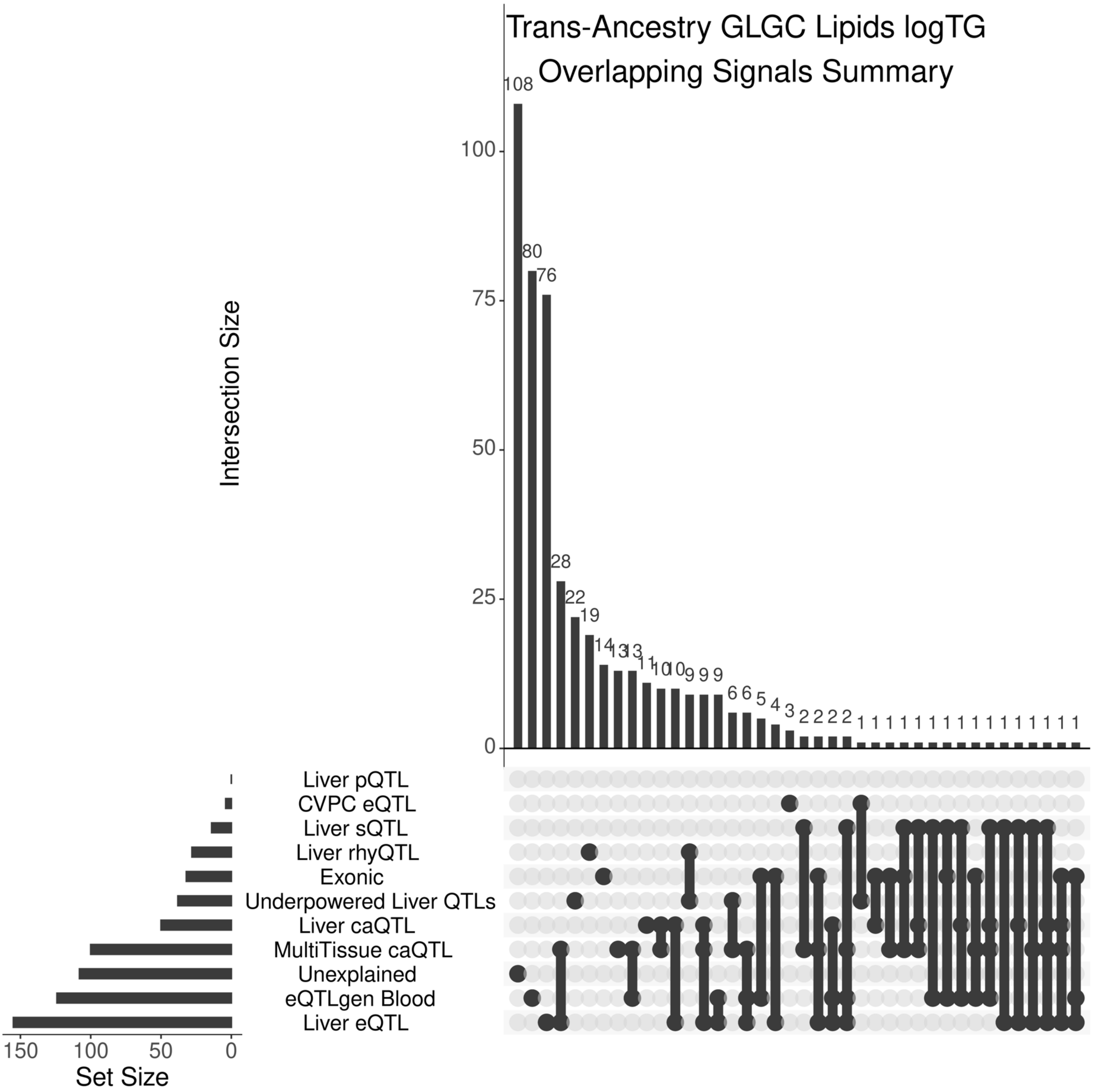


Supplementary Figure 35: Upset plot showing the overlap and sharing of various mechanistic hypotheses with sentinel GWAS variants for logTG.

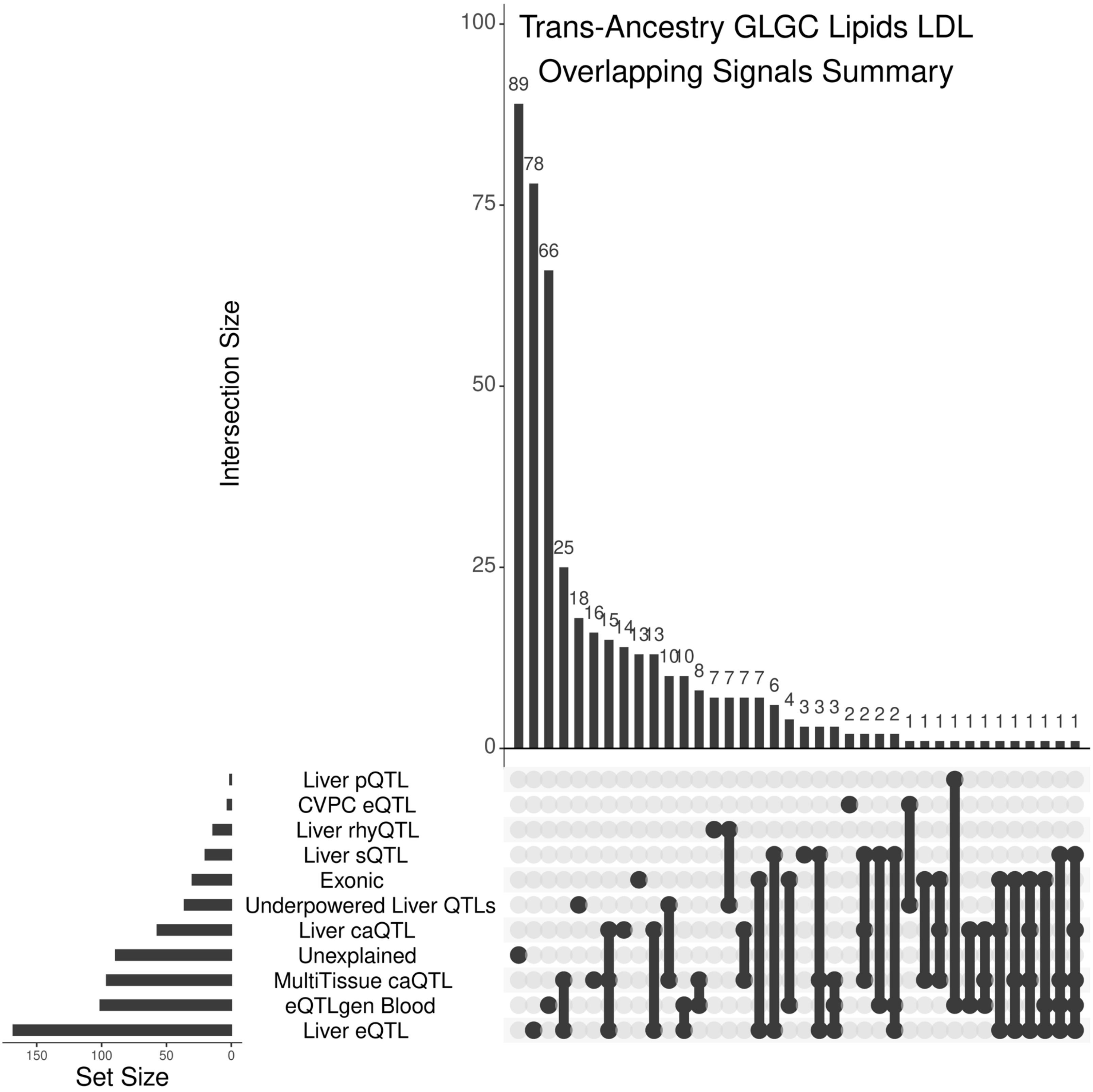


Supplementary Figure 36: Upset plot showing the overlap and sharing of various mechanistic hypotheses with sentinel GWAS variants for LDL.

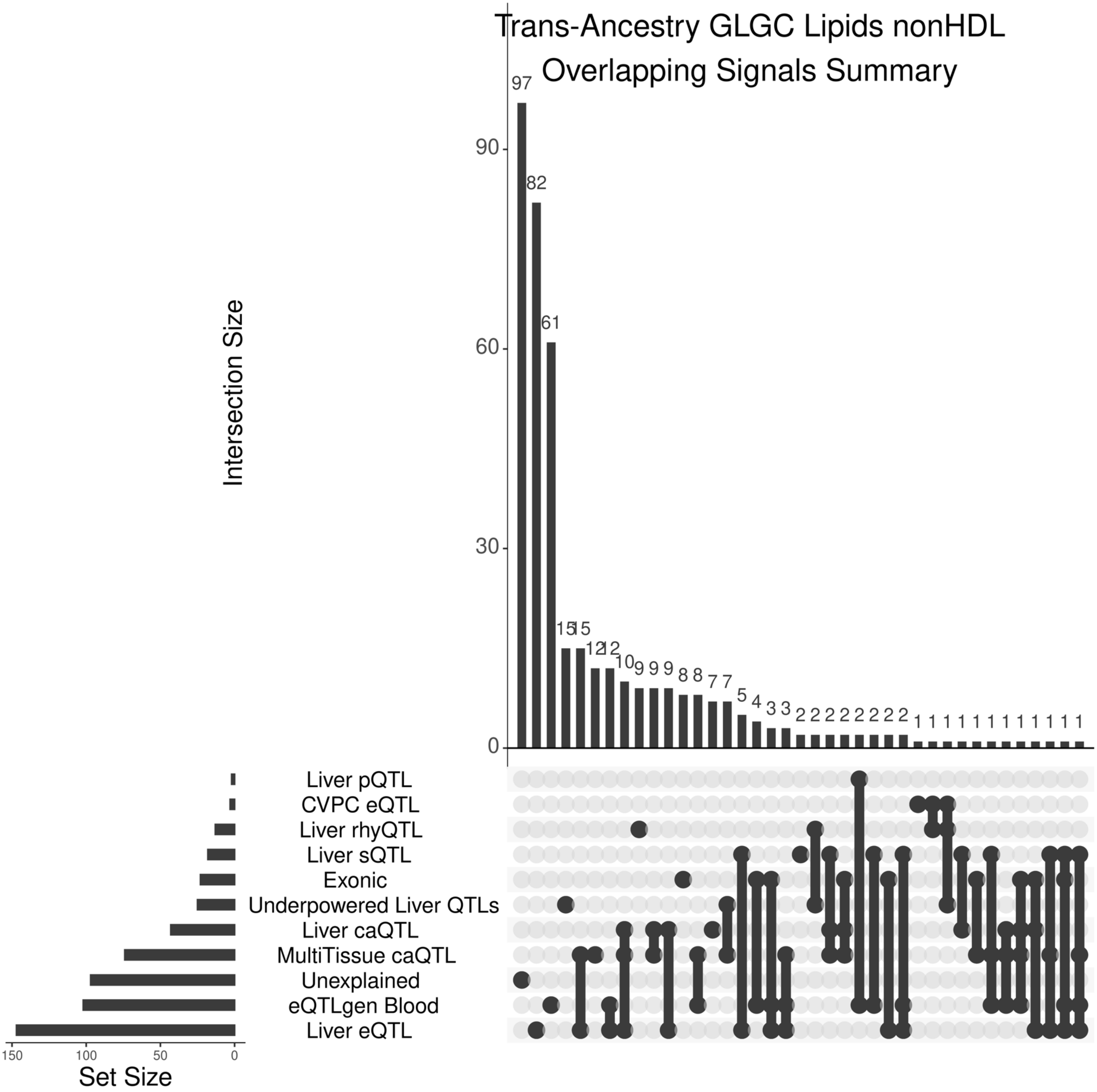


Supplementary Figure 37: Upset plot showing the overlap and sharing of various mechanistic hypotheses with sentinel GWAS variants for nonHDL.

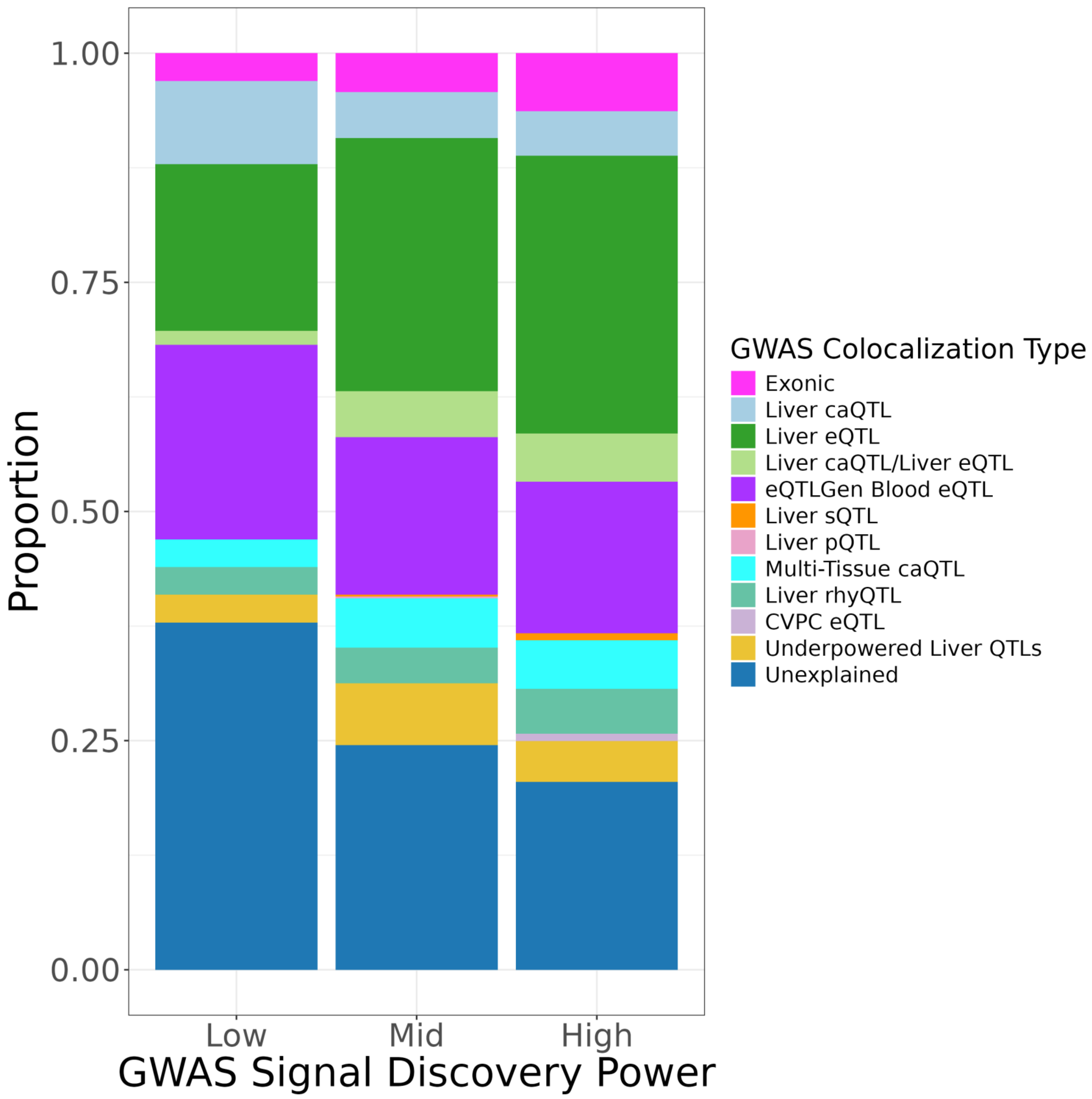


Supplementary Figure 38: We calculated the non-centrality parameter (NCP) and derived the discovery power for each GLGV GWAS sentinel variant. We split the variants into three groups based on the strength of discovery power and assessed the distribution of colocalization status within and across these power groups.

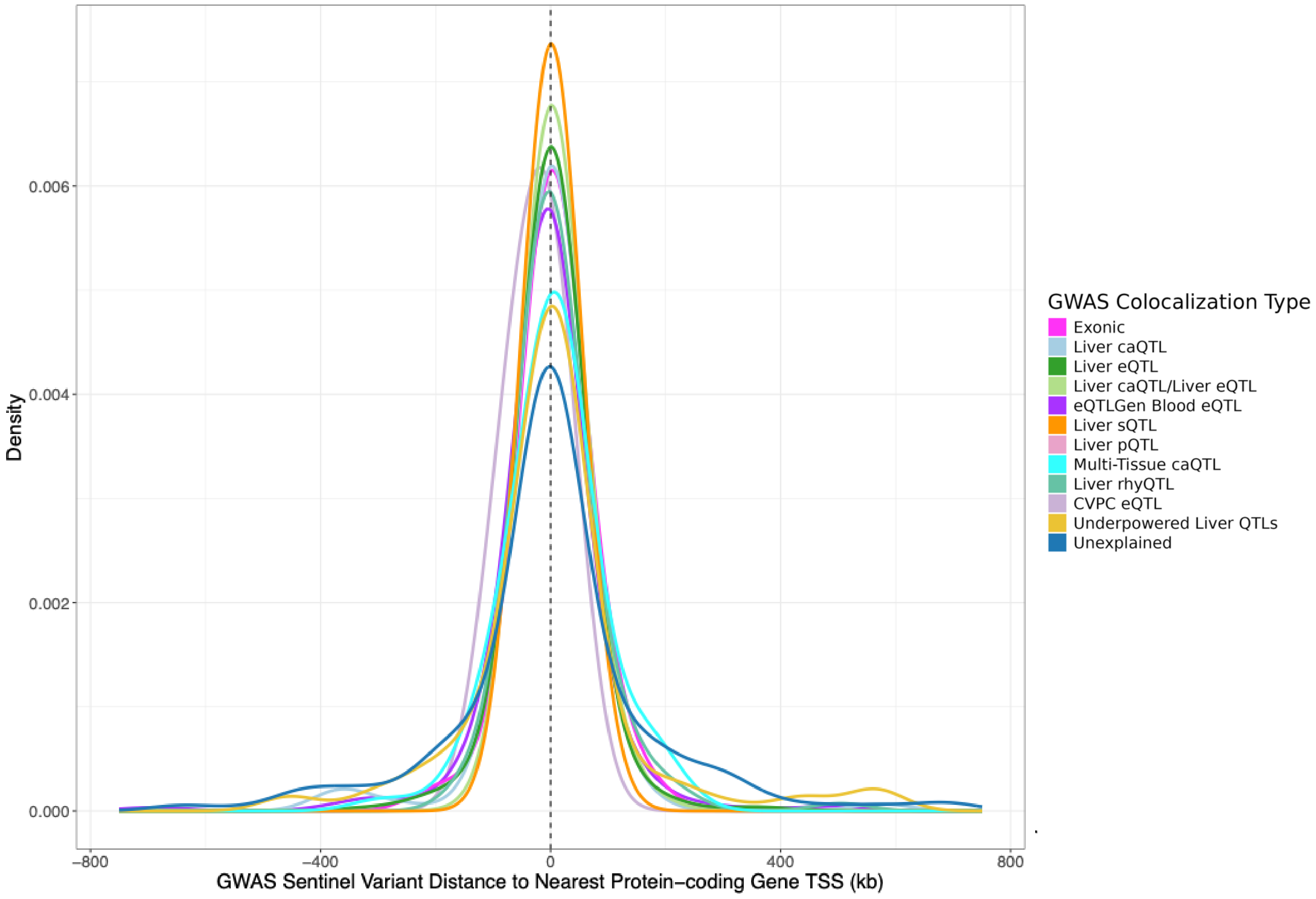


Supplementary Figure 39: For each GLGC GWAS sentinel variant, we calculated the distance to the nearest protein-coding gene TSS and plotted based on colocalization group.

| Coloc Group | Mean Value | Median Value | Min Value | Max Value | Variant Count |
|---|---|---|---|---|---|
| Liver pQTL | 14,438 | 14,438 | 14,438 | 14,438 | 1 |
| Exonic | 34,812 | 14,801 | 60 | 199,654 | 138 |
| Liver caQTL/Liver eQTL | 24,692 | 15,190 | 67 | 187,814 | 122 |
| Liver sQTL | 16,196 | 16,089 | 194 | 54,789 | 14 |
| Liver rhyQTL | 41,216 | 16,705 | 152 | 503,740 | 109 |
| Liver eQTL | 34,348 | 16,823 | 39 | 423,491 | 701 |
| CVPC eQTL | 34,111 | 19,252 | 216 | 85,030 | 14 |
| Liver caQTL | 44,029 | 21,471 | 67 | 573,456 | 119 |
| eQTLGen Blood eQTL | 50,604 | 22,374 | 10 | 720,117 | 402 |
| Underpowered Liver QTLs | 93,271 | 37,534 | 24 | 932,040 | 117 |
| Multi-Tissue caQTL | 63,740 | 40,355 | 298 | 830,417 | 125 |
| Unexplained | 126,256 | 46,290 | 39 | 1,529,838 | 537 |

Supplementary Figure 40: For each GLGC GWAS sentinel variant, we calculated the distance to the nearest protein-coding gene TSS and calculated statistics based on colocalization group.

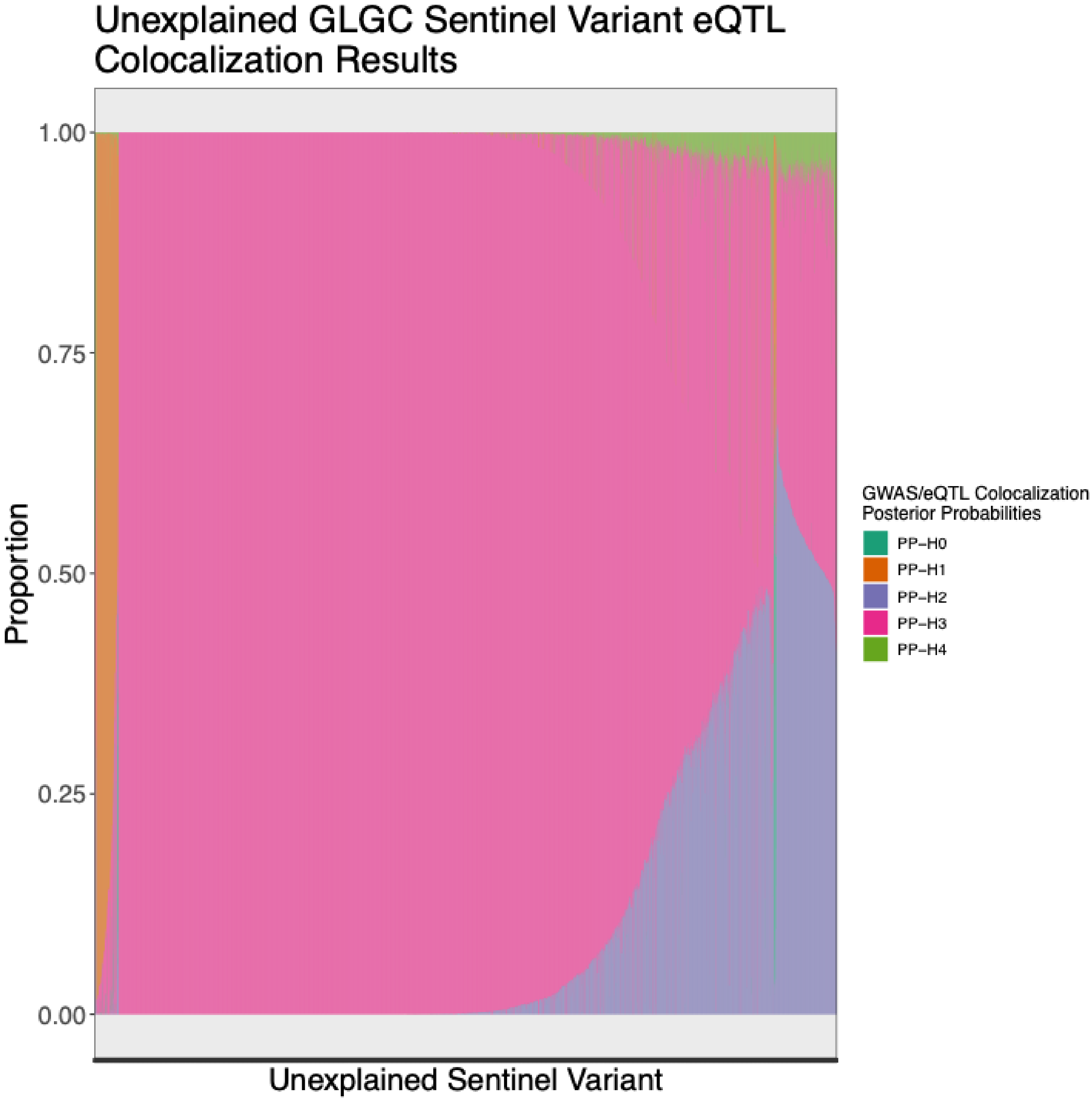


Supplementary Figure 41: 620 GLGC GWAS signals remained without a proposed mechanism after colocalizations with multiple molecular traits. We plotted the liver eQTL colocalization posterior probabilities for these signals.

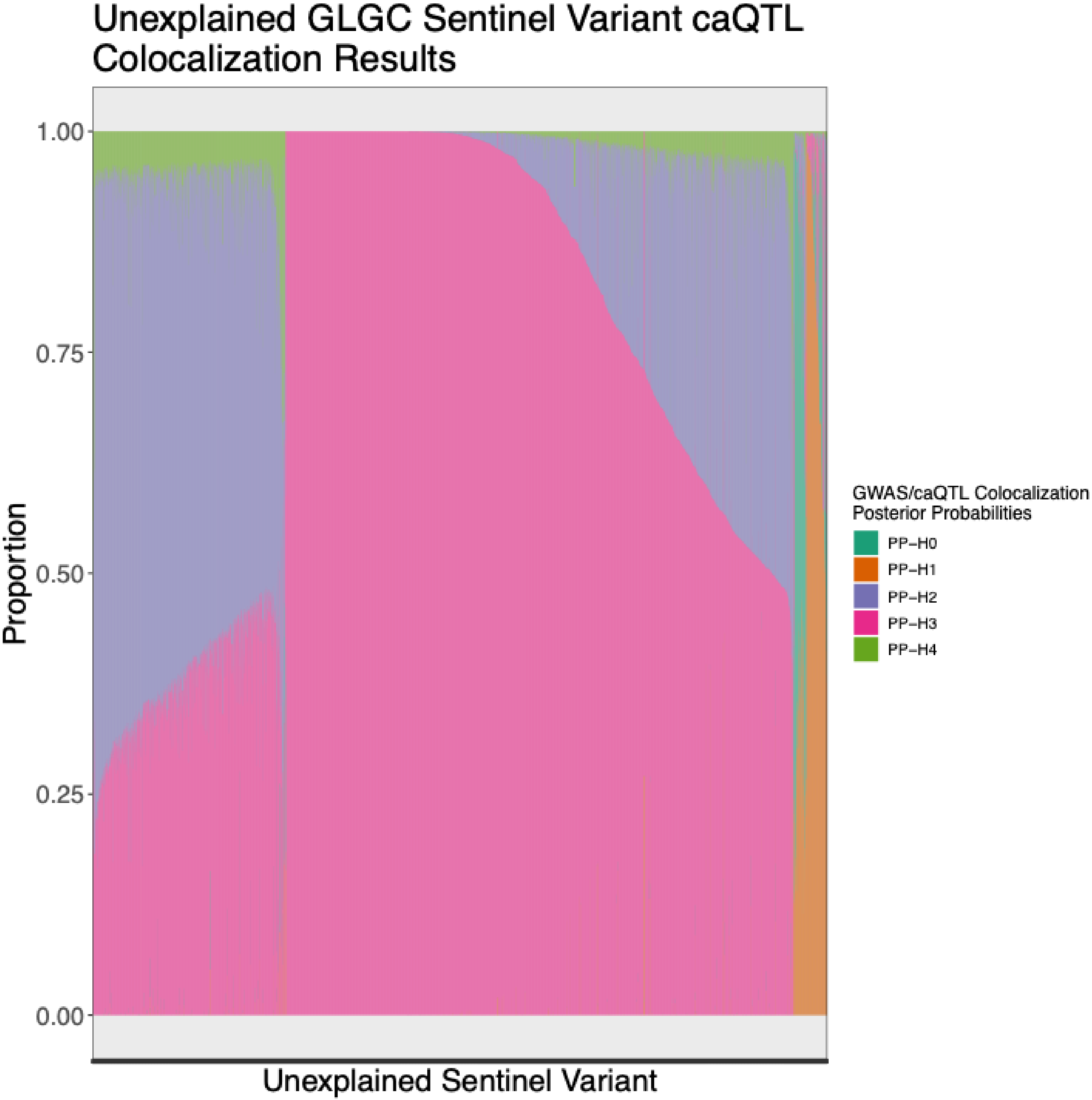


Supplementary Figure 42: 620 GLGC GWAS signals remained without a proposed mechanism after colocalizations with multiple molecular traits. We plotted the liver caQTL colocalization posterior probabilities for these signals.

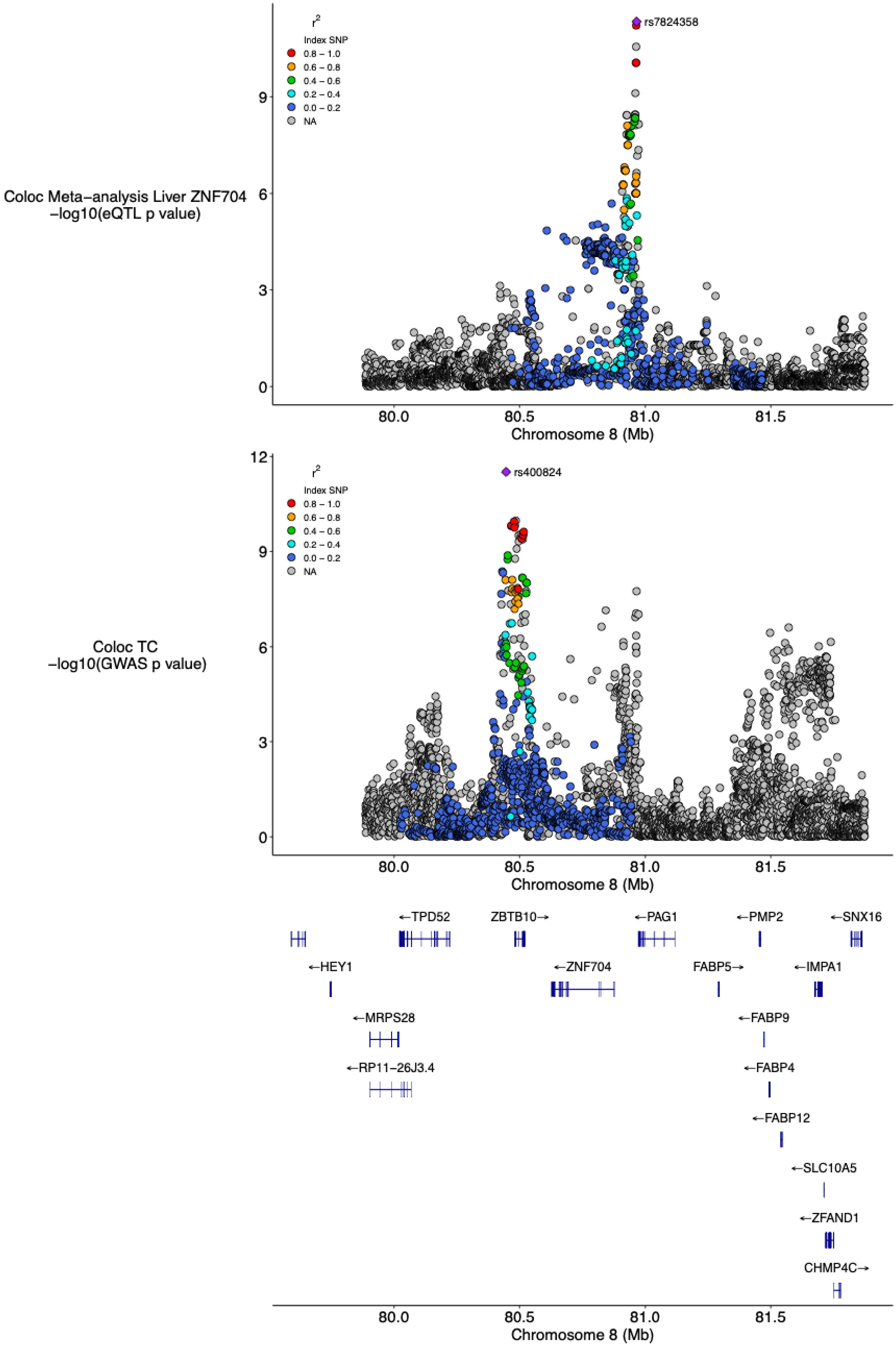


Supplementary Figure 43: Locus where GWAS and eQTL both have significant signals, but do not colocalize (PP-H3 = 0.86) and multiple signals appear to be present. Lead eQTL (rs2044397) and lead GWAS signal (rs400824) are not in strong LD ($r^2 = 0.0013$).

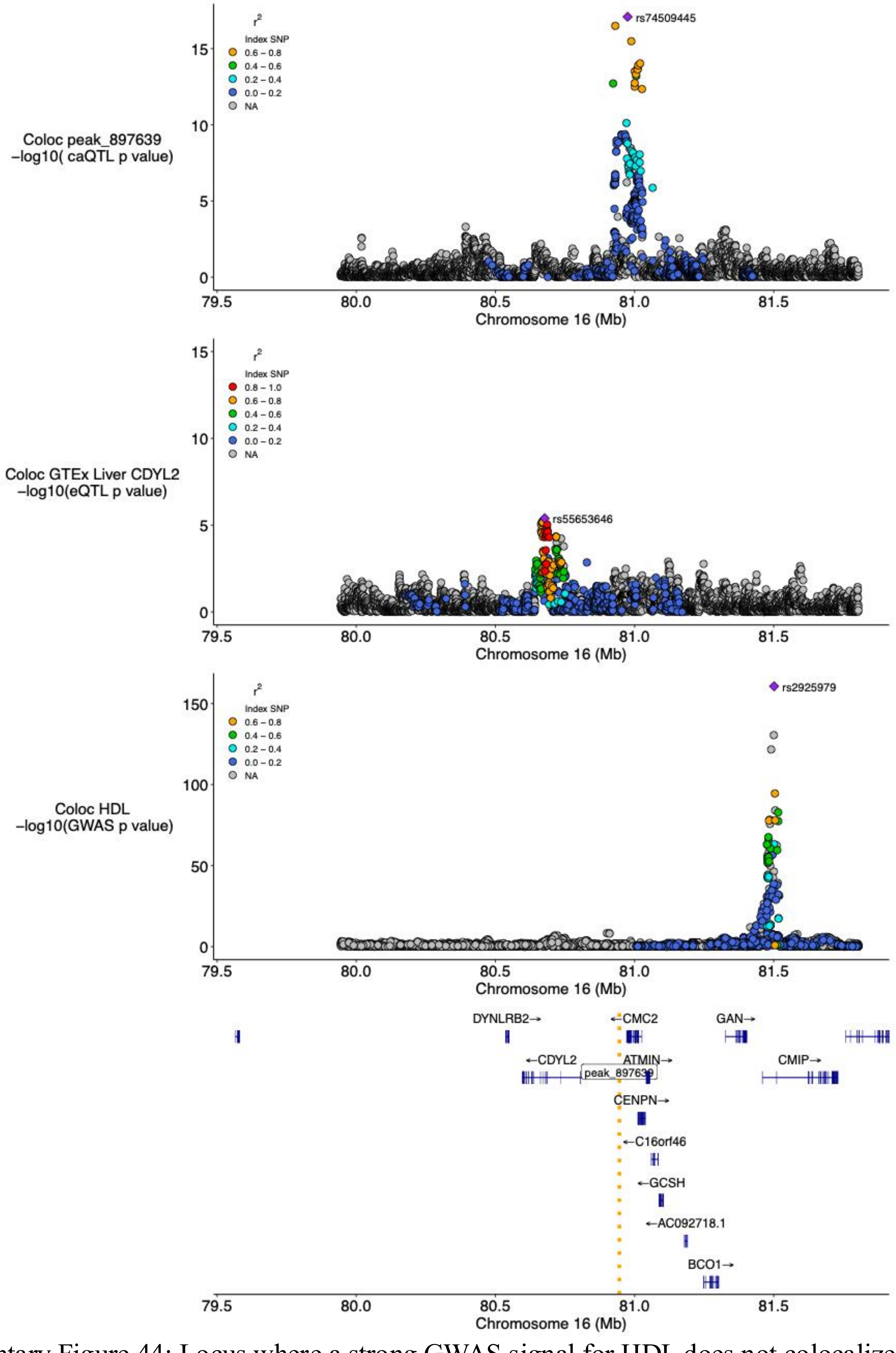


Supplementary Figure 44: Locus where a strong GWAS signal for HDL does not colocalize with liver caQTL and/or liver eQTL. In addition, lead signals for each trait were not in strong linkage disequilibrium (LD) with $R^2$ of 0.0004 between GWAS/eQTL lead signals and $R^2$ of 0.0002 between GWAS/caQTL lead signals. This locus additionally did not colocalize with any other potential exonic or regulatory mechanisms investigated. Loci such as this likely need larger sample sizes in liver epigenomic datasets, or experiments from a different developmental timepoint, cell type, and/or cellular context.

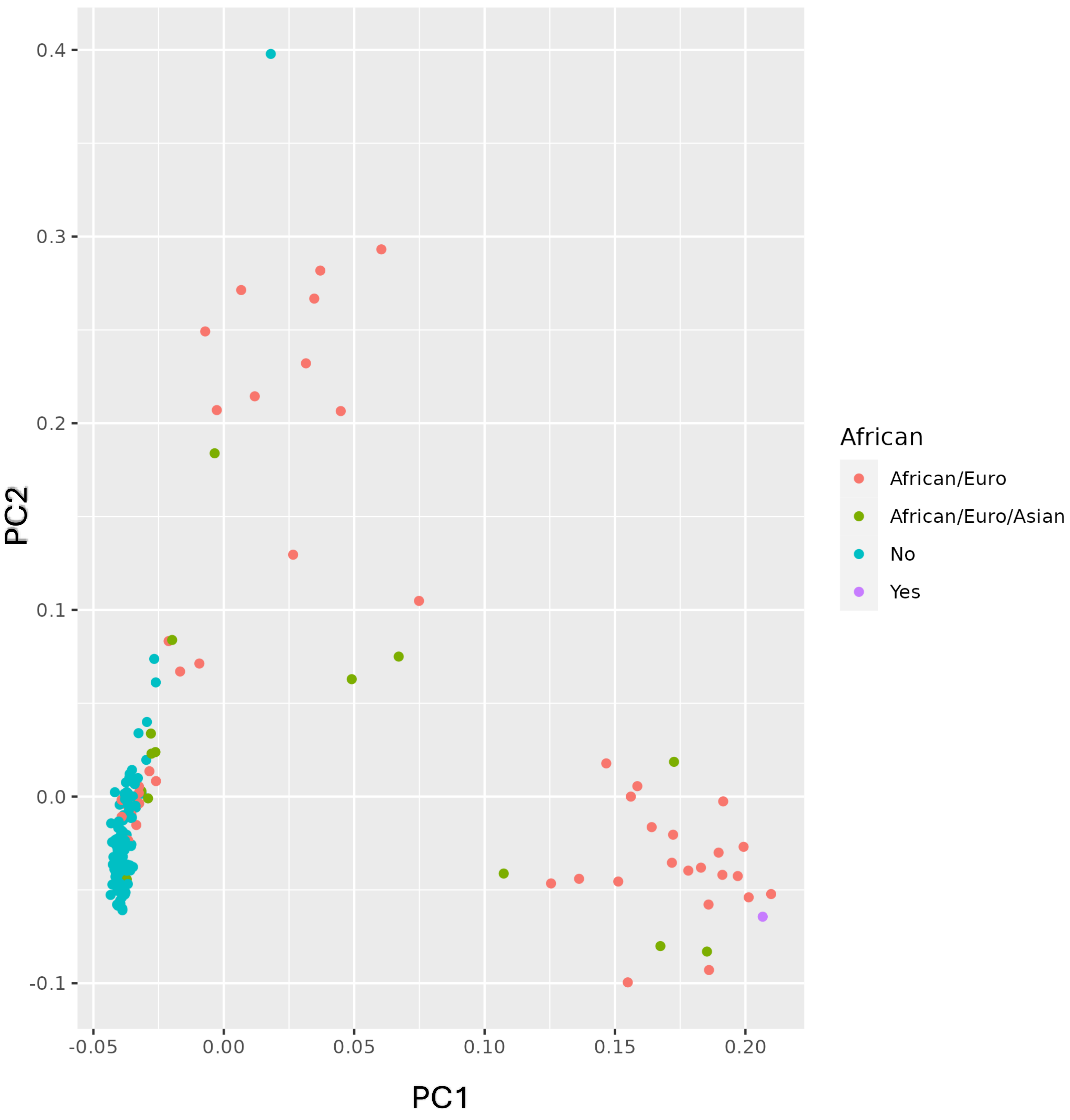


Supplementary Figure 45: Genotype PCA labeled with summarized Gencove reported ancestries. Yes = only African ancestry; No = no African ancestry; African/Euro = both African and European ancestry; African/Euro/Asian = multiple ancestries.

### Three Populations

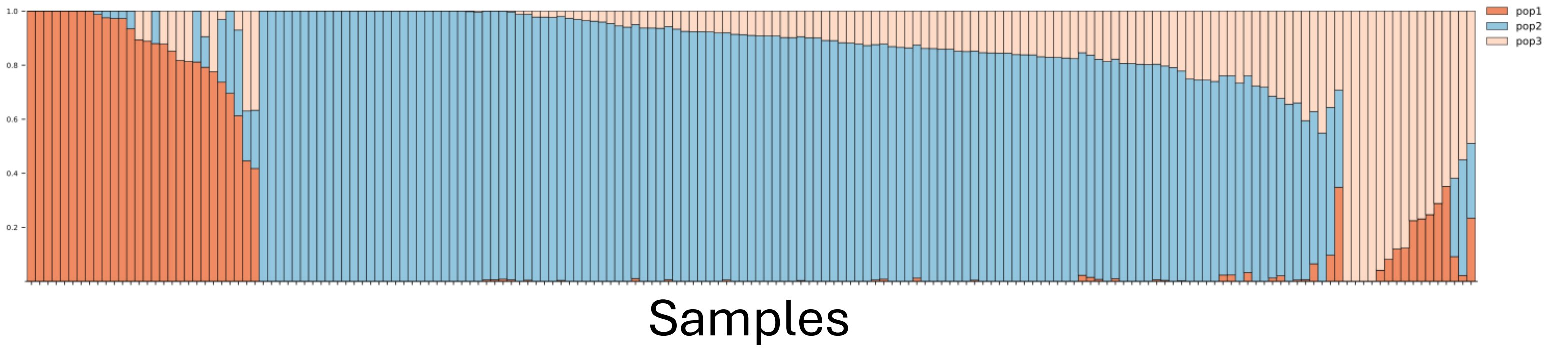


Supplementary Figure 46: ADMIXTURE cross validation was performed for K=1-5. Cross validation error was minimized with K=3.

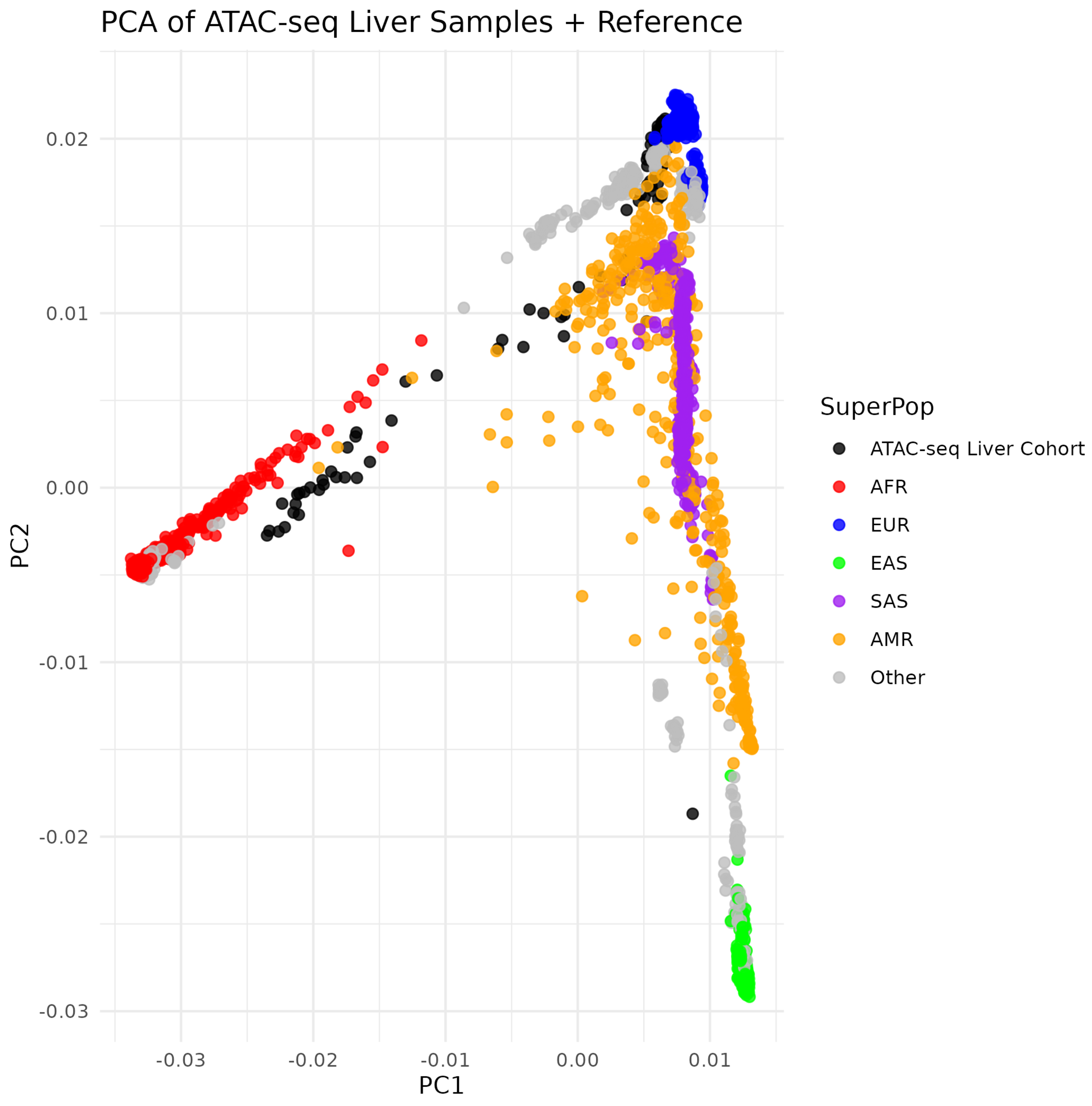


Supplementary Figure 47: 1000 Genomes genotype data were downloaded and merged with liver samples. PCA on genotypes shows groupings of liver samples in ancestral population groups. Liver samples with PC1 < -0.01 were characterized as “Non-European”.
